# CoMix: Changes in social contacts as measured by the contact survey during the COVID-19 pandemic in England between March 2020 and March 2021

**DOI:** 10.1101/2021.05.28.21257973

**Authors:** Amy Gimma, James D Munday, Kerry LM Wong, Pietro Coletti, Kevin van Zandvoort, Kiesha Prem, CMMID COVID-19 working group, Petra Klepac, G. James Rubin, Sebastian Funk, W John Edmunds, Christopher I Jarvis

**Affiliations:** Centre for Mathematical Modelling of Infectious Diseases, Department of Infectious Disease Epidemiology, London School of Hygiene and Tropical Medicine, Keppel Street, WC1E 7HT London, UK; UHasselt, Data Science Institute and I-BioStat, Hasselt 3500, Belgium; Department of Psychological Medicine, King’s College London, Denmark Hill, London, UK

**Keywords:** Covid-19, Contact survey, Pandemic, Disease Outbreak, Reproduction Number, CoMix

## Abstract

**Background:** During the COVID-19 pandemic, the UK government imposed public health policies in England to reduce social contacts in hopes of curbing virus transmission. We measured contact patterns weekly from March 2020 to March 2021 to estimate the impact of these policies, covering three national lockdowns interspersed by periods of lower restrictions.

**Methods:** Data were collected using online surveys of representative samples of the UK population by age and gender. We calculated the mean daily contacts reported using a (clustered) bootstrap and fitted a censored negative binomial model to estimate age-stratified contact matrices and estimate proportional changes to the basic reproduction number under controlled conditions using the change in contacts as a scaling factor.

**Results:** The survey recorded 101,350 observations from 19,914 participants who reported 466,710 contacts over 53 weeks. Contact patterns changed over time and by participants’ age, personal risk factors, and perception of risk. The mean of reported contacts among adults have reduced compared to previous surveys with adults aged 18 to 59 reporting a mean of 2.39 (95% CI 2.20 - 2.60) contacts to 4.93 (95% CI 4.65 - 5.19) contacts, and the mean contacts for school-age children was 3.07 (95% CI 2.89 - 3.27) to 15.11 (95% CI 13.87 - 16.41). The use of face coverings outside the home has remained high since the government mandated use in some settings in July 2020.

**Conclusions:** The CoMix survey provides a unique longitudinal data set for a full year since the first lockdown for use in statistical analyses and mathematical modelling of COVID-19 and other diseases. Recorded contacts reduced dramatically compared to pre-pandemic levels, with changes correlated to government interventions throughout the pandemic. Despite easing of restrictions in the summer of 2020, mean reported contacts only returned to about half of that observed pre-pandemic.

## Background

Since early 2020, governments across the world have asked or required people to change their behaviour in an attempt to slow transmission of the SARS-CoV-2 virus. In England, the government has implemented a variety of measures over the course of the pandemic, including three separate national “lockdowns” [1–5] as well as other local and national measures [6]. In addition, guidance has been issued on risk mitigation measures during social interactions, including meeting outdoors, maintaining space between people, frequent handwashing or use of hand sanitiser, and the use of face coverings (masks).

We conducted a weekly longitudinal survey on the social contacts, behaviours, and attitudes of people in the UK to quantify social interactions over time. We have previously described early findings during the first week of lockdown in England (24th to 27th March 2020) [7]. In this paper, we describe observed contact patterns and behaviour in England based on the CoMix social contact survey collected between 24th March 2020 (the first day of the first national lockdown in the UK) and 29th March 2021 (the final day of the third national lockdown in England). We present descriptive analyses showing the mean number of contacts people reported and how these differed during three national lockdowns, periods with more relaxed restrictions, and over the Christmas holiday period. We provide a one-year detailed longitudinal account of contact behaviour in England during the first year of the pandemic, create a historical record for future study and policy-making, and improve understanding of the patterns of disease spread and the effectiveness of different policies on reducing contacts to suppress transmission.

## Methods

### Ethics statement

Participation in this opt-in study was voluntary, and all analyses were carried out on anonymised data. The study was approved by the ethics committee of the London School of Hygiene & Tropical Medicine Reference number 21795.

### Study design

CoMix is an international online behavioural survey that has been running weekly since it launched on the 24th of March 2020. In the UK, participants are invited to the survey and subsequently asked to respond once every two weeks, with two panels of participants who respond in alternating weeks. Initially, each panel consisted of roughly 1,500 participants, increasing to about 2,500 participants each week from August 2020 (Figure 1D). In May 2020, we launched two additional panels (each of approximately 500 participants) designed to collect data on children’s contact patterns. Parents completed the surveys on behalf of one of their children (<18 years old) who lived in the same household, based on which child had the closest upcoming birthday.

**Figure 1.**
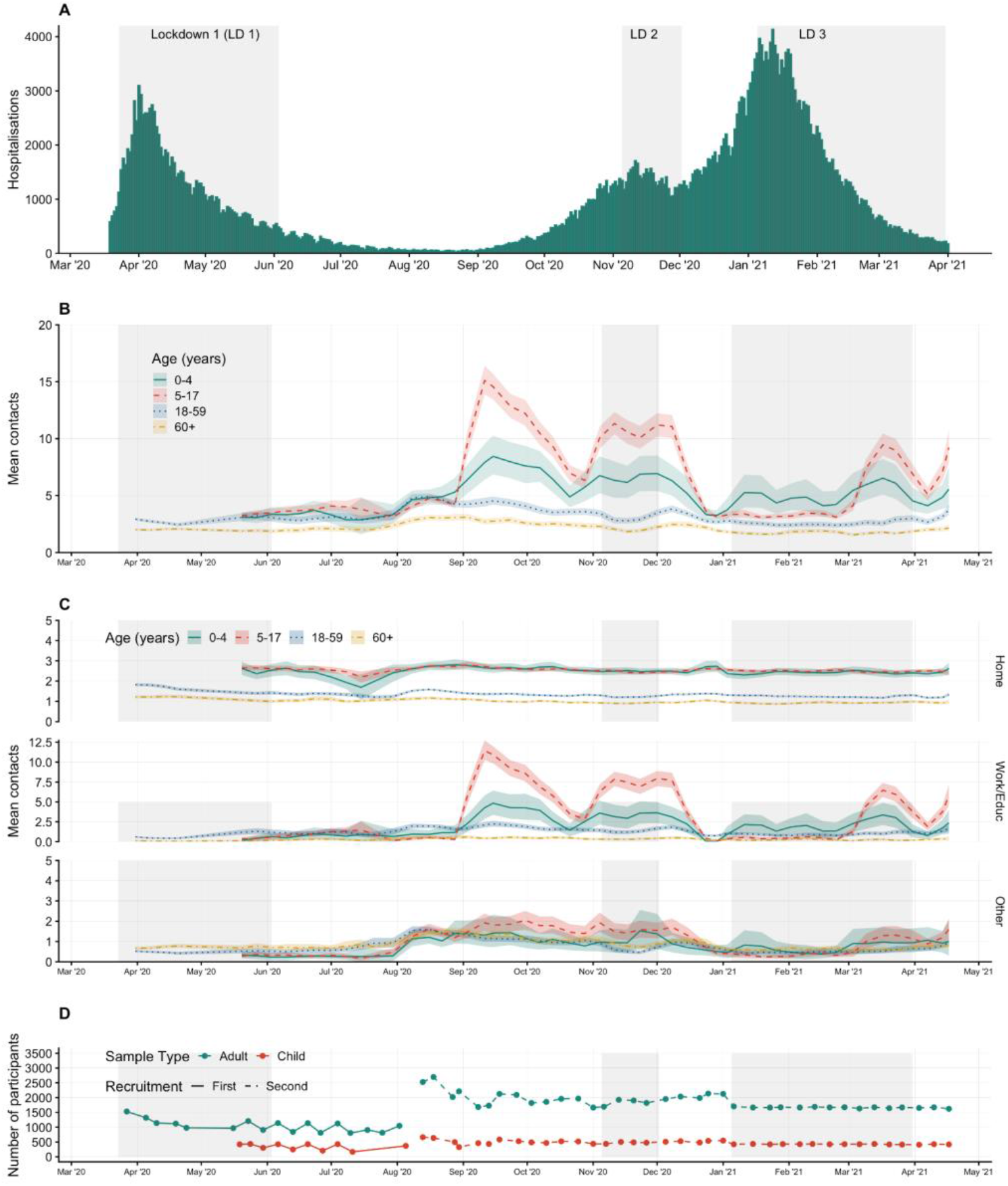
Mean contacts over time by age, and by age and setting with timeline of survey participation with 95% confidence interval of bootstrapped mean. A) Hospitalisations due to COVID-19 in England; B) Mean contacts and 95% bootstrapped confidence intervals in adults and children in age groups of 0 to 4, 5 to 17, 18 to 59, and 60 or more year; C) Mean contacts and 95% bootstrapped confidence intervals by age group and setting; and D) The number of participants and when they respond by panel over time.

A UK-representative sample was recruited by the market research company Ipsos-MORI using quota sampling, with quotas based on age, gender, and region. Ipsos-MORI recruits through a combination of social media and web advertising and email campaigns, and partners with other companies when necessary to meet quotas. New panels were recruited in August 2020, after the initially planned period of the study was completed and there was a high turnover of participants throughout January 2021 as participants reached their survey limit or dropped out of the study and new participants were recruited. Participants were included for a maximum of 10 survey rounds in the first group of panels (before 9 August) and 8 in the second group (after 8 August) to reduce the burden of participating on individuals. Because different policies were employed by the four nations of the UK, in this paper we restrict analysis to participants who reported living in England.

The survey design is based on the POLYMOD contact survey [8] with additional questions about work and school attendance, household composition, use of public transportation, and a variety of others. Details of the early rounds of the CoMix study including the protocol and survey instrument have been published previously [7] and details of the updated protocol and survey instruments are provided in the supplementary material.

### Reporting of contacts

Contacts that occurred on the day prior to the survey were reported in two ways: individual contacts and group contacts. First, participants were asked to list each contact and their characteristics separately (‘individual contacts’). Second, we asked participants to report the total number of contacts they had at work, school, or other settings for the age groups 0 to 17, 18 to 59, and 60+, both overall and for physical contacts only (‘group contacts’). We define direct contact as anyone who met the participant in person with whom at least a few words were exchanged or physical contact was made. Questions on group contacts were included at the end of the survey, and they were added to surveys from the 14th May 2020 to accommodate individuals – such as those working in patient- or public-facing roles – who could not record details of all individual contacts that they made. Since August 2020, participants were also asked to describe the average time spent with group contacts and any transmission-related precautions they implemented, including distancing, wearing face-coverings, and performing hand hygiene.

### Demographic information

The survey captures information about participant demographics, employment status and whether participants attended work (or school/university for participants <18 years old and self-reported students) in the previous week and on the day they recorded contacts.

### Risk perception, status, and mitigation

Participants were asked questions about their performance of risk mitigating activities and asked to respond to statements regarding their perception of risk. Participants were asked to respond to the statements: i) “I am likely to catch coronavirus”, ii) “I am worried that I might spread coronavirus to someone who is vulnerable”, and iii) “coronavirus would be a serious illness for me” with the likert scale of “Strongly Agree”, “Tend to Agree”, “Neutral”, “Tend to Disagree”, “Strongly Disagree”. Participants self-reported whether or not they considered themselves to be high risk based on definitions given in the survey, which changed between survey versions as government advice changed (see questionnaires for details). Participants were also asked whether they wore a face covering and in which settings, whether they had washed their hands in the three hours prior to the survey, and whether they sanitized their hands in the three hours prior to the survey.

### Analysis time periods

We categorised the dates of contacts in our survey into nine time periods to compare descriptive statistics and calculate contact matrices. The nine time periods were selected to reflect five stringency levels of non-pharmaceutical interventions we defined as lockdown, lockdown with schools open, lockdown easing, relaxed restrictions (school holiday) and relaxed restrictions (schools open) based on guidance released by the UK Government (Table 1) [1–5].

**Table 1.** Study periods for contact matrices in England. Nine time periods reflect five stringency levels of non-pharmaceutical interventions we defined as lockdown, lockdown with schools open, lockdown easing, relaxed restrictions (school holiday) and relaxed restrictions (schools open) that we created based on guidance released by the UK government. Not all dates are included in a study period.

| Number | Study period | Stringency | Date range |
| --- | --- | --- | --- |
| 1 | Lockdown 1 | Lockdown | 23rd March - 3rd June 2020 |
| 2 | Lockdown 1 easing | Easing | 4th June - 29th July 2020 |
| 3 | Reduce restrictions | Relaxed restrictions | 30th July - 3rd Sep 2020 |
| 4 | Schools open | Relaxed restrictions with Schools open | 4th Sept - 26th October 2020 |
| 5 | Lockdown 2 | Lockdown with schools open | 5th November - 2nd December 2020 |
| 6 | Lockdown 2 easing | Easing | 3rd December - 19th December 2020 |
| 7 | Christmas | Relaxed restrictions | 20 December 2020 - 2nd January 2021 |
| 8 | Lockdown 3 | Lockdown | 5th January - 8th March 2021 |
| 9 | Lockdown 3 (with schools open) | Lockdown with schools open | 9th March - 29th March 2021 |

Previously published analyses considered the period between the 2nd of September and the 5th of November 2020 (i.e., when the second Lockdown started), when England was under a range of local and less stringent restrictions, and therefore this time period has not been included as a study period for this paper [6].

### Statistical analysis

#### Descriptive

R version 4.0.5 was used for all analyses and the code and data are available on GitHub (see Availability of data and materials section) [9].

We calculated summary statistics of the age, gender, socio-economic status, household size, and National Health Service (NHS) Region for participants for each analysis time period and survey panel. While parents answer as proxies for children in the study, we describe the designated child as the “participant” where applicable. We calculated the number and percentage of participants that completed one, two to three, four to five, and six or more rounds of the survey by participant characteristics.

#### Mean contacts

We calculated the mean number of contacts and associated confidence intervals with 1000 samples using clustered bootstrapping [10]. Each participant was sampled with replacement and then all observations for selected participants were included in a bootstrapped sample to account for dependency from repeated observations of the same participants. We calculated the mean number of contacts with a moving window over two-week, overlapping intervals to increase the sample size per estimate and to include all participants from simultaneously running panels. While the initial panels were recruited to be representative of the UK population, we used post-stratification weights of the mean by age group and gender (if available) to address bias introduced by differences between each sample and the UK population [11]. We report contacts by age groups for preschool aged children (<5 years old), school-aged children (5 to 17 years old), adults (18 to 59 years old) and the elderly (60+ years old).

Weights were assigned by the age groups 0 to 4, 5 to 11, 12 to 17, 18 to 29, in 10 year age bands from 30 to 69, and 70 years old and over. We used the World Population Prospects 2019 standard projections overall and by gender for the 2020 UK population [12]. Estimates of the two-week intervals are presented with the data points aligned to the central time point of each survey round and therefore each data point shown is derived from information up to one week before and after the labelled date. We plotted hospitalised cases of COVID-19 alongside mean contact data by age and setting to illustrate the relationship between mean contacts and cases. We used hospitalisation data from the UK government online coronavirus dashboard [13], which we acquired using the *covidregionaldata* R package [14].

We calculated the mean number of contacts in various settings: home, work and school (all educational establishments, including childcare, nurseries and universities and colleges), and “other” (mostly leisure and social contacts, including shopping). The mean number of contacts was influenced by a few participants who reported very high numbers of contacts (often in a work context) relative to the rest of the panel. The distribution of reported contacts are right-skewed with high variance. The mean number of contacts shown here were calculated by censoring the maximum number of contacts recorded at 50 per individual per day to reduce the variance, meaning we counted any individual who reported more than 50 contacts as if they reported 50 contacts to reduce the weight of individuals reporting high numbers of contacts on the mean. We have found in previous analyses that censoring at 50 contacts most closely reflects changes in contacts relative to changes in R_t_ over time as estimated by the Real-time Assessment of Community Transmission (REACT) study, a large home testing study conducted in the UK with the aim of quantifying COVID-19 transmission and infections [15,16].

We report bootstrapped mean contacts using the method previously described by responses to questions about reported risk and risk perception, and by employment and income categories. For likert-style questions we group participant responses of “Tend to Agree”, and “Strongly Agree” into one category of “Agree”, and we group the responses of “Tend to Disagree”, and “Strongly Disagree” into one category of “Disagree”. Only adult participants are included in these analyses. For contacts by employment, we only include participants who recorded working on the day in which they were reporting contacts.

#### Study periods

We calculated relative differences in mean contacts between study periods using an individual-level generalised additive model (GAM) [17,18]. We assumed reported contacts followed a negative binomial distribution, modelled using a log link function, with a random effect for participants by age group (0 to 4, 5 to 17, 18 to 59, and 60 years and over) with post-stratification weights for age and gender (when available) based on the UK population.

#### Face coverings

We present the bootstrapped 95% confidence intervals of the proportions of participants who reported wearing a face covering in any setting for all participants and separately for only those participants who reported contacts outside the household on the day of the survey.

#### Contact matrices

We constructed age-stratified contact matrices for nine age groups (0 to 4, 5 to 11, 12 to 17, 18 to 29, 30 to 39, 40 to 49, 50 to 59, 60 to 69, and 70+ years old). For child participants and contacts, we did not record exact ages and therefore sampled from the reported age-group with a weighting consistent with the age distribution of contacts for the participants’ own age group, according to the POLYMOD survey methods [8]. We fitted a negative binomial model censored to 50 per matrix cell, due to dispersion of the reported number of contacts, to calculate mean contacts between each participant and contact age groups. To find the population normalised reciprocal contact matrix, we first multiplied the columns of the matrix by the mean-normalised proportion of the UK population in each age-group [8,19]. Then we took the cross-diagonal mean of each element of the contact matrix. Finally, we divided the resulting symmetrical matrix by the population mean-normalised proportion of the UK population in each age-group.

We used this approach to construct a contact matrix for each of the analysis periods by filtering the contact data by date. For each time period (table 1) we calculated the dominant eigenvalue of the infectiousness and susceptibility corrected contact matrix (**C**_*SI*_), calculated from the measured contact matrix **C**_*t*_ and assumed age-dependent relative susceptibility and infectiousness vectors **s** and **i**:

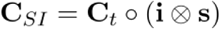

We calculated the relative difference in the basic reproduction number under controlled conditions to Lockdown 1 by taking the ratio of the dominant eigenvalue of the effective contact matrices from the period in question and the dominant eigenvalue from Lockdown 1 [7,20,21]. The value of R_c_, the basic reproduction number under controlled conditions, is defined as the expected number of secondary cases resulting from an initial infection in a completely susceptible population adjusted to changing conditions, in this case the change in social contacts over time. This value does not account for immunity in the population, and will therefore be higher than the actual reproduction number at a given time.

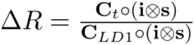

We applied two assumptions of age-dependent susceptibility and infectiousness. First, we assumed that all age-groups are equally infectious and susceptible. Second, we applied a weight for relative susceptibility and infectiousness by age as estimated by Davies et. al.[22] (Supplementary information).

## Results

### Participants characteristics

Overall, we recorded 101,350 observations from 19,914 participants who reported 466,710 contacts over 53 weeks (23rd March 2020 to 29th March 2021). About a quarter of the participants (n=4,574) were proxy respondents (i.e. the survey was completed by parents on behalf of children), and 15,340 were adults.The median number of responses per participant was 6 (min-max 1-9) with 20.6% (4,098) responding only once.

The sample consisted of 8,714 (52.8%) females and 7,790 (47.2%) males. Participants were assigned social grade based on occupation by the Ipsos-MORI company (see map in supplementary materials), which categorised 11,743 (63.1%) participants in social grades A, B, or C1 and 6,880 (36.9%) in C2, D or E. The NHS England region with the most participants was the Midlands with 4,029 (20.2%) participants and the North West had the fewest with 1,931 (9.7%). The characteristics of the participants were consistent over the different analysis periods, with slight variations over the course of the study, particularly in gender balance and household size (Table 2). For instance, around 14% of the participants lived in a single person household in the initial recruitment round versus around 16 to 17% for later recruitment periods.

**Table 2.** Participant characteristics. The number and percentage of participants surveyed during the first two panels (Initial recruitment), the beginning of the next two panels (Second recruitment) and the period since the end of the Christmas study period (New year), as most of the sample was refreshed by this point. Number of participants presented overall and by sample type, age, gender, household size, social group, and NHS region. Participants are counted once per study period but may have participated in several waves within a study period. Adult and child samples were recruited separately, and percentages of age groups were calculated by sample type. The “Other” gender category includes participants who do not describe themselves as either male or female and those who declined to answer. *Some parent participants may have incorrectly completed this question. We have included the observation in this dataset and record the ages as “unknown”.

| Group | Value | Initial recruitment | Second recruitment | New year |
| --- | --- | --- | --- | --- |
| Dates | Start | 23 Mar 20 | 09 Aug 20 | 02 Jan 21 |
|  | End | 08 Aug 20 | 01 Jan 21 | 29 Mar 21 |
| All | - | 5080 | 13087 | 8455 |
| Adult | - | 3815 | 10230 | 6389 |
| Child | - | 1265 | 2857 | 2066 |
| Age Group (Children) | 0-4 | 190 (15.3%) | 451 (16.3%) | 311 (15.8%) |
|  | 5-11 | 469 (37.9%) | 1076 (38.8%) | 749 (38.0%) |
|  | 12-17 | 580 (46.8%) | 1245 (44.9%) | 909 (46.2%) |
|  | Unknown age* | 26 | 85 | 97 |
| Age Group (Adults) | 18-29 | 556 (14.6%) | 1872 (18.3%) | 1070 (16.7%) |
|  | 30-39 | 602 (15.8%) | 1786 (17.5%) | 1167 (18.3%) |
|  | 40-49 | 653 (17.1%) | 1624 (15.9%) | 1069 (16.7%) |
|  | 50-59 | 722 (18.9%) | 1890 (18.5%) | 1183 (18.5%) |
|  | 60-69 | 708 (18.6%) | 1826 (17.8%) | 1147 (18.0%) |
|  | 70+ | 574 (15.0%) | 1232 (12.0%) | 753 (11.8%) |
| Gender | Female | 2580 (50.8%) | 6864 (52.4%) | 4475 (52.9%) |
|  | Male | 2477 (48.8%) | 6162 (47.1%) | 3937 (46.6%) |
|  | Other | 23 | 61 | 43 |
| Household Size | 1 | 698 (13.7%) | 2276 (17.4%) | 1398 (16.5%) |
|  | 2 | 1357 (26.7%) | 4604 (35.2%) | 2900 (34.3%) |
|  | 3-5 | 2769 (54.5%) | 5894 (45.0%) | 3968 (46.9%) |
|  | 6+ | 256 (5.0%) | 313 (2.4%) | 189 (2.2%) |
| Social Group | A - Upper middle class | 247 (4.9%) | 651 (5.0%) | 420 (5.0%) |
|  | B - Middle class | 1278 (25.2%) | 3423 (26.2%) | 2305 (27.3%) |
|  | C1 - Lower middle class | 1535 (30.2%) | 4357 (33.3%) | 2816 (33.3%) |
|  | C2 - Skilled working class | 967 (19.0%) | 1979 (15.1%) | 1297 (15.3%) |
|  | D - Working class | 716 (14.1%) | 1870 (14.3%) | 1141 (13.5%) |
|  | E - Lower level of subsistence | 337 (6.6%) | 807 (6.2%) | 476 (5.6%) |
| NHS Region | East of England | 552 (10.9%) | 1511 (11.5%) | 1009 (11.9%) |
|  | Greater London | 774 (15.2%) | 2001 (15.3%) | 1298 (15.4%) |
|  | Midlands | 1001 (19.7%) | 2688 (20.5%) | 1751 (20.7%) |
|  | North East and Yorkshire | 728 (14.3%) | 2046 (15.6%) | 1315 (15.6%) |
|  | North West | 630 (12.4%) | 1093 (8.4%) | 753 (8.9%) |
|  | South East | 802 (15.8%) | 2263 (17.3%) | 1438 (17.0%) |
|  | South West | 593 (11.7%) | 1485 (11.3%) | 891 (10.5%) |

While participants were recruited to fill quotas by age and gender, participation varies by wave. 32.0% of participants 18 to 29 completed six or more rounds of the survey, while 27.9% completed only one round (Table S1). 60 to 69 year olds had the highest percentage of participants complete six or more rounds at 64.8% and the lowest percentage of participants completing only one round at 10.0%. In children’s panels 36.6% to 38.7% of participants in the child’s age group completed six or more rounds and 18.9% to 22.5% completed only one round, not including those with an unknown age group (Table S2).

### Mean contacts, risk perception, and face coverings

Overall, mean daily contacts for working-aged adults (18 to 59 years) recorded over the study period varied from 2.39 (95% CI 2.20-2.60) during periods of lockdown to 4.93 (95% CI 4.65-5.19) during the summer of 2020, when many restrictions were relaxed (supplementary table 3). Contacts for older adults (60+ years) were consistently lower throughout the study period ranging from 1.55 (95% CI 1.42-1.69) to 3.09 (95% CI 2.82-3.39) contacts per person per day. Mean recorded contacts for school-age children were more variable, between 2.87 (95% CI 1.59-4.74) contacts per day for 0 to 4 year olds during lockdown when their schools were fully or partially closed to 15.11 (95% CI 13.87-16.41) contacts per day for 5 to 17 year olds in September 2020 when schools were open (Figure 1B). Baseline surveys, conducted before the COVID-19 parndemic give an indication of normal levels of contact. The POLYMOD study in 2005/06 reported a mean of 11.7 (standard deviation (sd) 7.7) contacts per participant per day, with the highest mean contacts per day recorded by children aged 10 to 14 (18.2, sd 12.2) while the lowest were in adults over 70 years old (6.9, sd 5.8 contacts per day). The more recent BBC Pandemic social contact study in 2017/18 had similar results, with a mean of 10.5 contacts overall [19].

Following the lifting of Lockdown 1 from late May to early July 2020, recorded contacts remained low until August 2020 (Figure 1B). Contact patterns rebounded much more quickly after the second lockdown in December 2020, despite the continuing imposition of restrictions (a tiered system of restrictions was in place in England, which was strengthened after the second lockdown). Reported contacts were very low during the Christmas period, with a modest easing of restrictions over the holiday period in some parts of England and tighter restrictions in others. Finally, adult contact rates remained low during the third lockdown, with substantial restrictions remaining in place through the end of the study period in March 2021. The patterns of schools opening and closing was the main determinant of children’s contacts (Figure 1B and 1C).

#### Contacts by setting

For adults, contacts made at home mostly reflected household size (Figure S1) and were consistently below a mean of two contacts per day over the study period, with little change in reported contacts across each of the analysis time periods (Figure 1C). Work and other contacts followed a similar pattern to adults: staying low but steadily increasing towards the end of the Lockdown 1, increasing in August 2020, decreasing slightly and then returning to levels similar to the Lockdown 1 during the Lockdown 2 in November, and then reducing again over Christmas and throughout Lockdown 3.

During the first Lockdown schools were closed to all except vulnerable children and the children of essential workers, and recorded childrens’ contact rates were very low (Figure 1B and 1C). From early June 2020 until the third week of July 2020 (when schools were closed for the summer vacation) there was a limited reopening of schools, but most parents reported that their children continued to be educated from home. Average recorded contact patterns amongst children remained very low during this period (Figure 1B and 1C). When schools re-opened fully in September 2020, the number of contacts rapidly increased for both school-aged (5 to 17) and preschool-aged children (0 to 4), though the increase in contacts in the latter age group was smaller. Children’s contacts declined significantly during the “half-term” vacation at the end of October 2020 but remained high during the second national lockdown (November 2020) as schools remained open. Schools were closed for the Christmas period, remained closed during the third national lockdown, and reopened on 8th March 2021. However preschools were the first educational setting to reopen during the relaxation of the first lockdown and were not closed during the third lockdown. The contact patterns of 0-4 year olds reflect this, with mean rates of contact for this age group being higher than other children during the periods when preschools were open but primary and secondary schools were closed.

#### Contacts by study period

Contacts remained at similar levels to Lockdown 1 (the reference period) through Lockdown 1 easing for all age groups until the Reduced restrictions study period (Figure 2; Table 3). The relative difference was highest for adults over the 12 month study period during the period of Reduced restrictions with the relative difference of 1.59 (95% CI 1.54 to 1.64) for adults ages 18 to 59 years and 1.51 (95% CI 1.45 to 1.57) for adults ages over 60 years. For children, the relative difference was highest while schools were open during the Schools open, Lockdown 2, Lockdown 2 easing, and Lockdown 3 with schools open study periods, ranging from 1.74 (95% CI 1.54 to 1.97) for ages 0 to 4 to 3.03 (95% CI 2.82 to 3.26) for ages 5 to 17.

**Figure 2.**
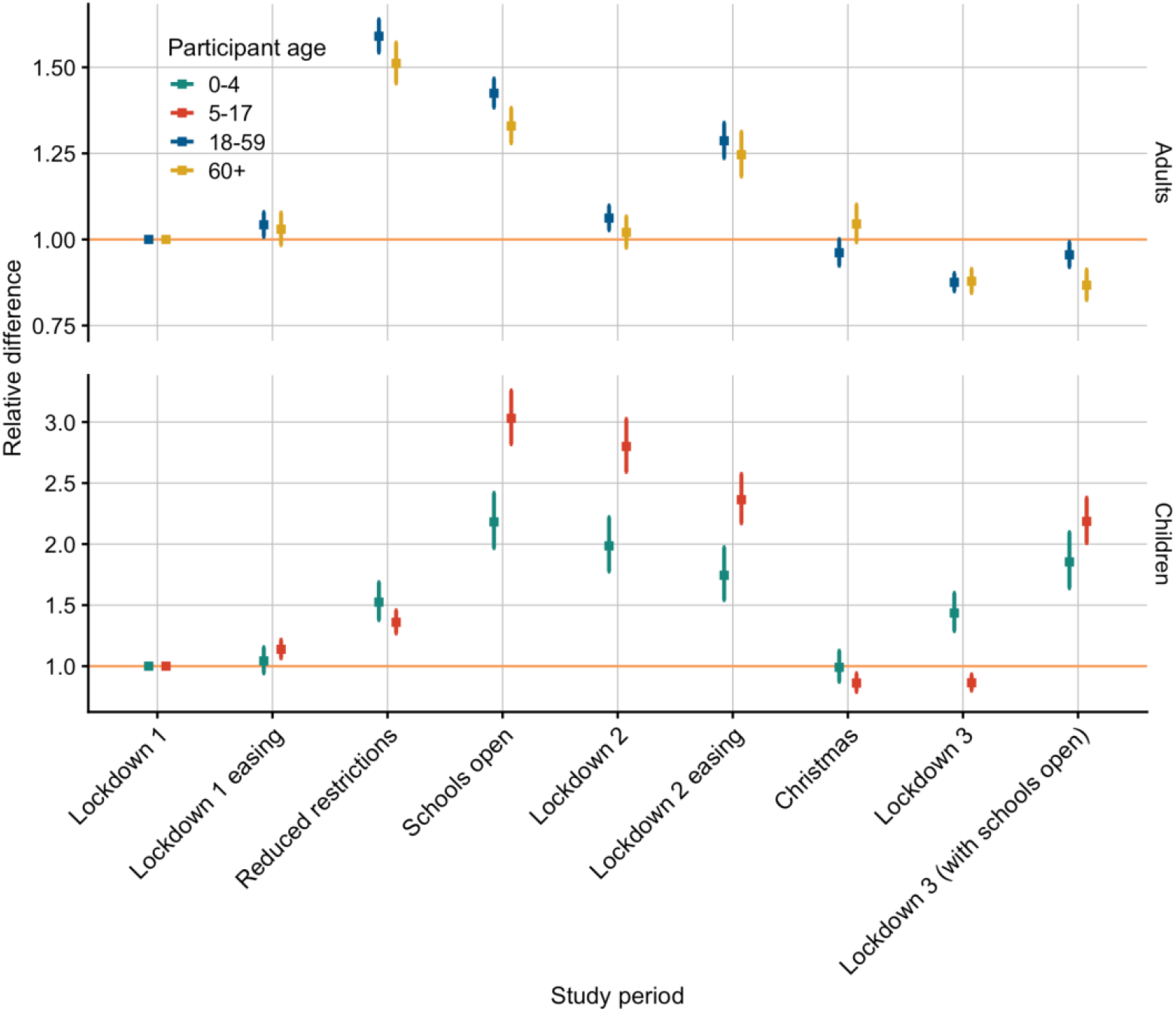
Relative difference in mean contacts by study period and age group with 95% confidence intervals. Relative differences calculated using a generalised additive model with Lockdown 1 as the reference period for each age group adjusted to the UK population by age and gender (when available) for the age groups 0 to 4, 5 to 17, 18 to 59, and over 60 years old. Note the facets have different scales on the y-axes.

**Table 3.** Relative difference in mean contacts by study period with 95% confidence intervals. Relative differences calculated using a generalised additive model with Lockdown 1 as the reference period for each age group adjusted to the UK population by age and gender (when available) for the age groups 0 to 4, 5 to 17, 18 to 59, and over 60 years old.

| Number | Study period | Relative difference in contacts (95% CI) |  |  |  |
| --- | --- | --- | --- | --- | --- |
|  |  | Ages 0-4 years | Ages 5-17 years | Ages 18-59 years | Ages 60+ years |
| 1 | Lockdown 1 | ref | ref | ref | ref |
| 2 | Lockdown 1 easing | 1.04 (0.94 to 1.16) | 1.14 (1.06 to 1.22) | 1.04 (1.01 to 1.08) | 1.03 (0.98 to 1.08) |
| 3 | Reduced restrictions | 1.52 (1.38 to 1.69) | 1.36 (1.27 to 1.46) | 1.59 (1.54 to 1.64) | 1.51 (1.45 to 1.57) |
| 4 | Schools open | 2.18 (1.97 to 2.42) | 3.03 (2.82 to 3.26) | 1.42 (1.38 to 1.47) | 1.33 (1.28 to 1.38) |
| 5 | Lockdown 2 | 1.99 (1.78 to 2.22) | 2.80 (2.59 to 3.02) | 1.06 (1.03 to 1.10) | 1.02 (0.98 to 1.07) |
| 6 | Lockdown 2 easing | 1.74 (1.54 to 1.97) | 2.36 (2.17 to 2.57) | 1.29 (1.24 to 1.34) | 1.25 (1.18 to 1.31) |
| 7 | Christmas | 0.99 (0.87 to 1.13) | 0.86 (0.79 to 0.94) | 0.96 (0.92 to 1.00) | 1.05 (0.99 to 1.10) |
| 8 | Lockdown 3 | 1.44 (1.29 to 1.60) | 0.86 (0.80 to 0.93) | 0.88 (0.85 to 0.90) | 0.88 (0.84 to 0.91) |
| 9 | Lockdown 3 (with schools open) | 1.85 (1.64 to 2.10) | 2.19 (2.01 to 2.38) | 0.96 (0.92 to 0.99) | 0.87 (0.82 to 0.91) |

#### Precautionary behaviours and risk perception

The majority (around 50%) of participants answered “Neutral” to a statement indicating that they were likely to catch coronavirus and this remained fairly consistent over the course of the study (Figure S1), amongst all adult age groups. Survey participants who agreed with a statement that they were likely to catch coronavirus recorded higher mean contacts (Figure 3A), especially in August 2020 and during the period following the second lockdown. Mean contacts for those who disagreed or were neutral were very similar.

**Figure 3.**
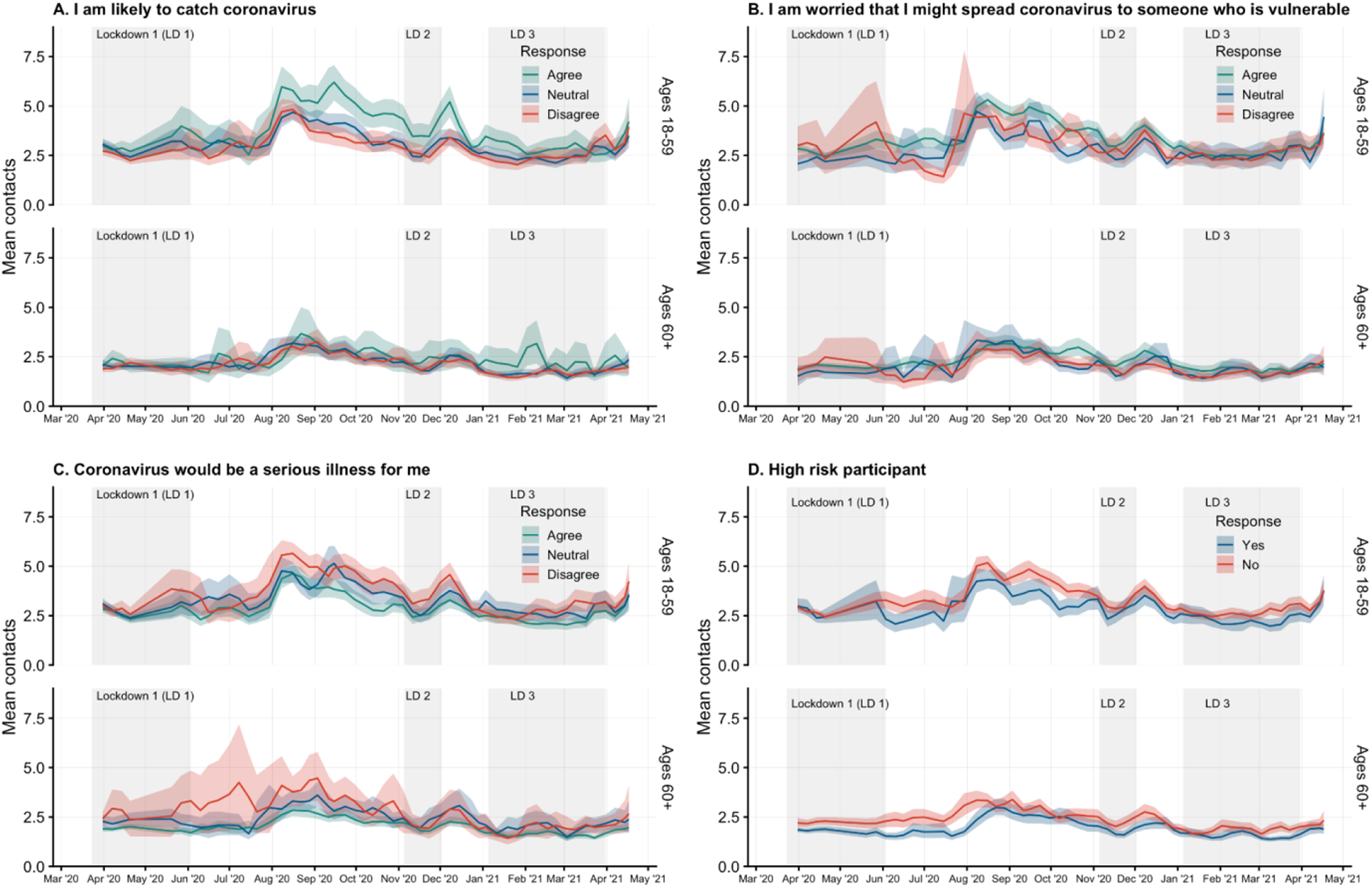
Mean contacts by risk perception or risk category by adult age groups of 18 to 59 and 60 or more years with 95% confidence interval of bootstrapped mean weighted by age, gender, and weekday. Participants answered a series of questions about their risk perception with likert scale response options. Answers of “Strongly agree” and “Somewhat agree” were combined into a category of “Agree, as were answers of “Strongly disagree” and “Somewhat disagree” to “ Disagree”. **A)** Answers to the statement “I am likely to catch coronavirus”; **B)** Answers to the statement “ I am worried I might spread coronavirus to someone who is vulnerable”; **C)** Answers to the statement “Coronavirus would be a serious illness for me”‘; and **D)** Participant reported they were an individual at high risk for complications as defined in the questionnaire.

Participants who agreed with a statement indicating that they were worried that they might spread coronavirus to others generally had slightly higher mean contacts between the first and second lockdowns than those who disagreed with the same statement (Figure 3B). During all three lockdowns, the mean contact confidence intervals overlap for participants in all three categories (Agree, Neutral, Disagree) responding to this question.

Survey participants aged 18 to 59 years who disagreed that coronavirus would be serious for them reported slightly higher contacts than those who agreed with the statement, while participants over 60 years of age who disagreed were few in number and reported a wide range of contact behaviours (Figure 3C). Participants who were not high risk generally reported more contacts on average than those who were high risk in both age groups, especially during periods outside of lockdown and towards the end of the third lockdown, with the differences being more pronounced in the over 60 age group (Figure 3D).

In terms of protective behaviour, the reported use of facemasks at least once on the previous day was low (∼12% for 18–59-year-olds, ∼3% for 60+ years olds) at the end of March 2020 in participants who reported contacts outside of the household (Figure 4). The proportion who self-reported wearing masks increased gradually for both age groups through June 2020, and a sharp increase in mask use was reported in late July and early August 2020, shortly after mask-wearing became mandatory for entering shops on 24th July 2020 [23]. From the 1st of August through 26 March 2021, mask wearing ranged between 73% and 86% for adults 18 to 59 and between 70% and 84% for adults over 60 amongst participants with contacts outside the home.

**Figure 4.**
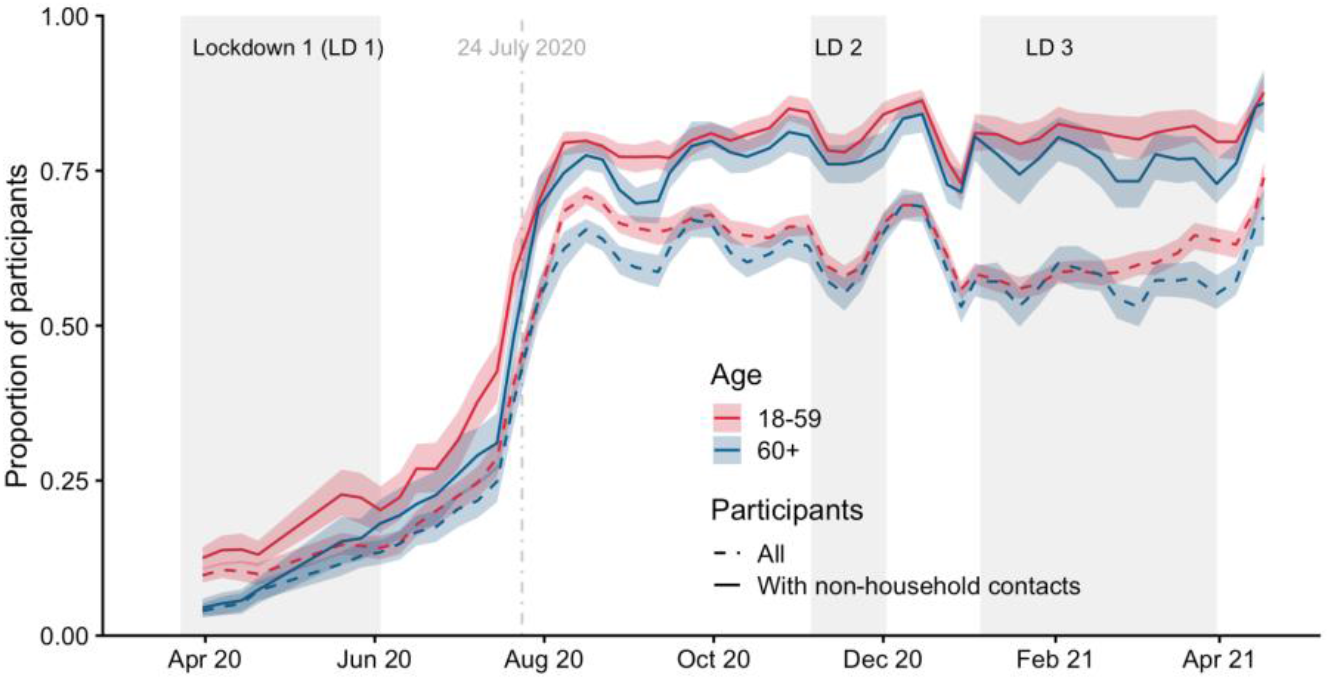
Proportion of adult participants who report wearing a mask by age category with 95% confidence interval of bootstrapped proportion. Proportions plotted for all participants and for participants who reported any non-household contacts, with the start date of face covering mandates in some settings indicated on 24^th^ July 2020.

#### Employment and income

Participants who were employed part-time consistently reported more contacts on average than full-time or self-employed participants with a wider range of contacts, and full-time workers reported contact means similar or slightly higher than self-employed workers in between lockdowns (Figure 5A).

**Figure 5.**
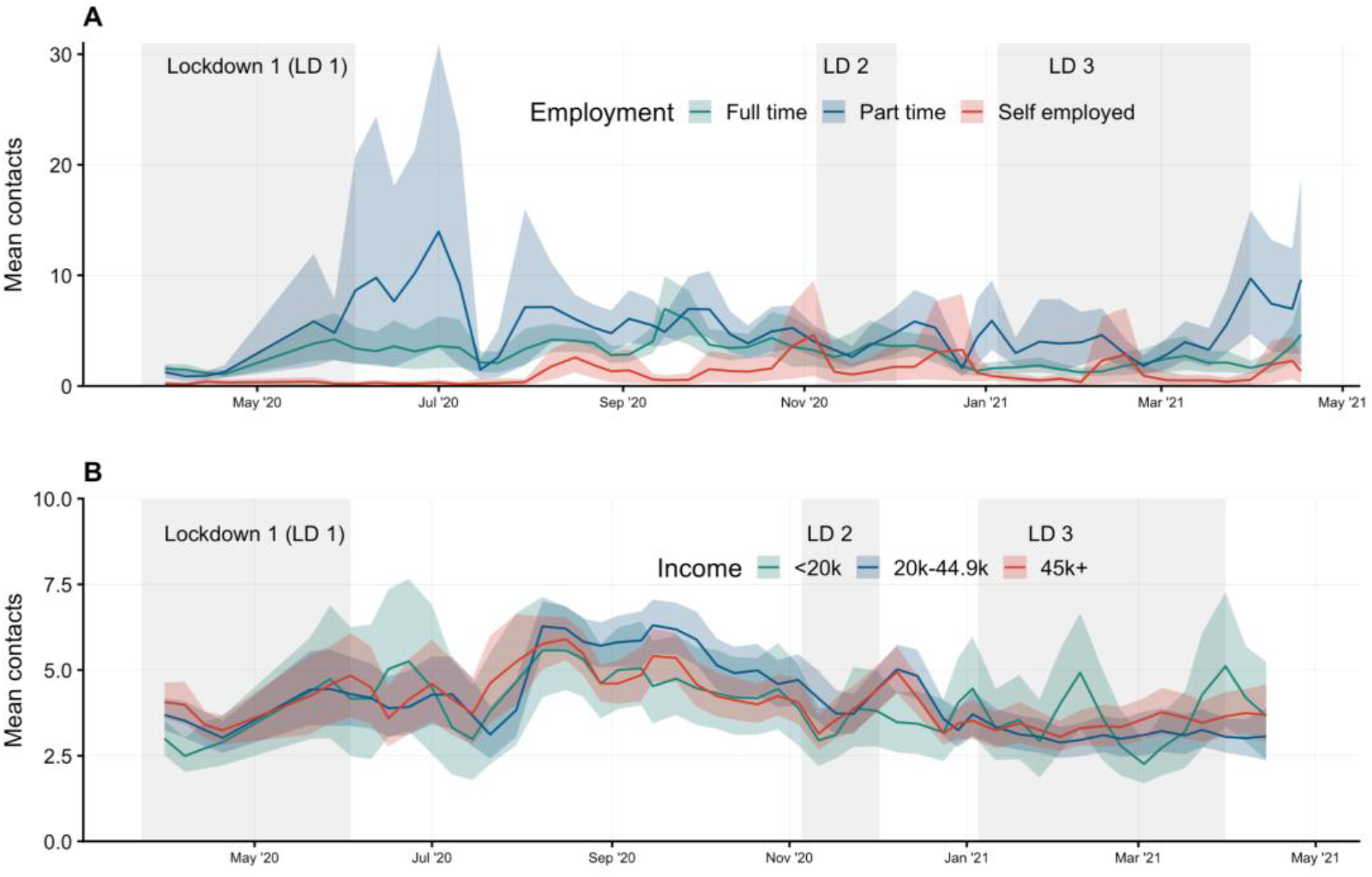
Mean contacts by employment and income status. Mean contacts of participants who worked on the previous day and their workplace was open on the previous day weighted by age, gender, and weekday. **A)** By employment type: full-time, part-time, or self-employed; **B)** By annual income level: less than £20k, £20k to £44.9k, and over £45k.

Mean contacts for adult participants over 18 years of age grouped by annual income levels (less than £20k, £20k to £44.9k, and over £45k), were similar, and follow consistent patterns of decreasing during lockdowns and increasing slightly between the first and second lockdown (Figure 5B).

#### Contact matrices

The contact matrices showed overall higher contacts between all age groups in every period compared to Lockdown 1, with increased clustering around the diagonal matrix elements indicating higher rates of contact between those of similar ages (Figure 6, Supplementary Figure C1, C2 and C3). This resulted in higher estimates of the basic reproduction number under controlled conditions (*R*_*c*_). The periods with the highest *R*_*c*_ were between July and August 2020, which corresponds to lockdown easing and government incentives encouraging the public to dine in restaurants, where contacts particularly increased in 18 to 49 year olds and older adults (>60 year olds). As schools reopened in September 2020 following the summer vacation, an increase in contacts between children increased *R*_*c*_ by a factor of 3.17 (95% CI 3.06 - 3.27) relative to Lockdown 1, or 2.12 (95% CI 2.05 - 2.18) when assuming equal transmissibility in all age groups or assuming age-dependent transmissibility estimates relevant to SARS-CoV-2, respectively.

**Figure 6.**
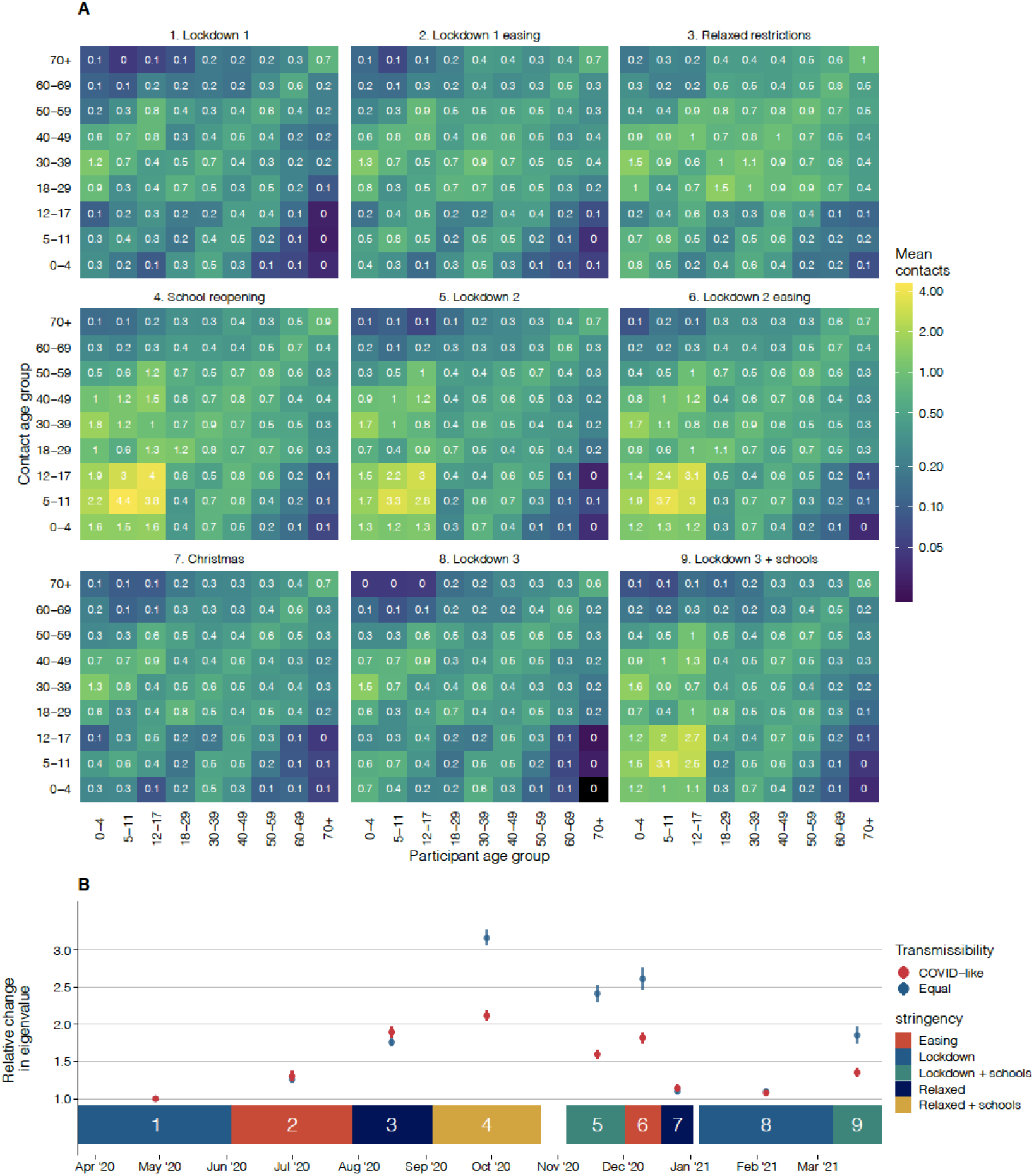
Contact matrices and their dominant eigenvalues for England in each period considered. A) Contact matrices for England across the nine periods (1. Lockdown 1, 2. Lockdown 1 easing, 3. Relaxed restrictions, 4. School reopening, 5. Lockdown 2, 6. Lockdown 2 easing, 7. Christmas, 8. Lockdown 3, 9. Lockdown 3 + schools), B) Points show relative change in R_0 (compared to Lockdown 1) based on the dominant eigenvalues of effective contact matrices calculated for periods 1 - 9, with equal transmissibility in all age groups and age-stratified transmissibility based on Davies et. al. for SARS-CoV-2. Coloured blocks show durations of each period as annotated.

Severe restrictions remained in place over the Christmas holidays (for much of the population) and during Lockdown 3 .However, because early childhood education institutions remained open during Lockdown 3, higher contact rates were reported between <4-year-olds (Supplementary Figure C3). An increase in contacts between school-aged children and a slight increase in contacts amongst adults were found during the periods when schools reopened during Lockdowns 2 and 3 (Supplementary Figure C2, C3). This resulted in an *R*_*c*_ much greater than other lockdown periods, for example during Lockdown 2 we estimated *R*_*c*_ to be 2.42 (2.31 - 2.53, 95%CI) and 1.60 (1.54 - 1.66, 95% CI) times higher than Lockdown 1, for equal and COVID-like transmissibility by age, respectively.

## Discussion

### Summary of findings

We conducted a large, detailed longitudinal social contact survey that has quantified temporal changes in contacts from a representative sample of the English population over the first year of the COVID-19 pandemic. This period (from late March 2020 to late March 2021) encapsulates three periods of national lockdowns interspersed with periods in which fewer restrictions were in place. Mean contact rates have remained low throughout the year - even at the period of minimum restrictions they were only about half of that observed in pre-pandemic surveys, conducted using similar questionnaires [8,19]. These large reductions in contacts have helped reduce the reproduction number, although only during periods of lockdown has the reproduction number been maintained below one [16,24,25].

The survey results suggest that government action was a major factor in the mean number of social contacts, with contact rates dropping markedly during every lockdown. However, it was not the only determinant. Age was clearly associated with contacts, with children reporting the greatest average number of contacts during periods of school opening. Among adults, younger individuals (18 to 30 years) reported the highest mean numbers of contacts throughout the year and the elderly the fewest - a pattern that is consistent with pre-pandemic data, albeit at much lower levels of contact.

In addition, there appeared to be some association with actual or perceived risk: those who were not in a risk group or did not perceive coronavirus as likely to be severe for them reported higher rates of contact. Employment status was also associated with contact patterns, with part-time workers documenting more contacts than others. Income levels were not strongly associated with mean contacts, nor is there any evidence of seasonality in contacts - the observed temporal changes in contacts corresponded to government action and advice. Although easing of restrictions did lead to an increase in contacts, this did not necessarily occur immediately. In particular, the easing of the first lockdown was not associated with a rapid rise in the mean number of contacts until August when the government introduced an incentive scheme to encourage individuals to dine in restaurants, cafes and bars.

The use of face coverings was also strongly associated with changes in government policy. Although the proportion of individuals reporting having worn a face covering increased gradually over time, the rates of mask-wearing increased significantly when it became mandatory for entry into shops on the 24th July 2020.

### Limitations

The survey is conducted online, using a quota-based sample of individuals who have agreed to participate in marketing surveys. This recruitment method is biased towards people with access to the internet and who may be reached by banner ads, email campaigns, and social media advertisements. Participants only received guidance through the text in the questionnaires, and may interpret questions differently. This may be especially evident in the reporting of group contacts. Responses are also subject to recall bias, which may under- or over-estimate contacts depending on the nature of the contacts. Additionally, due to child protection concerns and age-dependent ability to complete the survey, children’s contacts are collected through a parent acting as a proxy for a child, which may lead to inaccurate reporting. Mean contacts are sensitive to a few participants who report many contacts, which we have addressed by assigning all reports of over 50 contacts to 50 contacts. Further research is needed to create standardised methods for analysing highly dispersed contact data, although a standardised approach may not be feasible as it may be context dependent.

### CoMix in context

The CoMix survey contributes to the growing study of social contacts and their implication in disease transmission. Studies prior to the COVID-19 pandemic provide a baseline of contacts in several countries which have been used to project estimates of contacts in other countries [26,27]. Many surveys implemented during the pandemic, conducted in countries throughout the world, provide data on behaviour and social contacts during periods of heightened risk of transmission and with restrictions on social behaviour and have been summarized in a recent review of the literature [28]. CoMix is the largest longitudinal survey in comparison to other surveys in the review and appears to be the only survey to have recorded data every week for at least a year. All surveys in the review reflect fewer social contacts during periods of social restrictions throughout 2020 and 2021. A number of mobility indices such as google and facebook have been made available during the pandemic, which also provide an indication of movement based on monitoring mobile phones. However, these are less direct measures that reflect less epidemiologically-relevant contacts, and although previous work has suggested that google mobility data correlates well with the CoMix data [29] the data is usually shared at aggregate level and therefore is impossible to analyse by factors such as age and working status.

### Conclusion

This study quantifies changes in epidemiologically relevant contact behaviour for one full year of the COVID-19 pandemic in England. Contacts have remained suppressed far below normal levels throughout the year, though changes in contact have occurred following relaxation or tightening of social distancing measures.

The CoMix survey is unique in both length and frequency of the data and in its longitudinal study design, which provides a detailed historical record of social behaviour during the COVID-19 pandemic. Importantly, CoMix contact data is age-stratified for both participants and contacts and can be used to construct social contact matrices for age-stratified modelling. This data can be used to inform future outbreak response and can be applied to transmission of other infectious diseases, particularly for a large scale pandemic.

## Supporting information

Final survey versions for panels A&B, C&D, and E&F.

Recruitment methods and social grades mapped to reported occupations

Original research proposal for the CoMix survey

Bootstrapped mean contacts with 95% confidence interval in all settings

## Data Availability

The code and data used to conduct these analyses are found at: https://github.com/amygimma/comix_uk_summary_analysis
Data will be available to download through the Socrates social contact tool website and Zenodo:
http://www.socialcontactdata.org/socrates-comix/

http://www.socialcontactdata.org/socrates-comix/

## Additional information

## Abbreviations

CI: Confidence interval
GAM: Generalised additive model
IQR: Interquartile range
NHS: National Health Service
ONS UK: Office of National Statistics for the United Kingdom
R_c_: Basic reproduction number under controlled conditions
R_t_: Reproduction number at time t
REACT Study: Real-time Assessment of Community Transmission Study
SD: Standard deviation
UK: United Kingdom

Centre for Mathematical Modelling of Infectious Disease Working Group Authors

The following authors were part of the Centre for Mathematical Modelling of Infectious Disease COVID-19 Working Group. Each contributed in processing, cleaning and interpretation of data, interpreted findings, contributed to the manuscript, and approved the work for publication: Lloyd A C Chapman, Samuel Clifford, Thibaut Jombart, Kathleen O’Reilly, Jiayao Lei, Kaja Abbas, Fabienne Krauer, Stefan Flasche, Alicia Rosello, Gwenan M Knight, Damien C Tully, Katherine E. Atkins, Rachael Pung, Rosalind M Eggo, David Hodgson, Mihaly Koltai, Yalda Jafari, Timothy W Russell, Frank G Sandmann, Oliver Brady, Naomi R Waterlow, Mark Jit, Fiona Yueqian Sun, Carl A B Pearson, William Waites, Emilie Finch, Akira Endo, Graham Medley, Ciara V McCarthy, Adam J Kucharski, Paul Mee, Hamish P Gibbs, Nicholas G. Davies, Billy J Quilty, Sophie R Meakin, C Julian Villabona-Arenas, Nikos I Bosse, Joel Hellewell, Simon R Procter, Yang Liu, Rachel Lowe, Rosanna C Barnard, Sam Abbott, Matthew Quaife, Emily S Nightingale.

## Authors’ contributions

AG, JDM, KLMW, CIJ, WJE conceived of and planned the analysis; AG, JDM, KLMW, and CIJ performed the main analysis with input from PC, KP, PK, GJR, SF and WJE; CIJ, KvZ, and WEJ designed the CoMix contact survey, CIJ, AG, KW, and KvZ cleaned and managed the contact survey data; All authors wrote and reviewed the manuscript. The CMMID COVID-19 Working Group provided discussion and comments.

## Acknowledgements

The authors would like to thank the team at Ipsos-MORI, an international research agency which implemented the surveys, with particular thanks to Sean Doherty, Alexandru Toreanik, Lorena Iovu, Corneliu Caloian, Alina Pancu, Rares Eremia and Kim Brown for their work on coordinating and implementing multiple waves of the survey.

We would also like to thank the EpiPose management team and collaborating researchers, including Niel Hens, Jacco Wallinga, Philippe Beutels, Jantien Backer, James Wambua, Laurens Bogaardt, Veronika Jaeger, and Andre Karch.

## Ethics approval and consent to participate

Participation in this opt-in study was voluntary, and all analyses were carried out on anonymised data. The study and method of informed consent was approved by the ethics committee of the London School of Hygiene & Tropical Medicine Reference number 21795.

## Consent for publication

Not applicable

## Availability of data and materials

The code and data used to conduct these analyses are found at: https://github.com/amygimma/comix_uk_summary_analysis

Data will be available to download through the Socrates social contact tool website and Zenodo: http://www.socialcontactdata.org/socrates-comix/

## Additional Files

**Additional File 1: CoMix Contact Survey Questionnaires – Final Versions**. Final survey versions for panels A&B, C&D, and E&F.

**Additional File 2: CoMix study Proposal**. Original research proposal for the CoMix survey.

**Additional File 3. Ipsos-MORI Survey Recruitment and Social Grades**. Recruitment methods and social grades mapped to reported occupations.

**Additional File 4. Mean Contacts by Age and Date**. Bootstrapped mean contacts with 95% confidence interval in all settings.

## Supplementary Figures and Tables

### Contact matrices

**Figure C1:**
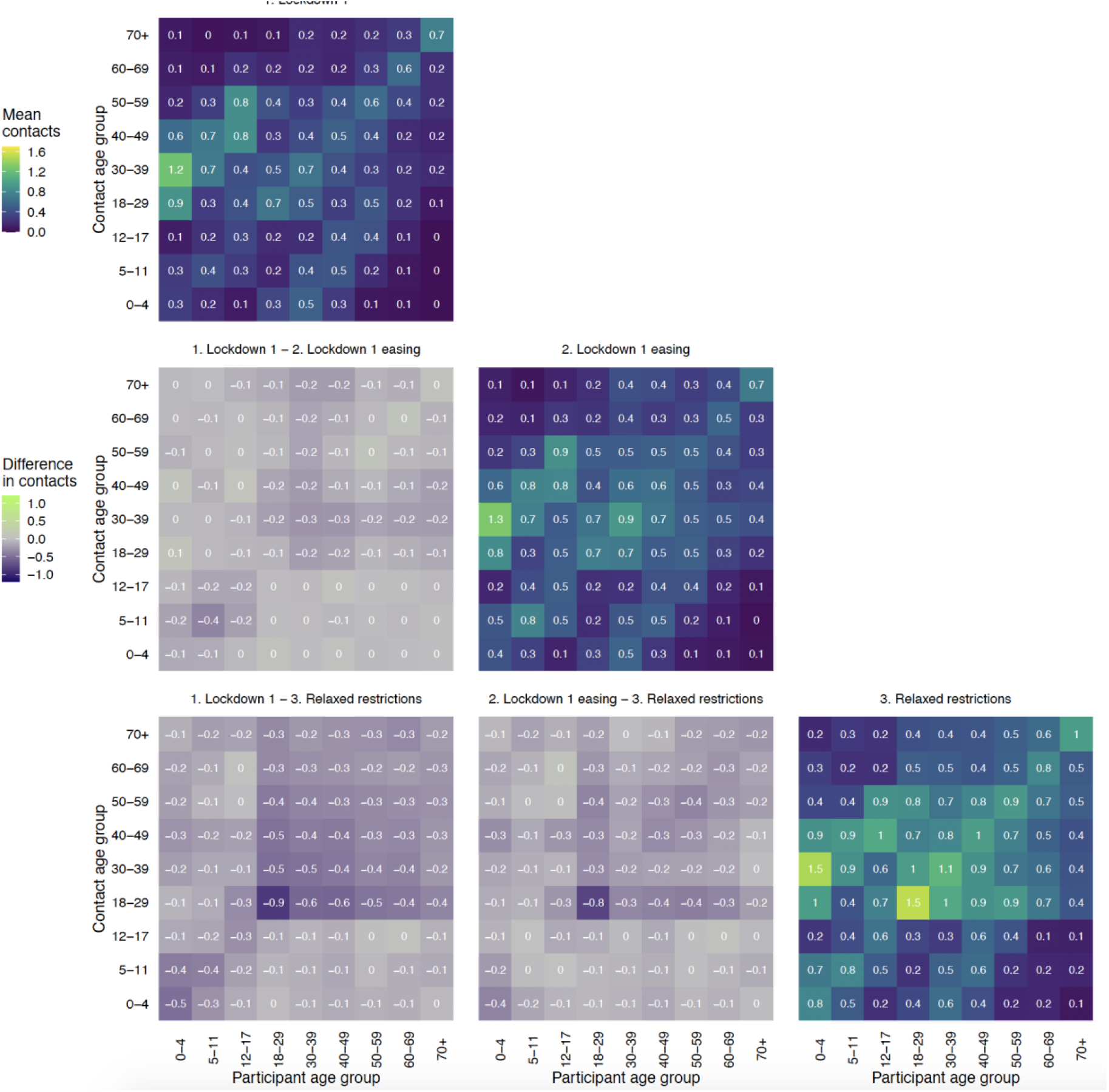
Contact matrices for all contacts in England for Lockdown 1, Lockdown 1 easing and Relaxed restrictions (Diagonal) and the element-wise absolute difference between the matrices (off diagonal). Contacts censored to 50 contacts per participant. Lockdown 1 data from 23rd of March to 3rd of June 2020 Lockdown 3 data from 5th to 18th of January 2021

**Figure C2:**
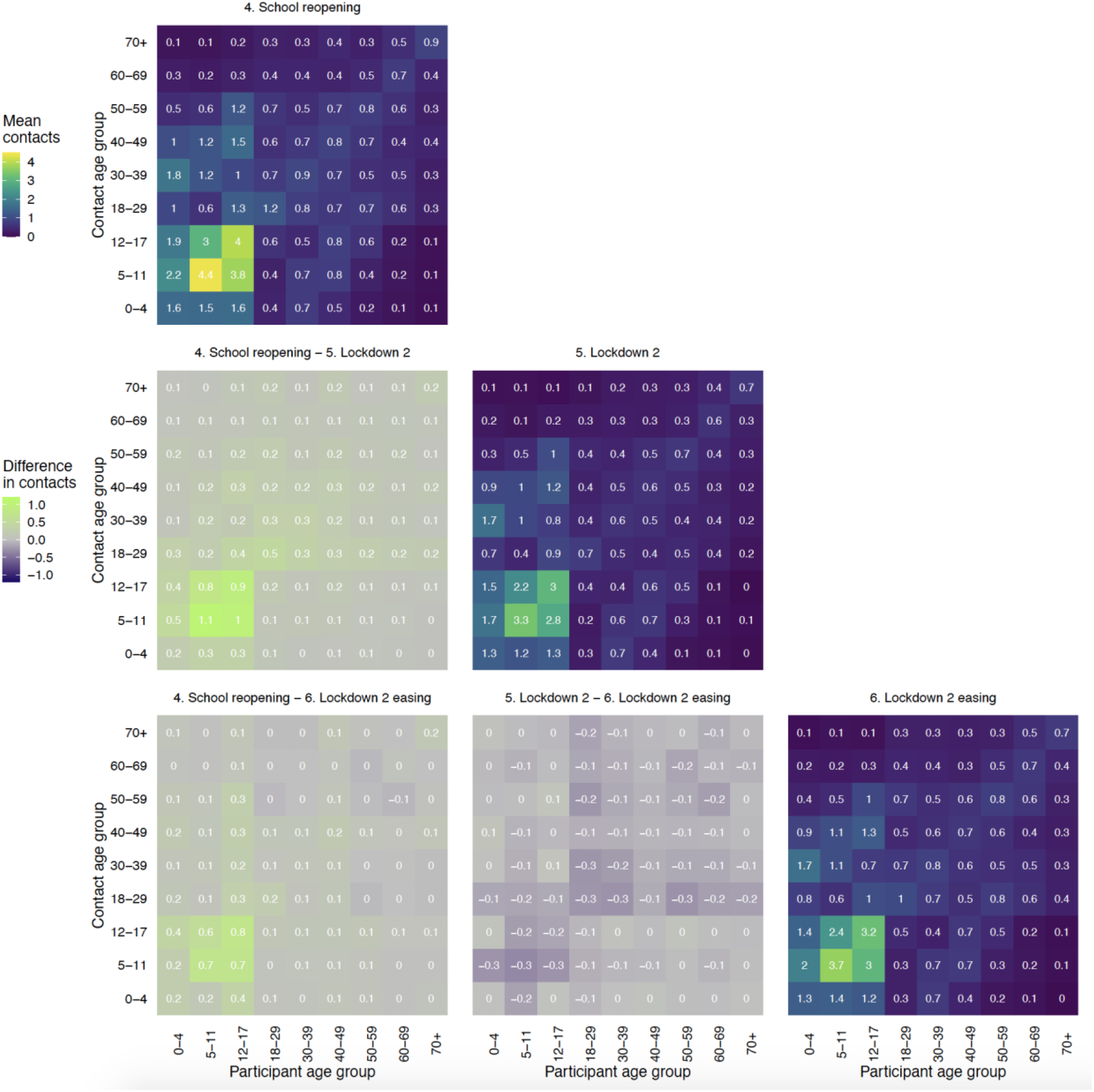
Contact matrices for all contacts in England for Schools reopening, Lockdown 2 and Lockdown 2 easing (Diagonal) and the element-wise absolute difference between the matrices (off diagonal). Contacts censored to 50 contacts per participant. Lockdown 1 data from 23rd of March to 3rd of June 2020 Lockdown 3 data from 5th to 18th of January 2021

**Figure C3:**
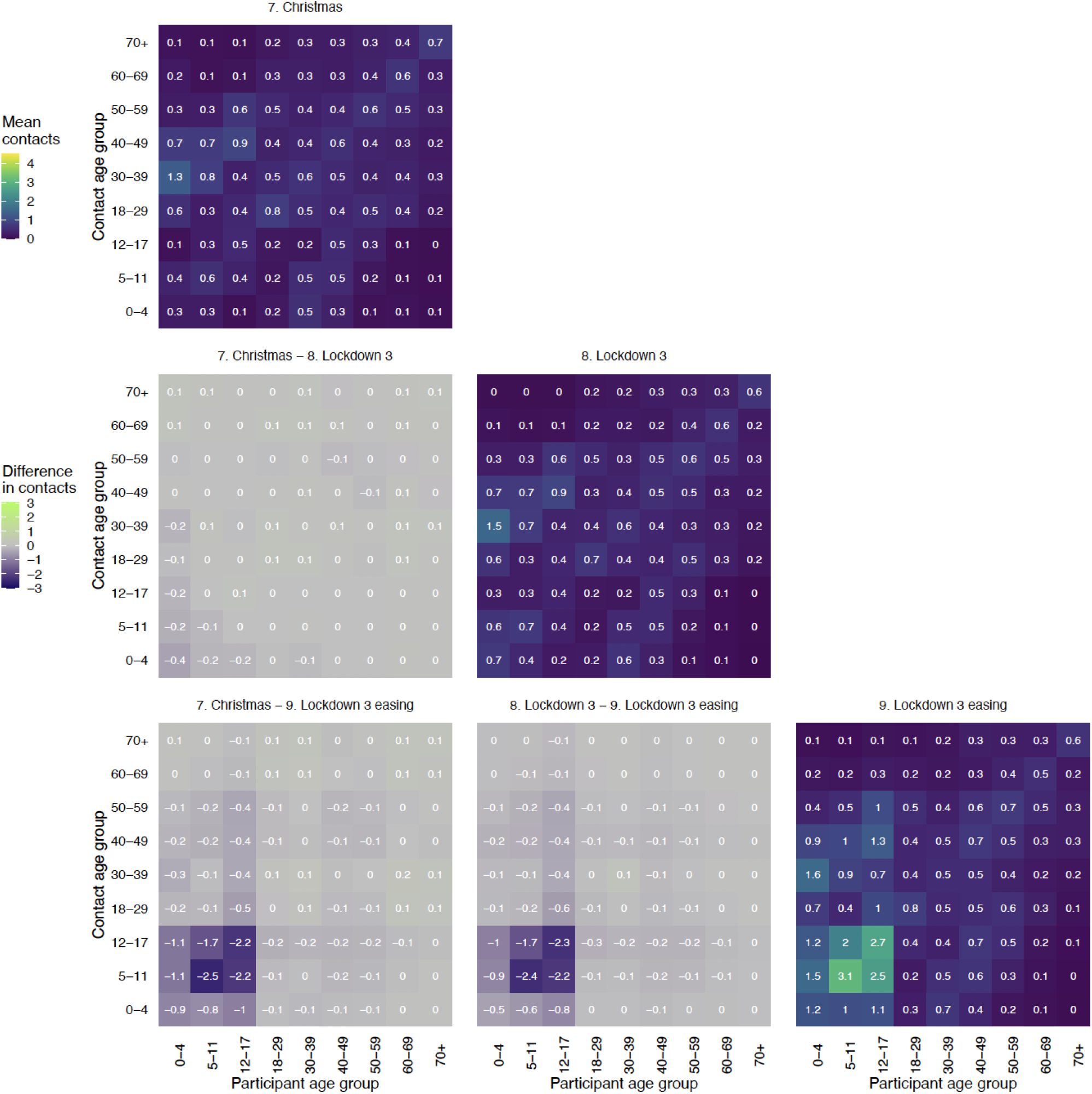
Contact matrices for all contacts in England for Christmas, Lockdown 3 and Lockdown 3 easing (Diagonal) and the element-wise absolute difference between the matrices (off diagonal). Contacts censored to 50 contacts per participant. Lockdown 1 data from 23rd of March to 3rd of June 2020 Lockdown 3 data from 5th to 18th of January 2021

## Additional figures and tables

**Figure S1.**
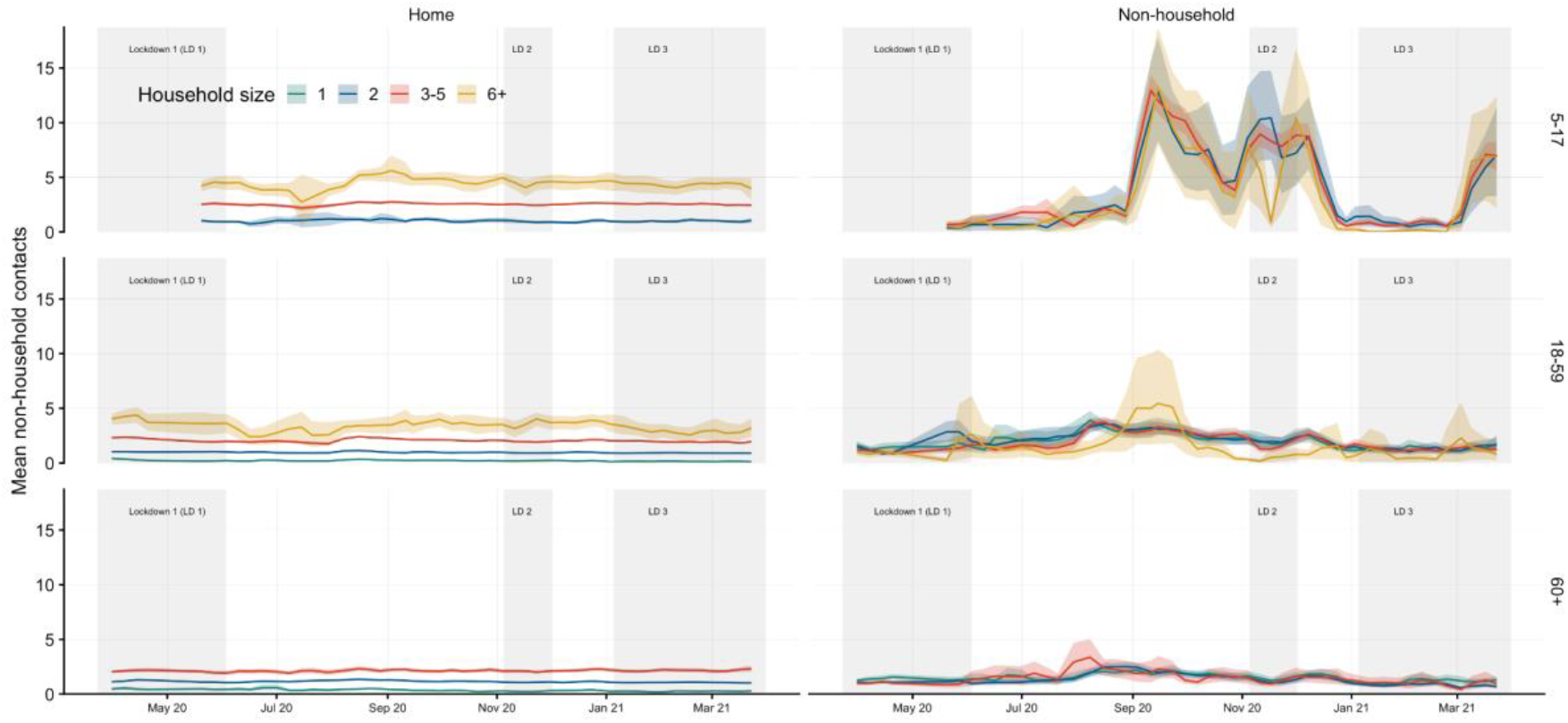
Mean contacts by age group and household size. Means weighted by age, gender, and weekday. Age and household size categories with fewer than 50 participants in total are not shown, and the age category 0 to 4 years old not shown as most participants were in household sizes of 3 to 5. Some time periods have very few participants in households 6 or more and should be interpreted with caution.

**Figure S2.**
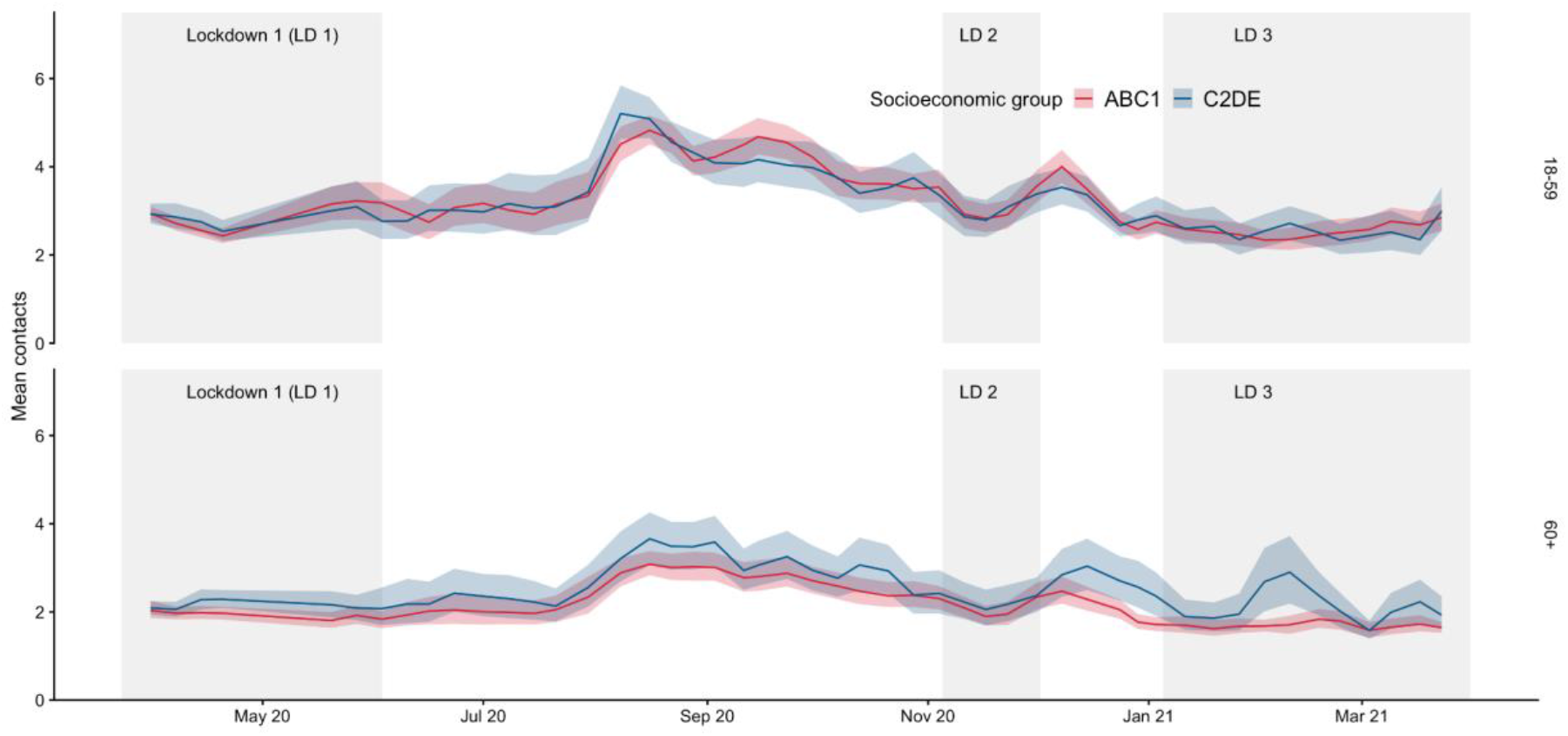
Mean contacts and 95% confidence intervals by age group and socioeconomic groups combined into the groups ABC1 and C2DE. Bootstrapped mean contacts of participants weighted by age, gender, and weekday.

**Figure S3.**
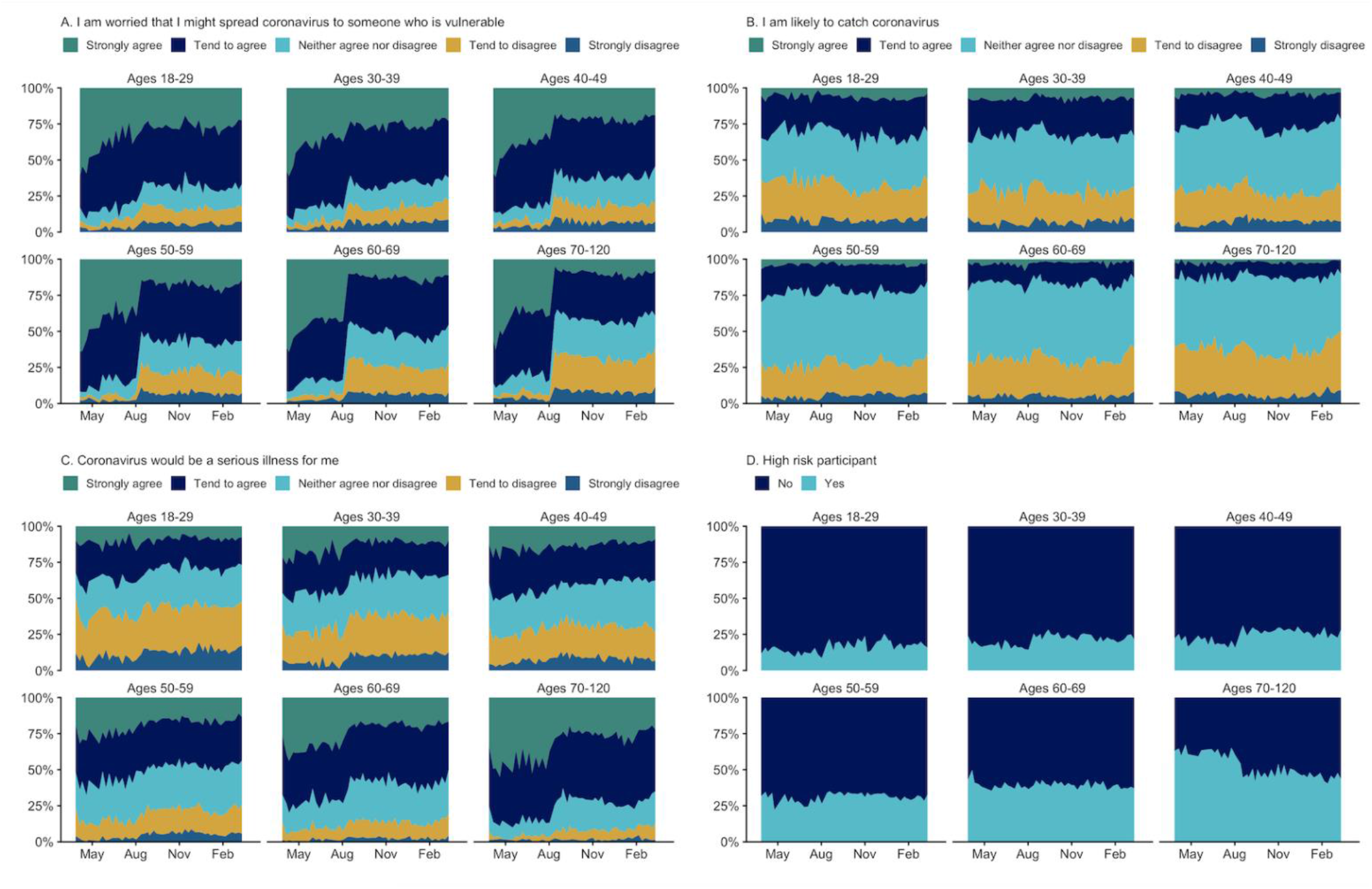
Risk perception by age group over time. The raw proportion of likert scale responses and self-reported risk status amongst adult participants

**Table S1.**
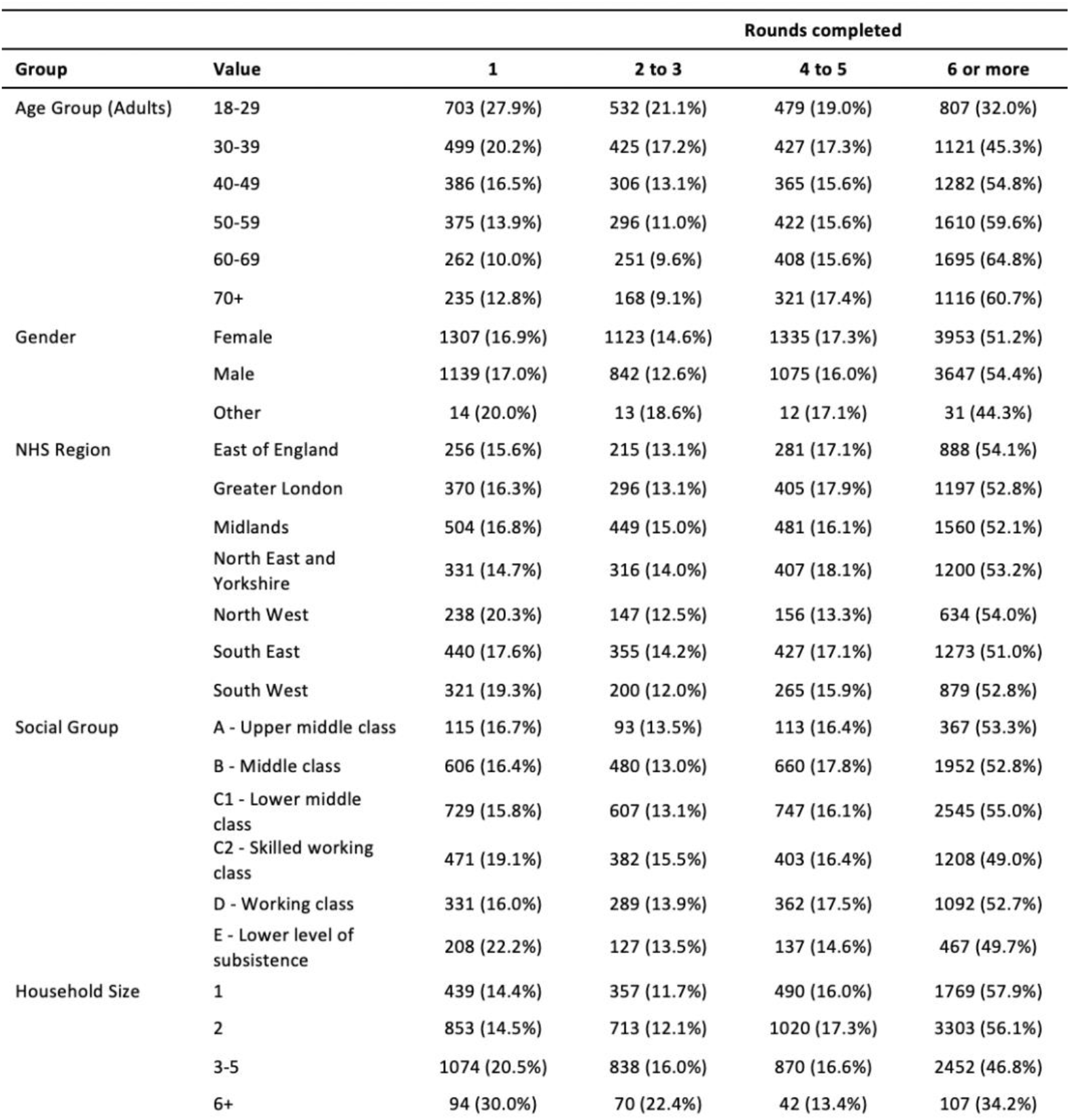
Number and percentage of participants in the adult panel who completed 1 round, 2 to 3 rounds, or 5 or more rounds, stratified by gender, age, country, NHS England region, household size, and day of week.

| Group | Value | Rounds completed |  |  |  |
| --- | --- | --- | --- | --- | --- |
|  |  | 1 | 2 to 3 | 4 to 5 | 6 or more |
| Age Group (Adults) | 18-29 | 703 (27.9%) | 532 (21.1%) | 479 (19.0%) | 807 (32.0%) |
|  | 30-39 | 499 (20.2%) | 425 (17.2%) | 427 (17.3%) | 1121 (45.3%) |
|  | 40-49 | 386 (16.5%) | 306 (13.1%) | 365 (15.6%) | 1282 (54.8%) |
|  | 50-59 | 375 (13.9%) | 296 (11.0%) | 422 (15.6%) | 1610 (59.6%) |
|  | 60-69 | 262 (10.0%) | 251 (9.6%) | 408 (15.6%) | 1695 (64.8%) |
|  | 70+ | 235 (12.8%) | 168 (9.1%) | 321 (17.4%) | 1116 (60.7%) |
| Gender | Female | 1307 (16.9%) | 1123 (14.6%) | 1335 (17.3%) | 3953 (51.2%) |
|  | Male | 1139 (17.0%) | 842 (12.6%) | 1075 (16.0%) | 3647 (54.4%) |
|  | Other | 14 (20.0%) | 13 (18.6%) | 12 (17.1%) | 31 (44.3%) |
| NHS Region | East of England | 256 (15.6%) | 215 (13.1%) | 281 (17.1%) | 888 (54.1%) |
|  | Greater London | 370 (16.3%) | 296 (13.1%) | 405 (17.9%) | 1197 (52.8%) |
|  | Midlands | 504 (16.8%) | 449 (15.0%) | 481 (16.1%) | 1560 (52.1%) |
|  | North East and Yorkshire | 331 (14.7%) | 316 (14.0%) | 407 (18.1%) | 1200 (53.2%) |
|  | North West | 238 (20.3%) | 147 (12.5%) | 156 (13.3%) | 634 (54.0%) |
|  | South East | 440 (17.6%) | 355 (14.2%) | 427 (17.1%) | 1273 (51.0%) |
|  | South West | 321 (19.3%) | 200 (12.0%) | 265 (15.9%) | 879 (52.8%) |
| Social Group | A - Upper middle class | 115 (16.7%) | 93 (13.5%) | 113 (16.4%) | 367 (53.3%) |
|  | B - Middle class | 606 (16.4%) | 480 (13.0%) | 660 (17.8%) | 1952 (52.8%) |
|  | C1 - Lower middle class | 729 (15.8%) | 607 (13.1%) | 747 (16.1%) | 2545 (55.0%) |
|  | C2 - Skilled working class | 471 (19.1%) | 382 (15.5%) | 403 (16.4%) | 1208 (49.0%) |
|  | D - Working class | 331 (16.0%) | 289 (13.9%) | 362 (17.5%) | 1092 (52.7%) |
|  | E - Lower level of subsistence | 208 (22.2%) | 127 (13.5%) | 137 (14.6%) | 467 (49.7%) |
| Household Size | 1 | 439 (14.4%) | 357 (11.7%) | 490 (16.0%) | 1769 (57.9%) |
|  | 2 | 853 (14.5%) | 713 (12.1%) | 1020 (17.3%) | 3303 (56.1%) |
|  | 3-5 | 1074 (20.5%) | 838 (16.0%) | 870 (16.6%) | 2452 (46.8%) |
|  | 6+ | 94 (30.0%) | 70 (22.4%) | 42 (13.4%) | 107 (34.2%) |

**Table S2.** Number and percentage (by number of rounds completed) of parent participants who completed 1 round, 2 to 3 rounds, or 5 or more rounds, overall and stratified by gender, age, country, NHS England region, household size, and day of week. Parents of children report gender by answering the question “As far as you know, which of the following describes how [NAME OF CHILD] thinks of themselves?”, with the options “Male”, “Female”, “In another way”, “Don’t know” and “Prefer not to answer”.

| Group | Value | 1 | 2 to 3 | 4 to 5 | 6 or more |
| --- | --- | --- | --- | --- | --- |
| Age Group<br>(Child) | 0-4 | 134 (20.6%) | 139 (21.4%) | 137 (21.1%) | 240 (36.9%) |
|  | 5-11 | 354 (22.5%) | 321 (20.4%) | 321 (20.4%) | 574 (36.6%) |
|  | 12-17 | 353 (18.9%) | 411 (22.1%) | 378 (20.3%) | 721 (38.7%) |
|  | Unknown age | 23 (20.5%) | 25 (22.3%) | 20 (17.9%) | 44 (39.3%) |
| Gender | Female | 536 (19.9%) | 574 (21.3%) | 527 (19.6%) | 1057 (39.2%) |
|  | Male | 327 (21.9%) | 318 (21.3%) | 327 (21.9%) | 521 (34.9%) |
|  | Other | 1 (12.5%) | 4 (50.0%) | 2 (25.0%) | 1 (12.5%) |
| NHS Region | East of England | 85 (18.1%) | 101 (21.5%) | 97 (20.6%) | 187 (39.8%) |
|  | Greater London | 142 (24.1%) | 119 (20.2%) | 109 (18.5%) | 219 (37.2%) |
|  | Midlands | 158 (19.7%) | 177 (22.1%) | 161 (20.1%) | 305 (38.1%) |
|  | North East and Yorkshire | 116 (19.3%) | 122 (20.3%) | 135 (22.5%) | 227 (37.8%) |
|  | North West | 119 (20.5%) | 121 (20.8%) | 116 (20.0%) | 225 (38.7%) |
|  | South East | 140 (20.4%) | 136 (19.8%) | 150 (21.9%) | 260 (37.9%) |
|  | South West | 104 (22.2%) | 120 (25.6%) | 88 (18.8%) | 156 (33.3%) |
| Social Group | A - Upper middle class | 68 (29.3%) | 37 (15.9%) | 44 (19.0%) | 83 (35.8%) |
|  | B - Middle class | 209 (18.8%) | 226 (20.3%) | 222 (20.0%) | 455 (40.9%) |
|  | C1 - Lower middle class | 247 (17.5%) | 315 (22.3%) | 303 (21.4%) | 548 (38.8%) |
|  | C2 - Skilled working class | 131 (21.9%) | 140 (23.4%) | 122 (20.4%) | 206 (34.4%) |
|  | D - Working class | 147 (24.7%) | 121 (20.3%) | 114 (19.2%) | 213 (35.8%) |
|  | E - Lower level of subsistence | 62 (25.4%) | 57 (23.4%) | 51 (20.9%) | 74 (30.3%) |
| Household Size | 1 | 7 (20.0%) | 7 (20.0%) | 14 (40.0%) | 7 (20.0%) |
|  | 2 | 91 (19.3%) | 108 (22.9%) | 130 (27.5%) | 143 (30.3%) |
|  | 3-5 | 711 (20.3%) | 745 (21.3%) | 678 (19.4%) | 1366 (39.0%) |
|  | 6+ | 55 (29.3%) | 36 (19.1%) | 34 (18.1%) | 63 (33.5%) |

**Table S3.**
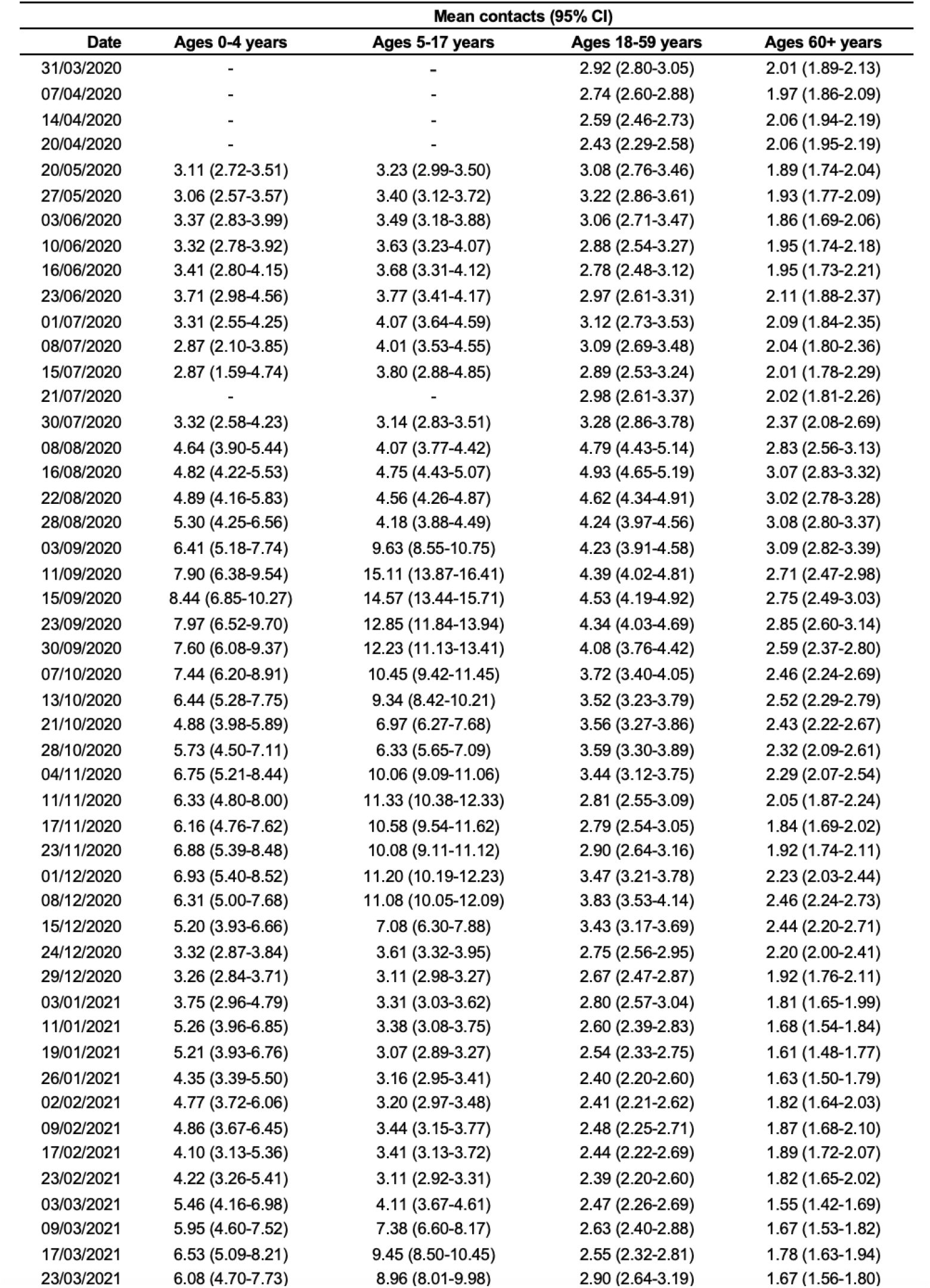
Mean contacts over time by age with 95% confidence interval of bootstrapped mean. Mean reported contacts of participants weighted by age, gender, and weekday.

| Date | Mean contacts (95% CI) |  |  |  |
| --- | --- | --- | --- | --- |
|  | Ages 0-4 years | Ages 5-17 years | Ages 18-59 years | Ages 60+ years |
| 31/03/2020 | - | - | 2.92 (2.80-3.05) | 2.01 (1.89-2.13) |
| 07/04/2020 | - | - | 2.74 (2.60-2.88) | 1.97 (1.86-2.09) |
| 14/04/2020 | - | - | 2.59 (2.46-2.73) | 2.06 (1.94-2.19) |
| 20/04/2020 | - | - | 2.43 (2.29-2.58) | 2.06 (1.95-2.19) |
| 20/05/2020 | 3.11 (2.72-3.51) | 3.23 (2.99-3.50) | 3.08 (2.76-3.46) | 1.89 (1.74-2.04) |
| 27/05/2020 | 3.06 (2.57-3.57) | 3.40 (3.12-3.72) | 3.22 (2.86-3.61) | 1.93 (1.77-2.09) |
| 03/06/2020 | 3.37 (2.83-3.99) | 3.49 (3.18-3.88) | 3.06 (2.71-3.47) | 1.86 (1.69-2.06) |
| 10/06/2020 | 3.32 (2.78-3.92) | 3.63 (3.23-4.07) | 2.88 (2.54-3.27) | 1.95 (1.74-2.18) |
| 16/06/2020 | 3.41 (2.80-4.15) | 3.68 (3.31-4.12) | 2.78 (2.48-3.12) | 1.95 (1.73-2.21) |
| 23/06/2020 | 3.71 (2.98-4.56) | 3.77 (3.41-4.17) | 2.97 (2.61-3.31) | 2.11 (1.88-2.37) |
| 01/07/2020 | 3.31 (2.55-4.25) | 4.07 (3.64-4.59) | 3.12 (2.73-3.53) | 2.09 (1.84-2.35) |
| 08/07/2020 | 2.87 (2.10-3.85) | 4.01 (3.53-4.55) | 3.09 (2.69-3.48) | 2.04 (1.80-2.36) |
| 15/07/2020 | 2.87 (1.59-4.74) | 3.80 (2.88-4.85) | 2.89 (2.53-3.24) | 2.01 (1.78-2.29) |
| 21/07/2020 | - | - | 2.98 (2.61-3.37) | 2.02 (1.81-2.26) |
| 30/07/2020 | 3.32 (2.58-4.23) | 3.14 (2.83-3.51) | 3.28 (2.86-3.78) | 2.37 (2.08-2.69) |
| 08/08/2020 | 4.64 (3.90-5.44) | 4.07 (3.77-4.42) | 4.79 (4.43-5.14) | 2.83 (2.56-3.13) |
| 16/08/2020 | 4.82 (4.22-5.53) | 4.75 (4.43-5.07) | 4.93 (4.65-5.19) | 3.07 (2.83-3.32) |
| 22/08/2020 | 4.89 (4.16-5.83) | 4.56 (4.26-4.87) | 4.62 (4.34-4.91) | 3.02 (2.78-3.28) |
| 28/08/2020 | 5.30 (4.25-6.56) | 4.18 (3.88-4.49) | 4.24 (3.97-4.56) | 3.08 (2.80-3.37) |
| 03/09/2020 | 6.41 (5.18-7.74) | 9.63 (8.55-10.75) | 4.23 (3.91-4.58) | 3.09 (2.82-3.39) |
| 11/09/2020 | 7.90 (6.38-9.54) | 15.11 (13.87-16.41) | 4.39 (4.02-4.81) | 2.71 (2.47-2.98) |
| 15/09/2020 | 8.44 (6.85-10.27) | 14.57 (13.44-15.71) | 4.53 (4.19-4.92) | 2.75 (2.49-3.03) |
| 23/09/2020 | 7.97 (6.52-9.70) | 12.85 (11.84-13.94) | 4.34 (4.03-4.69) | 2.85 (2.60-3.14) |
| 30/09/2020 | 7.60 (6.08-9.37) | 12.23 (11.13-13.41) | 4.08 (3.76-4.42) | 2.59 (2.37-2.80) |
| 07/10/2020 | 7.44 (6.20-8.91) | 10.45 (9.42-11.45) | 3.72 (3.40-4.05) | 2.46 (2.24-2.69) |
| 13/10/2020 | 6.44 (5.28-7.75) | 9.34 (8.42-10.21) | 3.52 (3.23-3.79) | 2.52 (2.29-2.79) |
| 21/10/2020 | 4.88 (3.98-5.89) | 6.97 (6.27-7.68) | 3.56 (3.27-3.86) | 2.43 (2.22-2.67) |
| 28/10/2020 | 5.73 (4.50-7.11) | 6.33 (5.65-7.09) | 3.59 (3.30-3.89) | 2.32 (2.09-2.61) |
| 04/11/2020 | 6.75 (5.21-8.44) | 10.06 (9.09-11.06) | 3.44 (3.12-3.75) | 2.29 (2.07-2.54) |
| 11/11/2020 | 6.33 (4.80-8.00) | 11.33 (10.38-12.33) | 2.81 (2.55-3.09) | 2.05 (1.87-2.24) |
| 17/11/2020 | 6.16 (4.76-7.62) | 10.58 (9.54-11.62) | 2.79 (2.54-3.05) | 1.84 (1.69-2.02) |
| 23/11/2020 | 6.88 (5.39-8.48) | 10.08 (9.11-11.12) | 2.90 (2.64-3.16) | 1.92 (1.74-2.11) |
| 01/12/2020 | 6.93 (5.40-8.52) | 11.20 (10.19-12.23) | 3.47 (3.21-3.78) | 2.23 (2.03-2.44) |
| 08/12/2020 | 6.31 (5.00-7.68) | 11.08 (10.05-12.09) | 3.83 (3.53-4.14) | 2.46 (2.24-2.73) |
| 15/12/2020 | 5.20 (3.93-6.66) | 7.08 (6.30-7.88) | 3.43 (3.17-3.69) | 2.44 (2.20-2.71) |
| 24/12/2020 | 3.32 (2.87-3.84) | 3.61 (3.32-3.95) | 2.75 (2.56-2.95) | 2.20 (2.00-2.41) |
| 29/12/2020 | 3.26 (2.84-3.71) | 3.11 (2.98-3.27) | 2.67 (2.47-2.87) | 1.92 (1.76-2.11) |
| 03/01/2021 | 3.75 (2.96-4.79) | 3.31 (3.03-3.62) | 2.80 (2.57-3.04) | 1.81 (1.65-1.99) |
| 11/01/2021 | 5.26 (3.96-6.85) | 3.38 (3.08-3.75) | 2.60 (2.39-2.83) | 1.68 (1.54-1.84) |
| 19/01/2021 | 5.21 (3.93-6.76) | 3.07 (2.89-3.27) | 2.54 (2.33-2.75) | 1.61 (1.48-1.77) |
| 26/01/2021 | 4.35 (3.39-5.50) | 3.16 (2.95-3.41) | 2.40 (2.20-2.60) | 1.63 (1.50-1.79) |
| 02/02/2021 | 4.77 (3.72-6.06) | 3.20 (2.97-3.48) | 2.41 (2.21-2.62) | 1.82 (1.64-2.03) |
| 09/02/2021 | 4.86 (3.67-6.45) | 3.44 (3.15-3.77) | 2.48 (2.25-2.71) | 1.87 (1.68-2.10) |
| 17/02/2021 | 4.10 (3.13-5.36) | 3.41 (3.13-3.72) | 2.44 (2.22-2.69) | 1.89 (1.72-2.07) |
| 23/02/2021 | 4.22 (3.26-5.41) | 3.11 (2.92-3.31) | 2.39 (2.20-2.60) | 1.82 (1.65-2.02) |
| 03/03/2021 | 5.46 (4.16-6.98) | 4.11 (3.67-4.61) | 2.47 (2.26-2.69) | 1.55 (1.42-1.69) |
| 09/03/2021 | 5.95 (4.60-7.52) | 7.38 (6.60-8.17) | 2.63 (2.40-2.88) | 1.67 (1.53-1.82) |
| 17/03/2021 | 6.53 (5.09-8.21) | 9.45 (8.50-10.45) | 2.55 (2.32-2.81) | 1.78 (1.63-1.94) |
| 23/03/2021 | 6.08 (4.70-7.73) | 8.96 (8.01-9.98) | 2.90 (2.64-3.19) | 1.67 (1.56-1.80) |

## References

1. Cabinet Office. National lockdown: Stay at Home. 4 Jan 2021 [cited 2 Feb 2021]. Available: https://www.gov.uk/guidance/national-lockdown-stay-at-home

2. Cabinet Office. Coronavirus (COVID-19): What has changed – 22 September. In: GOV.UK [Internet]. 22 Sep 2020 [cited 2 Feb 2021]. Available: https://www.gov.uk/government/news/coronavirus-covid-19-what-has-changed-22-september

3. Prime Minister’s Office. Prime Minister’s statement on coronavirus (COVID-19): 23 March 2020. In: GOV.UK [Internet]. 23 Mar 2020 [cited 2 Feb 2021]. Available: https://www.gov.uk/government/speeches/pm-address-to-the-nation-on-coronavirus-23-march-2020

4. Prime Minister’s Office. PM announces easing of lockdown restrictions: 23 June 2020. In: GOV.UK [Internet]. 23 Jun 2020 [cited 2 Feb 2021]. Available: https://www.gov.uk/government/news/pm-announces-easing-of-lockdown-restrictions-23-june-2020

5. UK Government. Coronavirus Act 2020. In: UK.gov [Internet]. 25 Mar 2020 [cited 2 Feb 2021]. Available: https://www.legislation.gov.uk/ukpga/2020/7/contents

6. Jarvis CI, Gimma A, van Zandvoort K, Wong KLM, CMMID COVID-19 working group, Edmunds WJ. The impact of local and national restrictions in response to COVID-19 on social contacts in England: a longitudinal natural experiment. BMC Med. 2021;19: 52.

7. Jarvis CI, Van Zandvoort K, Gimma A, Prem K, CMMID COVID-19 working group, Klepac P, et al. Quantifying the impact of physical distance measures on the transmission of COVID-19 in the UK. BMC Med. 2020;18: 124.

8. Mossong J, Hens N, Jit M, Beutels P, Auranen K, Mikolajczyk R, et al. Social contacts and mixing patterns relevant to the spread of infectious diseases. PLoS Med. 2008;5: e74.

9. R Core Team. R: A Language and Environment for Statistical Computing. Vienna, Austria: R Foundation for Statistical Computing; 2020. Available: https://www.R-project.org/

10. Efron B. Bootstrap Methods: Another Look at the Jackknife. The Annals of Statistics. 1979. pp. 1–26. doi:10.1214/aos/1176344552

11. Introduction to post-stratification. World Food Programme VAM; 2017. Available: https://docs.wfp.org/api/documents/WFP-0000121326/download/

12. WPP Annual Population by Age. World Population Prospects - Population Division - United Nations. 2019. Available: https://population.un.org/wpp/Download/Standard/Interpolated/

13. Official UK Coronavirus Dashboard. [cited 21 Apr 2021]. Available: https://coronavirus.data.gov.uk

14. Abbott S, Sherratt K, Palmer J, Martin-Nielsen R, Bevan J, Gibbs H, et al. covidregionaldata: Subnational Data for the COVID-19 Outbreak. -. 2020. p. -. doi:10.5281/zenodo.3957539

15. Jarvis C, Munday J, Gimma A, Wong K, Van Zandvoort K, Funk S, Edmunds WJ, CMMID working group. The effect of social distancing on the reproduction number and number of contacts in the UK from a social contact survey - Report for survey week 34. CMMD - London School of Hygiene and Tropical Medicine; 2020 Nov. Available: https://cmmid.github.io/topics/covid19/reports/comix/Comix%20Weekly%20Report%2034.p df

16. Real-time Assessment of Community Transmission findings. In: Imperial College London [Internet]. 2021 [cited 20 May 2021]. Available: https://www.imperial.ac.uk/medicine/research-and-impact/groups/react-study/real-time-assessment-of-community-transmission-findings/

17. Wood SN. Fast stable restricted maximum likelihood and marginal likelihood estimation of semiparametric generalized linear models. Journal of the Royal Statistical Society (B). 2011. pp. 3–36.

18. Wood SN. Generalized Additive Models: An Introduction with R. Chapman and Hall/CRC; 2017.

19. Klepac P, Kucharski AJ, Conlan AJK, Kissler S, Tang ML, Fry H, et al. Contacts in context: large-scale setting-specific social mixing matrices from the BBC Pandemic project. bioRxiv. medRxiv; 2020. doi:10.1101/2020.02.16.20023754

20. Wallinga J, Teunis P, Kretzschmar M. Using data on social contacts to estimate age-specific transmission parameters for respiratory-spread infectious agents. Am J Epidemiol. 2006;164: 936–944.

21. Diekmann O, Heesterbeek JAP, Roberts MG. The construction of next-generation matrices for compartmental epidemic models. J R Soc Interface. 2010;7: 873–885.

22. Davies NG, Klepac P, Liu Y, Prem K, Jit M, CMMID COVID-19 working group, et al. Age-dependent effects in the transmission and control of COVID-19 epidemics. Nat Med. 2020;26: 1205–1211.

23. Department of Health and Social Care. Face coverings to be mandatory in shops and supermarkets from 24 July. In: GOV.UK [Internet]. 14 Jul 2020 [cited 9 Mar 2021]. Available: https://www.gov.uk/government/speeches/face-coverings-to-be-mandatory-in-shops-and-supermarkets-from-24-july

24. Jarvis C, Gimma A, Van Zandvoort K, Wong KLM, CMMID COVID-19 working group, Edmunds WJ. The impact of local and national restrictions in response to COVID-19 on social contacts in the UK: a longitudinal natural experiment. In: CMMID Repository [Internet]. 19 Oct 2020 [cited 2 Feb 2021]. Available: https://cmmid.github.io/topics/covid19/comix-national-local-restrictions-in-the-UK.html

25. Sherratt K, Abbott S, Meakin SR, Hellewell J, Munday JD, Bosse N, et al. Exploring surveillance data biases when estimating the reproduction number: with insights into subpopulation transmission of Covid-19 in England. bioRxiv. medRxiv; 2020. doi:10.1101/2020.10.18.20214585

26. Hoang T, Coletti P, Melegaro A, Wallinga J, Grijalva CG, Edmunds JW, et al. A Systematic Review of Social Contact Surveys to Inform Transmission Models of Close-contact Infections. Epidemiology. 2019;30: 723–736.

27. Prem K, Cook AR, Jit M. Projecting social contact matrices in 152 countries using contact surveys and demographic data. PLoS Comput Biol. 2017;13: e1005697.

28. Liu CY, Berlin J, Kiti MC, Fava ED, Grow A, Zagheni E, et al. Rapid review of social contact patterns during the COVID-19 pandemic. bioRxiv. medRxiv; 2021. doi:10.1101/2021.03.12.21253410

29. Davies NG, Abbott S, Barnard RC, Jarvis CI, Kucharski AJ, Munday JD, et al. Estimated transmissibility and impact of SARS-CoV-2 lineage B.1.1.7 in England. Science. 2021;372. doi:10.1126/science.abg3055

