## Supplementary material for "CoMix: Changes in social contacts as measured by the contact survey during the COVID-19 pandemic in England between March 2020 and March 2021": Final survey versions for panels A&B, C&D, and E&F.

### Covid 19 behaviour and contact survey

**QCOUNTRY**
1. UK
2. BE
3. NL

**CAPTURE DATA FROM IMPORT:**

**Qsample=1 WAVE-TO-WAVE**
**Qsample=2 FRESH SAMPLE**

**Panel**
1. **Panel A**
2. **Panel B**

**Wave**
1. **Wave 1**
2. **Wave 2**
3. **Wave 3**
4. **Wave 4**
5. **Wave 5**
6. **Wave 6**
7. **Wave 7**
8. **Wave 8**

**Demographics**

*[Standard Screener: DO NOT MODIFY OR TRANSLATE]*
RESP_AGE [Hidden].
*ASK ALL*
*SA*
What is your age in years?
*YEAR/MONTH*

Hidden Question - RESP_AGE "this is a dummy question that will hold age"
USE RESP_AGE [Hidden] response list
1. _18 "18"
2. _19 "19"
3. _20 "20"
4. _21 "21"
5. _22 "22"
6. _23 "23"
7. _24 "24"

8. _25 "25"
9. . . .
10. _65 "65"
11. _66 "66"
12. _67 "67"
13. _68 "68"
14. _69 "69"
15. _70 "70"
16. _71 "71"
17. _72 "72"
18. _73 "73"
19. _74 "74"
20. _75 "75"
21. _76 "76"
22. _77 "77"
23. _78 "78"
24. _79 "79"
25. _80 "80"
26. _81 "81"
27. _82 "82"
28. _83 "83"
29. _84 "84"
30. _85 "85"
31. _86 "86"
32. _87 "87"
33. _88 "88"
34. _89 "89"
35. _90 "90"
36. _91 "91"
37. _92 "92"
38. _93 "93"
39. _94 "94"
40. _95 "95"
41. _96 "96"
42. _97 "97"
43. _98 "98"
44. _99 "99"
45. _100 "100"
46. _101 "101"
47. _102 "102"
48. _103 "103"
49. _104 "104"
50. _105 "105"
51. _998 "Age not calculated"
52. _999 "Age < 1"

*TERMINATE IS RESP_AGE < 18 YO*

*RECODE QUOTAGERANGE*

| |
|---|
| 18-24 |
| 25-34 |
| 35-44 |
| 45-54 |
| 55-64 |
| 65+ |

ASK FOR UK, BE AND NL

*SA*

*[Standard Screener: DO NOT MODIFY OR TRANSLATE]*

[PN: STOP_REALLOCATION if GENDER_NONBINARY is 3 or 4, and RESP_GENDER was not answered in a previous survey]

GENDER_NONBINARY. Which of the following describes how you think of yourself?

Select only one

1. _1 Male
2. _2 Female
3. _3 In another way
4. _4 Prefer not to answer

GENDER_NONBINARY_NL_BE. Which of the following describes how you think of yourself?

Select only one

1. _1 Male
2. _2 Female
3. _3 In another way
4. _4 Prefer not to answer

*[Standard Screener: DO NOT MODIFY OR TRANSLATE]*

*IF Qsample=1 HIDE AND IMPORT DATA, CREATE RECODE VARIABLE WITH THE NAME QMktSize_GB*

*IF Qsample=2 ASK QMktSize_GB*

[PN: If Prefer not to answer is selected, ask UKREGION1]

QMktSize_GB. Where do you live? Please note: This question may be considered personal. We would like to remind you that your participation is strictly voluntary and that your responses are used for research purposes only. The answers that you provide will be presented in aggregate form and none of them will be linked back to you in any way. All data will be collected and processed in adherence to the Market Research Society's Code of Conduct and the General Data Protection Regulation (GDPR).

| |
|---|
| **Postcode** |
| **Postal Town** |
| **Prefer Not to Answer** |

*[Standard Screener: DO NOT MODIFY OR TRANSLATE]*
*IF Qsample=1 HIDE AND IMPORT DATA, CREATE RECODE VARIABLE WITH THE NAME QMktSize_BE*

*IF Qsample=2 ASK QMktSize_BE*

QMktSize_BE. Where do you live?

- ☐ Province:
- ☐ Postal Code:
- ☐ City:

*[Standard Screener: DO NOT MODIFY OR TRANSLATE]*
*IF Qsample=1 HIDE AND IMPORT DATA, CREATE RECODE VARIABLE WITH THE NAME QMktSize_NL*

*IF Qsample=2 ASK QMktSize_NL*

QMKTSIZE_NL. Where do you live?

- ☐ Postal Code:
- ☐ City:
- ☐ Municipality:
- ☐ Prefer Not to Answer

#### MAIN INTRO SCREEN

Thank you for agreeing to take part in this important research about the new coronavirus (COVID-19) pandemic. Please take the time to carefully read the information below and the detailed survey participation notice.

The survey is being run by Ipsos on behalf of a group of experts in mathematical and statistical modelling of infectious diseases and public health – London School of Hygiene and Tropical Medicine (LSHTM; United Kingdom), University of Hasselt (Belgium), University of Antwerp (Belgium), National Institute for Public Health and Environment (RIVM; the Netherlands), University of Bern (Switzerland), and the ISI Foundation Turin (Italy).

The research is part of a larger programme of work funded by the European Commission which aims to provide urgently needed answers about the epidemiological characteristics of COVID-19, the social dynamics of the outbreak, and the related public health preparedness and response to the ongoing pandemic, as well as to assess its economic impact. It will be used directly on active communication and interaction with policy makers/authorities, other scientific groups and the general public, to help minimise the COVID-19’s public health, economic and social impact.

Taking part in this survey is completely voluntary, and you may refuse to do so. However, we would really value your support as the findings will provide invaluable information for decision making and strategies developed in the near future.

*NEW SCREEN*

#### Ethics form for participants

I confirm that I am 18 years or older.

I confirm that I have read and understood the information for this study. I have had the opportunity to consider the information before taking part in this study.

I understand that taking part is voluntary, and that I am free to withdraw at any time without giving any reason and without any of my rights being affected.

I understand that relevant sections of anonymized data collected during the study may be looked at by authorised individuals from the University of Antwerp, University of Hasselt, London School of Hygiene & Tropical Medicine, National Institute for Public Health and Environment in the Netherlands, University of Bern, and the ISI Foundation Turin.

I understand that ANONYMIZED data about me may be shared via a public data repository and that I and any of my household members or contacts will not be identifiable from this information.

I understand that this is a longitudinal study, and that I will be invited to complete multiple questionnaires over the course of the ongoing epidemic.

1. Agree to take part
2. Do not agree to take part

*SA*

Q3. What is your current employment status?

Select only one

1. _1 Employed full-time (34 hours or more)
2. _2 Employed part-time (less than 34 hours)
3. _3 Self employed
4. _4 Unemployed but looking for a job
5. _5 Unemployed and not looking for a job
6. _6 Full-time parent, homemaker
7. _7 Retired
8. _8 Student/Pupil
9. _9 Long-term sick or disabled

*ASK IN UK ONY*

*[Standard Screener: DO NOT MODIFY OR TRANSLATE]*

[To get UK01SG, need to ask EU01HINC. If EU01HINC=1,2, then ask UK01OCCR. If EU01HINC=3, ask UK01OCCHI. UK01SG is computed in the same manner as in the IIS panel. Details about that on panelstats.]

*[Standard Screener: DO NOT MODIFY OR TRANSLATE]*

[PN: if HHCMP10=1, then do not ask EU01HINC Q4 and assume EU01HINC Q4=_1]

*IF Qsample=1 HIDE AND IMPORT DATA, CREATE RECODE VARIABLE WITH THE NAME EU01HINC*
*IF Qsample=2 ASK EU01HINC*
*SA*
*EU01HINC.*

Are you the one in your household who has the highest income? [person with the largest income from employment, pensions, state benefits, investments or other sources]

Select only one

1. _1 Yes
2. _2 Yes, together with another household member
3. _3 No

*ASK IF IN UK AND EU01HINC=1 OR 2*
*[Standard Screener: DO NOT MODIFY OR TRANSLATE]*

*IF Qsample=1 HIDE AND IMPORT DATA, CREATE RECODE VARIABLE WITH THE NAME UK01OCCR.*
*IF Qsample=2 ASK UK01OCCR.*

*SA*
*UK01OCCR.*

In which of the below categories does your occupation fall? If retired or unemployed, please indicate the category closest to your previous occupation.

Select only one

1. USE UK01OCCR response list
2. Legislators, senior officials and managers [EXPANDABLE HEADER]
   a. _1100 Legislators and senior officials
3. Corporate managers [EXPANDABLE HEADER]
   a. _1210 Directors and chief executives
4. Production and operations department managers [EXPANDABLE HEADER]
   a. _1221 Production and operations department managers in agriculture, hunting, forestry and fishing
   b. _1222 Production and operations department managers in manufacturing
   c. _1223 Production and operations department managers in construction
   d. _1224 Production and operations department managers in wholesale and retail trade
   e. _1225 Production and operations department managers in restaurants and hotels
   f. _1226 Production and operations department managers in transport, storage and communications
   g. _1227 Production and operations department managers in business services

h. _1228 Production and operations department managers in personal care, cleaning and related services
i. _1229 Production and operations department managers not elsewhere classified

5. Other department managers [EXPANDABLE HEADER]
    a. _1231 Finance and administration department managers
    b. _1232 Personnel and industrial relations department managers
    c. _1233 Sales and marketing department managers
    d. _1234 Advertising and public relations department managers
    e. _1235 Supply and distribution department managers
    f. _1236 Computing services department managers
    g. _1237 Research and development department managers
    h. _1239 Other department managers not elsewhere classified
6. General managers [EXPANDABLE HEADER]
    a. _1311 General managers in agriculture, hunting, forestry/ and fishing
    b. _1312 General managers in manufacturing
    c. _1313 General managers in construction
    d. _1314 General managers in wholesale and retail trade
    e. _1315 General managers of restaurants and hotels
    f. _1316 General managers in transport, storage and communications
    g. _1317 General managers of business services
    h. _1318 General managers in personal care, cleaning and related services
    i. _1319 General managers not elsewhere classified
7. Physical, mathematical and engineering science professionals [EXPANDABLE HEADER]
    a. _2110 Physicists, chemists and related professionals
    b. _2120 Mathematicians, statisticians and related professionals
    c. _2130 Computing professionals
8. Architects, engineers and related professionals [EXPANDABLE HEADER]
    a. _2141 Architects, town and traffic planners
    b. _2142 Civil engineers
    c. _2143 Electrical engineers
    d. _2144 Electronics and telecommunications engineers
    e. _2145 Mechanical engineers
    f. _2146 Chemical engineers
    g. _2147 Mining engineers, metallurgists and related professionals
    h. _2148 Cartographers and surveyors
    i. _2149 Architects, engineers and related professionals not elsewhere classified
9. Life science and health professionals [EXPANDABLE HEADER]
    a. _2210 Life science professionals
    b. _2220 Health professionals (except nursing)
    c. _2230 Nursing and midwifery professionals
    d. _2300 Teaching professionals
10. Other professionals [EXPANDABLE HEADER]
    a. _2410 Business professionals
    b. _2420 Legal professionals
    c. _2430 Archivists, librarians and related information professionals
    d. _2440 Social science and related professionals
    e. _2450 Writers and creative or performing artists
    f. _2460 Religious professionals

11. Physical and engineering science associate professionals [EXPANDABLE HEADER]
    a. _3110 Physical and engineering science technicians
    b. _3120 Computer associate professionals
    c. _3130 Optical and electronic equipment operators
    d. _3140 Ship and aircraft controllers and technicians
    e. _3150 Safety and quality inspectors
    f. _3200 Life science and health associate professionals
    g. _3300 Teaching associate professionals
12. Other associate professionals [EXPANDABLE HEADER]
    a. _3410 Finance and sales associate professionals
    b. _3420 Business services agents and trade brokers
    c. _3430 Administrative associate professionals
    d. _3440 Customs, tax and related government associate professionals
    e. _3450 Police inspectors and detectives
    f. _3460 Social work associate professionals
    g. _3470 Artistic, entertainment and sports associate professionals
    h. _3480 Religious associate professionals
13. Clerks [EXPANDABLE HEADER]
    a. _4100 Office clerks
    b. _4200 Customer services clerks
14. Personal and protective services workers [EXPANDABLE HEADER]
    a. _5110 Travel attendants and related workers
    b. _5120 Housekeeping and restaurant services workers
    c. _5130 Personal care and related workers
    d. _5140 Other personal services workers
    e. _5160 Protective services workers
    f. _5200 Models, salespersons and demonstrators
15. Skilled agricultural and fishery workers [EXPANDABLE HEADER]
    a. _6000 Skilled agricultural and fishery workers
16. Craft and related trades workers [EXPANDABLE HEADER]
    a. _7100 Extraction and building trades workers
17. Metal, machinery and related trades workers [EXPANDABLE HEADER]
    a. _7210 Metal moulders, welders, sheet-metal workers, structural - metal preparers, and related trades workers
    b. _7220 Blacksmiths, tool-makers and related trades workers
    c. _7230 Machinery mechanics and fitters
    d. _7240 Electrical and electronic equipment mechanics and fitters
18. Precision, handicraft, printing and related trades workers [EXPANDABLE HEADER]
    a. _7310 Precision workers in metal and related materials
    b. _7320 Potters, glass-makers and related trades workers
    c. _7330 Handicraft workers in wood, textile, leather and related materials
    d. _7340 Printing and related trades workers
19. Other craft and related trades workers [EXPANDABLE HEADER]
    a. _7410 Food processing and related trades workers
    b. _7420 Wood treaters, cabinet-makers and related trades workers
    c. _7430 Textile, garment and related trades workers
    d. _7440 Pelt, leather and shoemaking trades workers
20. Plant and machine operators and assemblers [EXPANDABLE HEADER]

    a. _8000 Plant and machine operators and assemblers
21. Elementary occupations [EXPANDABLE HEADER]
    a. _9100 Sales and services elementary occupations
    b. _9200 Agricultural, fishery and related labourers
    c. _9300 Labourers in mining, construction, manufacturing and transport
22. Armed forces [EXPANDABLE HEADER]
    a. _9888 Armed forces
23. Did not work before [EXPANDABLE HEADER]
    a. _9991 Unemployed and not looking for a job / Long-term sick or disabled
    b. _9992 Pupil /Student/ in full time education
    c. _9993 Housewife

*ASK IF IN UK AND EU01HINC =3*

*[Standard Screener: DO NOT MODIFY OR TRANSLATE]*

[PN: Part of the occupation module, cannot be asked independent. Asked only if EU01HINC Q4=3. When respondent is also main earner (EU01HINC Q4=1 or 2), then values recorded in UK01OCCR are automatically transferred to UK01OCCHI. ]

*IF Qsample=1 HIDE AND IMPORT DATA, CREATE RECODE VARIABLE WITH THE NAME UK01OCCHI.*
*IF Qsample=2 ASK UK01OCCHI.*

*SA*
*UK01OCCHI.*

What is the occupation of the person with the highest income? If retired or unemployed, please indicate the category closest to his/her previous occupation.

Select only one

1. USE UK01OCCHI response list
2. Legislators, senior officials and managers [EXPANDABLE HEADER]
    a. _1100 Legislators and senior officials
3. Corporate managers [EXPANDABLE HEADER]
    a. _1210 Directors and chief executives
4. Production and operations department managers [EXPANDABLE HEADER]
    a. _1221 Production and operations department managers in agriculture, hunting, forestry and fishing
    b. _1222 Production and operations department managers in manufacturing
    c. _1223 Production and operations department managers in construction
    d. _1224 Production and operations department managers in wholesale and retail trade
    e. _1225 Production and operations department managers in restaurants and hotels
    f. _1226 Production and operations department managers in transport, storage and communications
    g. _1227 Production and operations department managers in business services
    h. _1228 Production and operations department managers in personal care, cleaning and related services
    i. _1229 Production and operations department managers not elsewhere classified
5. Other department managers [EXPANDABLE HEADER]

a. _1231 Finance and administration department managers
b. _1232 Personnel and industrial relations department managers
c. _1233 Sales and marketing department managers
d. _1234 Advertising and public relations department managers
e. _1235 Supply and distribution department managers
f. _1236 Computing services department managers
g. _1237 Research and development department managers
h. _1239 Other department managers not elsewhere classified

6. General managers [EXPANDABLE HEADER]
    a. _1311 General managers in agriculture, hunting, forestry/ and fishing
    b. _1312 General managers in manufacturing
    c. _1313 General managers in construction
    d. _1314 General managers in wholesale and retail trade
    e. _1315 General managers of restaurants and hotels
    f. _1316 General managers in transport, storage and communications
    g. _1317 General managers of business services
    h. _1318 General managers in personal care, cleaning and related services
    i. _1319 General managers not elsewhere classified
7. Physical, mathematical and engineering science professionals [EXPANDABLE HEADER]
    a. _2110 Physicists, chemists and related professionals
    b. _2120 Mathematicians, statisticians and related professionals
    c. _2130 Computing professionals
8. Architects, engineers and related professionals [EXPANDABLE HEADER]
    a. _2141 Architects, town and traffic planners
    b. _2142 Civil engineers
    c. _2143 Electrical engineers
    d. _2144 Electronics and telecommunications engineers
    e. _2145 Mechanical engineers
    f. _2146 Chemical engineers
    g. _2147 Mining engineers, metallurgists and related professionals
    h. _2148 Cartographers and surveyors
    i. _2149 Architects, engineers and related professionals not elsewhere classified
9. Life science and health professionals [EXPANDABLE HEADER]
    a. _2210 Life science professionals
    b. _2220 Health professionals (except nursing)
    c. _2230 Nursing and midwifery professionals
    d. _2300 Teaching professionals
10. Other professionals [EXPANDABLE HEADER]
    a. _2410 Business professionals
    b. _2420 Legal professionals
    c. _2430 Archivists, librarians and related information professionals
    d. _2440 Social science and related professionals
    e. _2450 Writers and creative or performing artists
    f. _2460 Religious professionals
11. Physical and engineering science associate professionals [EXPANDABLE HEADER]
    a. _3110 Physical and engineering science technicians
    b. _3120 Computer associate professionals
    c. _3130 Optical and electronic equipment operators

    d. _3140 Ship and aircraft controllers and technicians
    e. _3150 Safety and quality inspectors
    f. _3200 Life science and health associate professionals
    g. _3300 Teaching associate professionals
12. Other associate professionals [EXPANDABLE HEADER]
    a. _3410 Finance and sales associate professionals
    b. _3420 Business services agents and trade brokers
    c. _3430 Administrative associate professionals
    d. _3440 Customs, tax and related government associate professionals
    e. _3450 Police inspectors and detectives
    f. _3460 Social work associate professionals
    g. _3470 Artistic, entertainment and sports associate professionals
    h. _3480 Religious associate professionals
13. Clerks [EXPANDABLE HEADER]
    a. _4100 Office clerks
    b. _4200 Customer services clerks
14. Personal and protective services workers [EXPANDABLE HEADER]
    a. _5110 Travel attendants and related workers
    b. _5120 Housekeeping and restaurant services workers
    c. _5130 Personal care and related workers
    d. _5140 Other personal services workers
    e. _5160 Protective services workers
    f. _5200 Models, salespersons and demonstrators
15. Skilled agricultural and fishery workers [EXPANDABLE HEADER]
    a. _6000 Skilled agricultural and fishery workers
16. Craft and related trades workers [EXPANDABLE HEADER]
    a. _7100 Extraction and building trades workers
17. Metal, machinery and related trades workers [EXPANDABLE HEADER]
    a. _7210 Metal moulders, welders, sheet-metal workers, structural - metal preparers, and related trades workers
    b. _7220 Blacksmiths, tool-makers and related trades workers
    c. _7230 Machinery mechanics and fitters
    d. _7240 Electrical and electronic equipment mechanics and fitters
18. Precision, handicraft, printing and related trades workers [EXPANDABLE HEADER]
    a. _7310 Precision workers in metal and related materials
    b. _7320 Potters, glass-makers and related trades workers
    c. _7330 Handicraft workers in wood, textile, leather and related materials
    d. _7340 Printing and related trades workers
19. Other craft and related trades workers [EXPANDABLE HEADER]
    a. _7410 Food processing and related trades workers
    b. _7420 Wood treaters, cabinet-makers and related trades workers
    c. _7430 Textile, garment and related trades workers
    d. _7440 Pelt, leather and shoemaking trades workers
20. Plant and machine operators and assemblers [EXPANDABLE HEADER]
    a. _8000 Plant and machine operators and assemblers
21. Elementary occupations [EXPANDABLE HEADER]
    a. _9100 Sales and services elementary occupations
    b. _9200 Agricultural, fishery and related labourers

c. _9300 Labourers in mining, construction, manufacturing and transport

22. Armed forces [EXPANDABLE HEADER]
    a. _9888 Armed forces
23. Did not work before [EXPANDABLE HEADER]
    a. _9991 Unemployed and not looking for a job / Long-term sick or disabled
    b. _9992 Pupil /Student/ in full time education
    c. _9993 Housewife

UK01SG [Hidden]. Hidden Question:

Social Grade

1. _1 A - Upper middle class
2. _2 B - Middle class
3. _3 C1 - Lower middle class
4. _4 C2 - Skilled working class
5. _5 D - Working class
6. _6 E - Lower level of subsistence

*ASK IF IN BELGIUM AND EU01HINC=1 OR 2*

*[Standard Screener: DO NOT MODIFY OR TRANSLATE]*
*SA*
*IF Qsample=1 HIDE AND IMPORT DATA, CREATE RECODE VARIABLE WITH THE NAME BE02EDU.*
*IF Qsample=2 ASK BE02EDU.*

BE02EDU.

What is your highest level of education attained?

Select only one

1. _1 Without a diploma or primary education
2. _2 General lower secondary education (first 3 years completed)
3. _3 Technical, artistic or professional lower secondary education (first 3 years completed)
4. _4 General upper secondary education (6 years completed)
5. _5 Technical or artistic upper secondary education (6 years)
6. _6 Professional upper secondary (6 years)
7. _7 Higher education: graduat, candidature, bachelor
8. _8 University education: bachelor's degree, post-graduate, master's degree
9. _9 Complementary master
10. _10 Doctorate

*ASK IF IN BELGIUM AND EU01HINC=1 OR 2*
*[Standard Screener: DO NOT MODIFY OR TRANSLATE]*
*IF Qsample=1 HIDE AND IMPORT DATA, CREATE RECODE VARIABLE WITH THE NAME BE02OCCR.*
*IF Qsample=2 ASK BE02OCCR.*

BE02OCCR.

What is your occupation?

Select only one

1. USE BE02OCCR response list
2. Independent [Expandable Header]
   a. _1 farmer
   b. _2 craftsman, trader with 5 employees or less
   c. _3 industrial, wholesaler with 6 employees or more
   d. _4 liberal profession or profession for which qualification is required
3. Employee (public or private sector) [Expandable Header]
   a. _6 member of the general management, senior executive responsible for 5 employees or less
   b. _7 member of the general management, senior executive responsible for 6 to 10 employees
   c. _8 member of the general management, senior executive responsible for 11 employees or more
   d. _9 middle management, that is not part of the general management, responsible for 5 employees or less
   e. _10 middle management, that is not part of the general management, responsible for 6 employees or more
   f. _11 other employee who mainly performs office work
   g. _12 other employee who does not do office work (eg teacher, nurses ...)
4. Worker [Expandable Header]
   a. _13 skilled worker
   b. _14 non-skilled worker
5. Inactive [Expandable Header]
   a. _15 unable for work
   b. _16 pre-retired
   c. _17 retired
   d. _18 student
   e. _19 house man or housewife
   f. _20 unemployed
   g. _97 never worked
   h. _98 other
   i. _99 I do not know

*Ask if BE02OCCR =_16,_17 or _20, then ask BE02OCCR_LASTQ9.*
*[Standard Screener: DO NOT MODIFY OR TRANSLATE]*

[PN: if BE02OCCR=_16, _17 or _20, then ask BE02OCCR_LAST]

*[Standard Screener: DO NOT MODIFY OR TRANSLATE]*

[PN: if BE02OCCR=_16, _17 or _20, then ask BE02OCCR_LAST]

*SA*
*IF Qsample=1 HIDE AND IMPORT DATA, CREATE RECODE VARIABLE WITH THE NAME BE02OCCR_LAST.*
*IF Qsample=2 ASK BE02OCCR_LAST*

BE02OCCR_LAST.
What is the profession that you last exercised?

Select only one

1. Independent [Expandable Header]
   a. _1 farmer
   b. _2 craftsman, trader with 5 employees or less
   c. _3 industrial, wholesaler with 6 employees or more
   d. _4 liberal profession or profession for which qualification is required
2. Employee (public or private sector) [Expandable Header]
   a. _6 member of the general management, senior executive responsible for 5 employees or less
   b. _7 member of the general management, senior executive responsible for 6 to 10 employees
   c. _8 member of the general management, senior executive responsible for 11 employees or more
   d. _9 middle management, that is not part of the general management, responsible for 5 employees or less
   e. _10 middle management, that is not part of the general management, responsible for 6 employees or more
   f. _11 other employee who mainly performs office work
   g. _12 other employee who does not do office work (eg teacher, nurses ...)
3. Worker [Expandable Header]
   a. _13 skilled worker
   b. _14 non-skilled worker
4. Inactive [Expandable Header]
   a. _15 unable for work
   b. _18 student
   c. _19 house man or housewife
   d. _97 never worked
   e. _98 other
   f. _99 I do not know

*ASK IF IN BELGIUM AND EU01HINC=3*
*[Standard Screener: DO NOT MODIFY OR TRANSLATE]*
*SA*
*IF Qsample=1 HIDE AND IMPORT DATA, CREATE RECODE VARIABLE WITH THE NAME BE01EDU_ME.*
*IF Qsample=2 ASK BE01EDU_ME.*

BE01EDU_ME.

What is the highest education level of the main earner?

Select only one

1. _1 Without a diploma or primary education
2. _2 General lower secondary education (first 3 years completed)

3. _3 Technical, artistic or professional lower secondary education (first 3 years completed)
4. _4 General upper secondary education (6 years completed)
5. _5 Technical or artistic upper secondary education (6 years)
6. _6 Professional upper secondary (6 years)
7. _7 Higher education: graduat, candidature, bachelor
8. _8 University education: bachelor's degree, post-graduate, master's degree
9. _9 Complementary master
10. _10 Doctorate

*SA*
*ASK IF IN BELGIUM AND EU01HINC=3*
*[Standard Screener: DO NOT MODIFY OR TRANSLATE]*

[PN: Part of the occupation module, cannot be asked independent. Asked only if EU01HINC Q4=3. When respondent is also main earner (EU01HINC Q4=1 or 2), then values recorded in BE02OCCR Q9 are automatically transferred to BE02OCCHI Q11.]

*IF Qsample=1 HIDE AND IMPORT DATA, CREATE RECODE VARIABLE WITH THE NAME BE02OCCHI.*
*IF Qsample=2 ASK BE02OCCHI.*

BE02OCCHI.

What is the occupation of the main earner?

Select only one

1. USE BE02OCCHI response list
2. Independent [Expandable Header]
   a. _1 farmer
   b. _2 craftsman, trader with 5 employees or less
   c. _3 industrial, wholesaler with 6 employees or more
   d. _4 liberal profession or profession for which qualification is required
3. Employee (public or private sector) [Expandable Header]
   a. _6 member of the general management, senior executive responsible for 5 employees or less
   b. _7 member of the general management, senior executive responsible for 6 to 10 employees
   c. _8 member of the general management, senior executive responsible for 11 employees or more
   d. _9 middle management, that is not part of the general management, responsible for 5 employees or less
   e. _10 middle management, that is not part of the general management, responsible for 6 employees or more
   f. _11 other employee who mainly performs office work
   g. _12 other employee who does not do office work (eg teacher, nurses ...)
4. Worker [Expandable Header]
   a. _13 skilled worker
   b. _14 non-skilled worker
5. Inactive [Expandable Header]

a. _15 unable for work
b. _16 pre-retired
c. _17 retired
d. _18 student
e. _19 house man or housewife
f. _20 unemployed
g. _97 never worked
h. _98 other
i. _99 I do not know

*Ask if Q11=_16,_17 or _20, then ask BE02OCCHI_LAST*
*SA*
*[Standard Screener: DO NOT MODIFY OR TRANSLATE]*

[PN: if BE02OCCHI=_16, _17 or _20, then ask BE02OCCHI_LAST]

*IF Qsample=1 HIDE AND IMPORT DATA, CREATE RECODE VARIABLE WITH THE NAME BE02OCCHI_LAST.*
*IF Qsample=2 ASK BE02OCCHI_LAST.*

BE02OCCHI_LAST.

What is the profession that the main earner last exercised?

Select only one

1. Independent [Expandable Header]
   a. _1 farmer
   b. _2 craftsman, trader with 5 employees or less
   c. _3 industrial, wholesaler with 6 employees or more
   d. _4 liberal profession or profession for which qualification is required
2. Employee (public or private sector) [Expandable Header]
   a. _6 member of the general management, senior executive responsible for 5 employees or less
   b. _7 member of the general management, senior executive responsible for 6 to 10 employees
   c. _8 member of the general management, senior executive responsible for 11 employees or more
   d. _9 middle management, that is not part of the general management, responsible for 5 employees or less
   e. _10 middle management, that is not part of the general management, responsible for 6 employees or more
   f. _11 other employee who mainly performs office work
   g. _12 other employee who does not do office work (eg teacher, nurses ...)
3. Worker [Expandable Header]
   a. _13 skilled worker
   b. _14 non-skilled worker
4. Inactive [Expandable Header]
   a. _15 unable for work

b. _18 student
c. _19 house man or housewife
d. _97 never worked
e. _98 other
f. _99 I do not know

BE02SG [Hidden]. Hidden Question:

Social Grade new version

1. _1 Group 1&2
2. _2 Group 3&4
3. _3 Group 5&6
4. _4 Group 7&8
5. _9 Not allocated

*ASK IN NETHERLANDS ONY*
*ASK IF IN NETHERLANDS AND EU01HINC=1 OR 2*
*SA*
*[Standard Screener: DO NOT MODIFY OR TRANSLATE]*

*IF Qsample=1 HIDE AND IMPORT DATA, CREATE RECODE VARIABLE WITH THE NAME NL01OCCR.*
*IF Qsample=2 ASK NL01OCCR.*

NL01OCCR.

What is your occupation?

Select only one

1. USE NL01OCCR response list
2. Legislators, senior officials and managers [EXPANDABLE HEADER]
   a. _1100 Legislators and senior officials
3. Corporate managers [EXPANDABLE HEADER]
   a. _1210 Directors and chief executives
4. Production and operations department managers [EXPANDABLE HEADER]
   a. _1221 Production and operations department managers in agriculture, hunting, forestry and fishing
   b. _1222 Production and operations department managers in manufacturing
   c. _1223 Production and operations department managers in construction
   d. _1224 Production and operations department managers in wholesale and retail trade
   e. _1225 Production and operations department managers in restaurants and hotels
   f. _1226 Production and operations department managers in transport, storage and communications
   g. _1227 Production and operations department managers in business services
   h. _1228 Production and operations department managers in personal care, cleaning and related services
   i. _1229 Production and operations department managers not elsewhere classified

5. Other department managers [EXPANDABLE HEADER]
    a. _1231 Finance and administration department managers
    b. _1232 Personnel and industrial relations department managers
    c. _1233 Sales and marketing department managers
    d. _1234 Advertising and public relations department managers
    e. _1235 Supply and distribution department managers
    f. _1236 Computing services department managers
    g. _1237 Research and development department managers
    h. _1239 Other department managers not elsewhere classified
6. General managers [EXPANDABLE HEADER]
    a. _1311 General managers in agriculture, hunting, forestry/ and fishing
    b. _1312 General managers in manufacturing
    c. _1313 General managers in construction
    d. _1314 General managers in wholesale and retail trade
    e. _1315 General managers of restaurants and hotels
    f. _1316 General managers in transport, storage and communications
    g. _1317 General managers of business services
    h. _1318 General managers in personal care, cleaning and related services
    i. _1319 General managers not elsewhere classified
7. Physical, mathematical and engineering science professionals [EXPANDABLE HEADER]
    a. _2110 Physicists, chemists and related professionals
    b. _2120 Mathematicians, statisticians and related professionals
    c. _2130 Computing professionals
8. Architects, engineers and related professionals [EXPANDABLE HEADER]
    a. _2141 Architects, town and traffic planners
    b. _2142 Civil engineers
    c. _2143 Electrical engineers
    d. _2144 Electronics and telecommunications engineers
    e. _2145 Mechanical engineers
    f. _2146 Chemical engineers
    g. _2147 Mining engineers, metallurgists and related professionals
    h. _2148 Cartographers and surveyors
    i. _2149 Architects, engineers and related professionals not elsewhere classified
9. Life science and health professionals [EXPANDABLE HEADER]
    a. _2210 Life science professionals
    b. _2220 Health professionals (except nursing)
    c. _2230 Nursing and midwifery professionals
    d. _2300 Teaching professionals
10. Other professionals [EXPANDABLE HEADER]
    a. _2410 Business professionals
    b. _2420 Legal professionals
    c. _2430 Archivists, librarians and related information professionals
    d. _2440 Social science and related professionals
    e. _2450 Writers and creative or performing artists
    f. _2460 Religious professionals
11. Physical and engineering science associate professionals [EXPANDABLE HEADER]
    a. _3110 Physical and engineering science technicians
    b. _3120 Computer associate professionals

    c. _3130 Optical and electronic equipment operators
    d. _3140 Ship and aircraft controllers and technicians
    e. _3150 Safety and quality inspectors
    f. _3200 Life science and health associate professionals
    g. _3300 Teaching associate professionals
12. Other associate professionals [EXPANDABLE HEADER]
    a. _3410 Finance and sales associate professionals
    b. _3420 Business services agents and trade brokers
    c. _3430 Administrative associate professionals
    d. _3440 Customs, tax and related government associate professionals
    e. _3450 Police inspectors and detectives
    f. _3460 Social work associate professionals
    g. _3470 Artistic, entertainment and sports associate professionals
    h. _3480 Religious associate professionals
13. Clerks [EXPANDABLE HEADER]
    a. _4100 Office clerks
    b. _4200 Customer services clerks
14. Personal and protective services workers [EXPANDABLE HEADER]
    a. _5110 Travel attendants and related workers
    b. _5120 Housekeeping and restaurant services workers
    c. _5130 Personal care and related workers
    d. _5140 Other personal services workers
    e. _5160 Protective services workers
    f. _5200 Models, salespersons and demonstrators
15. Skilled agricultural and fishery workers [EXPANDABLE HEADER]
    a. _6000 Skilled agricultural and fishery workers
16. Craft and related trades workers [EXPANDABLE HEADER]
    a. _7100 Extraction and building trades workers
17. Metal, machinery and related trades workers [EXPANDABLE HEADER]
    a. _7210 Metal moulders, welders, sheet-metal workers, structural - metal preparers, and related trades workers
    b. _7220 Blacksmiths, tool-makers and related trades workers
    c. _7230 Machinery mechanics and fitters
    d. _7240 Electrical and electronic equipment mechanics and fitters
18. Precision, handicraft, printing and related trades workers [EXPANDABLE HEADER]
    a. _7310 Precision workers in metal and related materials
    b. _7320 Potters, glass-makers and related trades workers
    c. _7330 Handicraft workers in wood, textile, leather and related materials
    d. _7340 Printing and related trades workers
19. Other craft and related trades workers [EXPANDABLE HEADER]
    a. _7410 Food processing and related trades workers
    b. _7420 Wood treaters, cabinet-makers and related trades workers
    c. _7430 Textile, garment and related trades workers
    d. _7440 Pelt, leather and shoemaking trades workers
20. Plant and machine operators and assemblers [EXPANDABLE HEADER]
    a. _8000 Plant and machine operators and assemblers
21. Elementary occupations [EXPANDABLE HEADER]
    a. _9100 Sales and services elementary occupations

    b. _9200 Agricultural, fishery and related labourers
    c. _9300 Labourers in mining, construction, manufacturing and transport
22. Armed forces [EXPANDABLE HEADER]
    a. _9888 Armed forces
23. Not working [EXPANDABLE HEADER]
    a. _9990 Unemployed but looking for a job
    b. _9991 Unemployed and not looking for a job / Long-term sick or disabled
    c. _9992 Pupil /Student/ in full time education
    d. _9993 Housewife
    e. _9994 Retired

*ASK IF IN NETHERLANDS AND EU01HINC =1 OR 2*
*[Standard Screener: DO NOT MODIFY OR TRANSLATE]*

[PN: Part of the occupation module, cannot be asked independent. Asked only if EU01HINC=3. When respondent is also main earner (EU01HINC=1 or 2), then values recorded in NL01OCCR are automatically transferred to NL01OCCHI. ]

*IF Qsample=1 HIDE AND IMPORT DATA, CREATE RECODE VARIABLE WITH THE NAME NL01OCCHI.*
*IF Qsample=2 ASK NL01OCCHI.*

NL01OCCHI.

What is the occupation of the person with the highest income?

Select only one

1. USE NL01OCCHI response list
2. Legislators, senior officials and managers [EXPANDABLE HEADER]
    a. _1100 Legislators and senior officials
3. Corporate managers [EXPANDABLE HEADER]
    a. _1210 Directors and chief executives
4. Production and operations department managers [EXPANDABLE HEADER]
    a. _1221 Production and operations department managers in agriculture, hunting, forestry and fishing
    b. _1222 Production and operations department managers in manufacturing
    c. _1223 Production and operations department managers in construction
    d. _1224 Production and operations department managers in wholesale and retail trade
    e. _1225 Production and operations department managers in restaurants and hotels
    f. _1226 Production and operations department managers in transport, storage and communications
    g. _1227 Production and operations department managers in business services
    h. _1228 Production and operations department managers in personal care, cleaning and related services
    i. _1229 Production and operations department managers not elsewhere classified
5. Other department managers [EXPANDABLE HEADER]
    a. _1231 Finance and administration department managers
    b. _1232 Personnel and industrial relations department managers
    c. _1233 Sales and marketing department managers
    d. _1234 Advertising and public relations department managers
    e. _1235 Supply and distribution department managers
    f. _1236 Computing services department managers

g. _1237 Research and development department managers
h. _1239 Other department managers not elsewhere classified

6. General managers [EXPANDABLE HEADER]
    a. _1311 General managers in agriculture, hunting, forestry/ and fishing
    b. _1312 General managers in manufacturing
    c. _1313 General managers in construction
    d. _1314 General managers in wholesale and retail trade
    e. _1315 General managers of restaurants and hotels
    f. _1316 General managers in transport, storage and communications
    g. _1317 General managers of business services
    h. _1318 General managers in personal care, cleaning and related services
    i. _1319 General managers not elsewhere classified
7. Physical, mathematical and engineering science professionals [EXPANDABLE HEADER]
    a. _2110 Physicists, chemists and related professionals
    b. _2120 Mathematicians, statisticians and related professionals
    c. _2130 Computing professionals
8. Architects, engineers and related professionals [EXPANDABLE HEADER]
    a. _2141 Architects, town and traffic planners
    b. _2142 Civil engineers
    c. _2143 Electrical engineers
    d. _2144 Electronics and telecommunications engineers
    e. _2145 Mechanical engineers
    f. _2146 Chemical engineers
    g. _2147 Mining engineers, metallurgists and related professionals
    h. _2148 Cartographers and surveyors
    i. _2149 Architects, engineers and related professionals not elsewhere classified
9. Life science and health professionals [EXPANDABLE HEADER]
    a. _2210 Life science professionals
    b. _2220 Health professionals (except nursing)
    c. _2230 Nursing and midwifery professionals
    d. _2300 Teaching professionals
10. Other professionals [EXPANDABLE HEADER]
    a. _2410 Business professionals
    b. _2420 Legal professionals
    c. _2430 Archivists, librarians and related information professionals
    d. _2440 Social science and related professionals
    e. _2450 Writers and creative or performing artists
    f. _2460 Religious professionals
11. Physical and engineering science associate professionals [EXPANDABLE HEADER]
    a. _3110 Physical and engineering science technicians
    b. _3120 Computer associate professionals
    c. _3130 Optical and electronic equipment operators
    d. _3140 Ship and aircraft controllers and technicians
    e. _3150 Safety and quality inspectors
    f. _3200 Life science and health associate professionals
    g. _3300 Teaching associate professionals
12. Other associate professionals [EXPANDABLE HEADER]
    a. _3410 Finance and sales associate professionals

b. _3420 Business services agents and trade brokers
c. _3430 Administrative associate professionals
d. _3440 Customs, tax and related government associate professionals
e. _3450 Police inspectors and detectives
f. _3460 Social work associate professionals
g. _3470 Artistic, entertainment and sports associate professionals
h. _3480 Religious associate professionals

13. Clerks [EXPANDABLE HEADER]
    a. _4100 Office clerks
    b. _4200 Customer services clerks
14. Personal and protective services workers [EXPANDABLE HEADER]
    a. _5110 Travel attendants and related workers
    b. _5120 Housekeeping and restaurant services workers
    c. _5130 Personal care and related workers
    d. _5140 Other personal services workers
    e. _5160 Protective services workers
    f. _5200 Models, salespersons and demonstrators
15. Skilled agricultural and fishery workers [EXPANDABLE HEADER]
    a. _6000 Skilled agricultural and fishery workers
16. Craft and related trades workers [EXPANDABLE HEADER]
    a. _7100 Extraction and building trades workers
17. Metal, machinery and related trades workers [EXPANDABLE HEADER]
    a. _7210 Metal moulders, welders, sheet-metal workers, structural - metal preparers, and related trades workers
    b. _7220 Blacksmiths, tool-makers and related trades workers
    c. _7230 Machinery mechanics and fitters
    d. _7240 Electrical and electronic equipment mechanics and fitters
18. Precision, handicraft, printing and related trades workers [EXPANDABLE HEADER]
    a. _7310 Precision workers in metal and related materials
    b. _7320 Potters, glass-makers and related trades workers
    c. _7330 Handicraft workers in wood, textile, leather and related materials
    d. _7340 Printing and related trades workers
19. Other craft and related trades workers [EXPANDABLE HEADER]
    a. _7410 Food processing and related trades workers
    b. _7420 Wood treaters, cabinet-makers and related trades workers
    c. _7430 Textile, garment and related trades workers
    d. _7440 Pelt, leather and shoemaking trades workers
20. Plant and machine operators and assemblers [EXPANDABLE HEADER]
    a. _8000 Plant and machine operators and assemblers
21. Elementary occupations [EXPANDABLE HEADER]
    a. _9100 Sales and services elementary occupations
    b. _9200 Agricultural, fishery and related labourers
    c. _9300 Labourers in mining, construction, manufacturing and transport
22. Armed forces [EXPANDABLE HEADER]
    a. _9888 Armed forces
23. Not working [EXPANDABLE HEADER]
    a. _9990 Unemployed but looking for a job
    b. _9991 Unemployed and not looking for a job / Long-term sick or disabled

c. _9992 Pupil /Student/ in full time education
d. _9993 Housewife
e. _9994 Retired

*[Standard Screener: DO NOT MODIFY OR TRANSLATE]*

[To get NL01SG, need to ask EU01HINC. If EU01HINC=1,2, then ask NL01OCCR. If EU01HINC=3, ask NL01OCCHI. NL01SG is computed in the same manner as in the IIS panel. Details about that on panelstats.]

NL01SG [Hidden]. Hidden Question:

Social Grade

1. _1 Managers and professionals
2. _2 Technicians, Clerks, Service workers
3. _3 Workers, Elementary occupations, Armed forces
4. _4 Inactive / Unemployed

*MQB*

*ASK IF GENDER_NONBINARY Q2=2 OR 3 AND RESPAGE<55*
*SA*
PREGNANCY.
Q15. Are you currently pregnant?

Select only one

1. _1 Yes
2. _2 No
3. _3 Prefer not to answer

*ASK ALL*
*SA*
*[Standard Screener: DO NOT MODIFY OR TRANSLATE]*
*IF Qsample=1 HIDE AND IMPORT DATA, CREATE RECODE VARIABLE WITH THE NAME INCOMEUK.*
*IF Qsample=2 ASK INCOMEUK.*

INCOMEUK.
The next question may be considered personal, but it is not mandatory to answer. If you do, we assure you that your responses will be kept strictly confidential and used for research purposes only. What is the COMBINED TOTAL ANNUAL INCOME (pre-tax, pensions, or national insurance deductions) earned by all members of your family? Here, family is defined as everyone related by marriage, civil partnership, birth, or adoption living at the same address. Please include all your income sources: salaries, scholarships, pension and Social Security benefits, dividends from shares, income from rental properties, child support and alimony etc.

Select only one

1. _1 Under £5,000
2. _2 £5,000 - £9,999
3. _3 £10,000 - £14,999
4. _4 £15,000 - £19,999
5. _5 £20,000 - £24,999
6. _6 £25,000 - £34,999
7. _7 £35,000 - £44,999

8. _8 £45,000 - £54,999
9. _9 £55,000 - £64,999
10. _9 £65,000 - £99,999
11. _10 £100,000 or more
12. _11 Prefer not to answer

BE01INC

Single response

Into which category does your TOTAL NET HOUSEHOLD monthly income from all sources fall (i.e. income after tax deduction)?

*Please include all your income sources : salaries, scholarships, pension and Social Security benefits, dividends from shares, income from rental properties, child support and alimony etc.*

_1 € 0 - € 549

_2 € 550 - € 999

_3 € 1 000 - € 1 299

_4 € 1 300 - € 1 499

_5 € 1 500 - € 1 699

_6 € 1 700 - € 1 899

_7 € 1 900 - € 2 199

_8 € 2 200 - € 2 499

_9 € 2 500 - € 2 799

_10 € 2 800 - € 3 199

_11 € 3 200 - € 3 699

_12 € 3 700 - € 4 499

_13 € 4 500 - € 5 499

_14 € 5 500 - € 7 999

_15 € 8 000 or more

_16 Prefer not to answer

NL01INC

Single response

Into which category does your TOTAL HOUSEHOLD pre-tax annual income from all sources fall (i.e. income before tax deduction)?

[Please include all your income sources : salaries, scholarships, pension and Social Security benefits, dividends from shares, income from rental properties, child support and alimony etc.]

_1 0 € - 6000 €

_2 6001€ – 9000 €

_3 9001 € – 12000 €

_4 12001 € – 15000 €

_5 15001 € – 18000 €

_6 18001 € – 21000 €

_7 21001 € – 24000 €
_8 24001 € – 30000 €
_9 30001 € – 36000 €
_10 36001 € – 48000 €
_11 48001 € – 60000 €
_12 60001 € – 120000 €
_13 120001 € or more
_14 Prefer not to answer

*ASK ALL*
*SA*
QHOUSE
Q20. Not including you, how many other people live in your household? By household, we mean anyone living at the same address as you, that you share a kitchen with.
Select only one

None
1
2
3
4
5
6
7
8
9
10
11 or more

*ASK IF Q20=1 OR MORE (I.E. IF NOT A SINGLE PERSON HOUSEHOLD)*
*SA*
Q20a . Which of the following best describes your household?
Select only one

2. Two or more non-family adults
3. Couple with dependent children
4. Couple with independent children only
5. Couple with no children
6. Lone parent with dependent children
7. Lone parent with independent children only
8. Households containing two or more families
9. Other

*HAVE ONE ROW FOR EACH PERSON IN QHOUSE*
*LIMIT TO 11 ANSWER BOXES IF QHOUSE=11*
*WRITE IN*

QNAME
Q21. Please write the nickname of each other person in your household.
Note that this nickname is only needed to make it easier for you to complete the survey, so please pick a nickname that will help you identify each household member later in the questionnaire. Nicknames are not visible to anyone outside of this survey.

Please write in
1. NAME1
2. NAME2
3. NAME3
etc.

*ASK THE FOLLOWING LOOP Q23 TO Q27 FOR EACH NAME GIVEN AT Q21*
*SA*

QHHAGE

Q23. Which of the following age groups do they fit into?

1. Under 1
2. 1-4
3. 5-9
4. 10-14
5. 15-19
6. 20-24
7. 25-34
8. 35-44
9. 45-54
10. 55-64
11. 65-69
12. 70-74
13. 75-79
14. 80-84
15. 85+
16. Don’t know
17. Prefer not to answer

*SA*
QHHGENDER
Q24. As far as you know, which of the following describes how [NAME] thinks of themselves?
Select only one
1. _1 Male
2. _2 Female
3. _3 In another way
4. _4 Prefer not to answer
5. _5 Don’t know

*ONLY ASK THOSE AGED 16 AND OVER AT QHHAGE*
*SA*
Q25. What is [NAME]’s current employment status?
Select only one

1. _1 Employed full-time (34 hours or more)
2. _2 Employed part-time (less than 34 hours)
3. _3 Self employed
4. _4 Unemployed but looking for a job
5. _5 Unemployed and not looking for a job
6. _6 Full-time parent, homemaker
7. _7 Retired
8. _8 Student/Pupil
9. _9 Long-term sick or disabled

*SA*
QHHATTEND
Q26. Does [NAME] attend any of the following as a pupil or student?
Select only one
1. Nursery or pre-school [*UK ONLY SHOW THIS OPTION FOR THOSE AGED 4 OR UNDER AT QHHAGE]*
2. School [*UK ONLY SHOW THIS OPTION FOR THOSE AGED 4 TO 21 AT QHHAGE] [BE AND NL SHOW THIS OPTION FOR THOSE AGED 4-14 ONLY]*
3. Further education, e.g. college [*UK ONLY SHOW THIS OPTION FOR THOSE AGED 14 AND OVER AT QHHAGE] [DO NOT SHOW FOR BE AND NL]*
4. Higher education, e.g. university [*UK, BE AND NL SHOW THIS OPTION FOR THOSE AGED 14 AND OVER AT QHHAGE]*
5. None of the above
6. Don’t know
7. Prefer not to answer
*8. Secondary school (BE AND NL ONLY. SHOW THIS OPTION FOR THOSE AGED 10 AND OVER AT QHHAGE _ [PN. Show after option 2]*

*ASK IF Q24=2 OR 3 (NOT MALE) AND QHHAGE = CODES 5-9 (I.E AGED 15 TO 54)*
*SA*
QHHPREGNANT
Q27. Is [NAME] currently pregnant?
Select only one
1. Yes
2. No
3. Don’t know
4. Prefer not to answer

*ASK ALL*
*SA*
FLU
Q28. Are you or any other household member in a high-risk group under which the annual influenza vaccine would usually be offered by the NHS?
High risk groups include individuals with: chronic respiratory disease, chronic heart disease, chronic kidney disease, chronic liver disease, chronic neurological disease, diabetes (all types), immunosuppression (due to disease or treatment), asplenia or dysfunction of the spleen, class III obesity (BMI ≥ 40), and pregnant women.

Select only one for each row

*ASK FOR SELF AND EACH PERSON NAMED AT Q21*

*NOTE FOR SCRIPTER – PLEASE SCRIPT AS GRID*

*ROWS:*
0. Yourself
1. Name 1
2. Name 2
etc.

*COLUMNS:*
1. Yes
2. No
3. Don't know
4. Prefer not to answer

**Symptoms**
*ASK ALL*
*MA*
SYMPTOMS
Q29. Have you, or anyone else in your household, had any of the following symptoms in the last seven days?
Please tick all that apply for each row
*ASK FOR SELF AND EACH PERSON NAMED AT Q21*
*NOTE FOR SCRIPTER – PLEASE SCRIPT AS GRID – ROTATE COLUMNS 1-7*

*ROWS:*
0. Yourself
1. Name 1
2. Name 2
etc.

*COLUMNS:*
1. Fever or high temperature
2. A cough that has lasted for at least several hours

3. Shortness of breath
4. Aches and pains, e.g. in back, neck, shoulders or joints
5. Blocked nose
6. Sore throat
7. Feeling unusually tired
8. None of these
9 Don't know
10. Prefer not to answer

*ASK IF Q29=1,2,3,4,5,6 OR 7 (ALL WHO HAVE SUFFERED ANY SYMPTOMS)*

*MA*
SERVICE
Q30. Have you, or anyone else in your household, done any of the following for these symptoms?
*NOTE FOR SCRIPTER – PLEASE SCRIPT AS GRID – 6 MUST ALWAYS BE LAST*
Please tick all that apply for each row

*ROWS:*
0. Yourself
1. Name 1
2. Name 2
etc.

*COLUMNS:*
1. Phoned NHS 111 or used NHS 111 online service
2. Phoned a GP practice/GP out of hours service
3. Visited a GP practice/GP out of hours service
4. Visited a walk-in centre, urgent care centre, urgent treatment centre or minor injuries unit (PN. DO NOT SHOW IN BE or NL)
5. Visited Accident & Emergency (A&E) 6. Visited a testing location somewhere different to these services
7. Been admitted to hospital
8. Don't know
9. None of these
10. Prefer not to answer

*ASK IF Q30=1,2,3,4,5,6 OR 7 (FOR EACH SERVICE SELECTED AT SERVICE)*
*WRITE IN*
WHEN
Q31. You said that [*IF FOR SELF AT Q30 –* you *OR IF FOR [Q21] AT Q30* – NAME] have/has [*INSERT SERVICE FROM Q30*]. When did [you/they] do that? If you don't know exactly, please provide an approximate date.
Please write in
Please use the format "DD/MM"
INSERT DATE USE DATE FORMAT DD/MM. EARLIEST DATE IS 01/02 AND CANNOT BE A DATE IN THE FUTURE
1. Don't know
2. Prefer not to answer

*ASK ALL*
*SA*
TEST
Q32. Have you, or any other household member, ever been tested for Coronavirus (Covid-19)?

Select only one

1. Yes
2. No

*ASK IF Q32=1*

*ASK FOR SELF AND EACH PERSON NAMED AT Q21*
*SA*
TESTPERSON
Q33. Who has been tested for Coronavirus (Covid-19)?
*ASK FOR SELF AND EACH PERSON NAMED AT Q21*
*NOTE FOR SCRIPTER – PLEASE SCRIPT AS GRID*
Select only one for each row

*ROWS:*
0. Yourself
1. Name 1
2. Name 2
etc.

*COLUMNS:*
1. Tested and the test showed I/they have Coronavirus
2. Tested, and the test showed I/they <u>do not</u> have Coronavirus
3. Yes, and I'm still waiting to hear the result
4. Not tested
5. Don't know
6. Prefer not to answer

*ASK FOR SELF AND EACH PERSON NAMED AT Q21*
*SA*
CONTACT
Q34. To the best of your knowledge, do you think you or anyone else in your household have been in direct contact with someone who has Coronavirus (Covid-19) in the last seven days, or know someone close to them who has Coronavirus (Covid-19)?
Select only one for each row

*ASK FOR SELF AND EACH PERSON NAMED AT Q21*
*NOTE FOR SCRIPTER – PLEASE SCRIPT AS GRID*

*ROWS:*
0. Yourself
1. Name 1

2. Name 2
etc.

*COLUMNS:*
1. Yes, currently infected
2. Yes, passed away
3. Yes, recovered
4. No
5. Don't know
6. Prefer not to answer

**Attitudes**

*ASK ALL*
*SA*
ATT1
Q35. To what extent do you agree or disagree with each of the following statements?
*REVERSE ORDER OF SCALE. ROTATE STATEMENTS*
Select only one for each row

1. Strongly agree
2. Tend to agree
3. Neither agree nor disagree
4. Tend to disagree
5. Strongly disagree
6. Don't know

1. Coronavirus would be a serious illness for me
2. I am likely to catch coronavirus
3. If I don't follow the government's advice, I might spread coronavirus to someone who is vulnerable

*ASK ALL*
*SA*
EFFECT
Q36. How effective, if at all, do you think each of the following are at slowing the spread of coronavirus?
*REVERSE ORDER OF SCALE. ROTATE STATEMENTS*
Select only one for each row

1. Very effective
2. Fairly effective
3. Not very effective
4. Not at all effective
5. Don't know

1. Reducing the number of people you meet
2. Staying at home for 7 days if you have a mild symptom such as a mild cough

3. Staying at home for 7 days if you have more severe symptoms such as a severe cough or a high temperature
4. Avoiding crowded places
5. Stay at home for 14 days if anyone other than yourself in your household has mild symptom such as a mild cough
6. Stay at home for 14 days if anyone other than yourself in your household has severe symptoms such as a cough or a high temperature
7. School closures
8. Closing bars, restaurants, cinemas etc.
9. Banning the use of public transport
10. Banning international travel into [the UK/Belgium/ the Netherlands]
11. Banning travel within [the UK/Belgium/ the Netherlands]

*ASK ALL*
*SA*
CONF
Q37. How confident are you, if at all, that if you wanted to you could…?
*REVERSE ORDER OF SCALE. ROTATE STATEMENTS*
Select only one for each row

1. Very confident
2. Fairly confident
3. Not very confident
4. Not at all confident
5. Don't know

1. Reduce the number of people you meet
2. Stay at home for 7 days if you have a mild symptom such as a mild cough
3. Stay at home for 7 days if you have more severe symptoms such as a severe cough or a high temperature
4. Avoid crowded places
5. Stay at home for 14 days if anyone other than yourself in your household has mild symptom such as a mild cough
6. Stay at home for 14 days if anyone other than yourself in your household has severe symptoms such as cough or a high temperature
7. Not use public transport

*ASK ALL*
*SA*
ATT2
Q38. To what extent do you agree or disagree with each of the following statements?
*REVERSE ORDER OF SCALE. ROTATE STATEMENTS*
Select only one for each row

1. Strongly agree
2. Tend to agree
3. Neither agree nor disagree
4. Tend to disagree
5. Strongly disagree
6. Don't know

1. Other people I work with expect me to work, even when I am ill *SHOW IF Q3 = 1,2,3*
2. If I could not work because of coronavirus, I would still get paid *SHOW IF Q3 = 1,2,3*
3. If I had to isolate myself for 7 days because of coronavirus, someone else would be able to look after my children *ONLY SHOW THIS OPTION FOR THOSE CODING 1-5 AT Q23*
4. If I had to isolate myself for 7 days, this would cause problems for other people who I don't know
5. I have enough food and supplies to last for 7 days, if I had to isolate myself

**Behaviour**

*ASK ALL*
*SA*
*INTRO*
Q39. You may have been asked or decided to participate in different responses to coronavirus (covid-19).
Thinking about the last seven days, please select the appropriate response for each of the interventions listed below.

INTER In the last seven days, [*IF FOR SELF –* have you] [*IF FOR NAME 1/2 ETC AT Q21 –* has NAME] been asked to…
*ASK FOR SELF AND EACH PERSON NAMED AT Q21*
*NOTE FOR SCRIPTER – PLEASE SCRIPT AS GRID*

Select only one for each row

*ROWS:*
1. Quarantine [yourself/themselves]
*Quarantine is the act of staying at home after a potential exposure to an infected case. If you are in quarantine, you can leave the house, but limit your movements.*

2. Isolate [yourself/themselves]
*Isolation is the act of separating yourself from people who are not infected, including any household members. You can be in isolation in your house or in a health facility.*
3. Work from home due to coronavirus or limit your/their time at your/their workplace *SHOW IF Q3 = 1,2,3 OR Q25=1,2,3*
4. Limit [your/their] time at the [university or college] [*SHOW IF Q3 = 8 OR Q26 = 3 OR 4*] OR [pre-school or nursery] [*Q26 = 1]* OR [school] [*Q26 = 2*] OR [*Secondary* school] [IF NL OR BE AND *Q26 = 8*]due to coronavirus (covid-19)

*COLUMNS:*
1. Yes
2. No
3. Not applicable *ONLY SHOW FOR THIRD STATEMENT*
4. Don't know
5. Prefer not to answer

*ASK FOR SELF AND EACH PERSON NAMED AT Q21*
*SA*
INTER2

Q40. In the last seven days, has…
Select only one for each row

*NOTE FOR SCRIPTER – PLEASE SCRIPT AS GRID*
*ROWS:*
1. [Your/ NAME's] workplace been closed due to coronavirus (covid-19) for at least one day *ASK IF Q3-CODE 1,2,3 OR Q25 = 1,2,3*
2. [Your/-NAME's] university or college] [*ASK IF Q3 = 8 OR Q26 = 3 OR 4]* OR [pre-school or nursery] [*ASK IF Q26 = 1*] OR [school] [*ASK IF Q26 = 2*] OR [*Secondary* school] [AKS IF IF NL OR BE AND *Q26 = 8*] been closed for at least one day

*COLUMNS:*
1. Yes
2. No
3. Not applicable *ONLY SHOW FOR FIRST STATEMENT*
4. Don't know
5. Prefer not to answer

*ASK FOR SELF AND EACH PERSON NAMED AT Q21*
*SA*
INTER3
Q41. In the last seven days, [*IF FOR SELF –* have you] [*IF FOR NAME 1/2 ETC AT Q21 has NAME*]…
Select only one for each row

*NOTE FOR SCRIPTER – PLEASE SCRIPT AS GRID*

*ROWS:*
1. Been in quarantine for at least one day *Note for scripter to add an (i) button*
*Quarantine is the act of staying at home after a potential exposure to an infected case. If you are in quarantine, you can leave the house, but limit your movements.*
2. Been in isolation for at least one day *Note for scripter to add an (i) button*
*Isolation is the act of separating yourself from people who are not infected, including any household members. You can be in isolation in your house or in a health facility.*
3. Not been to your/their workplace for at least one day due to coronavirus (covid-19) *ASK IF Q3=1,2,3 OR Q25 = 1,2,3*
4. Not attended [university or college] [*ASK IF Q3 = 8 OR Q26 = 3 OR 4*] OR [pre-school or nursery] [*ASK IF Q26 = 1*] OR [school] [*ASK IF Q26 = 2*] OR [*Secondary* school] [AKS IF IF NL OR BE AND *Q26 = 8*]due to coronavirus (covid-19) for at least one day

*COLUMNS:*
1. Yes
2. No
3. Not applicable *ONLY SHOW FOR STATEMENT c)*
4. Don't know
5. Prefer not to answer

*ASK FOR EACH PERSON CODED Q39 1 = 1 (YES) OR Q41 1 = 1 (YES)*
*WRITE IN*
QUAR1

Q42. You said that [you have/NAME has] been in quarantine for at least one day. When did [you/they] start the quarantine?

Please write in

Please use the format “DD/MM"

1. INSERT DATE  USE DATE FORMAT DD/MM.  EARLIEST DATE IS 01/02 AND CANNOT BE A DATE IN THE FUTURE
2. Don’t know
3. Prefer not to answer

*ASK FOR EACH PERSON CODED Q39 1 = 1 (YES) OR Q41 1 = 1 (YES)*
*SA*
QUAR2
Q43. And when did [you/NAME] finish the quarantine?

Select one only

Please use the format “DD/MM"

1. INSERT DATE USE DATE FORMAT DD/MM.  EARLIEST DATE IS 01/02.  DATE MUST NOT BE EARLIER THAN Q42 AND CANNOT BE A DATE IN THE FUTURE
2. I am/they are still in quarantine
3. Don’t know
4. Prefer not to answer

*ASK FOR EACH PERSON CODED Q39 2 = 1 (YES) OR Q41 2 = 1 (YES)*
*WRITE IN*
ISO1
Q44. You said that [you have/NAME has] have been in isolation for at least one day. When did [you/they] start isolating?

Please write in

Please use the format “DD/MM"

1. INSERT DATE USE DATE FORMAT DD/MM.  EARLIEST DATE IS 01/02 AND CANNOT BE A DATE IN THE FUTURE.
2. Don’t know
3. Prefer not to answer

*ASK FOR EACH PERSON CODED Q39 2 = 1 (YES) OR Q41 2 = 1 (YES)*
*SA*
ISO2
Q45. And when did [you/NAME] finish isolating?

Select one only

Please use the format “DD/MM"

1. INSERT DATE  USE DATE FORMAT DD/MM.  EARLIEST DATE IS 01/02.  DATE MUST NOT BE EARLIER THAN Q42 AND CANNOT BE A DATE IN THE FUTURE

2. I am/they are still isolating
3. Don't know
4. Prefer not to answer

*ASK FOR EACH PERSON CODED Q39 3 = 1 (YES) OR Q39 4 = 1 (YES) OR Q40~~1~~ 1 = 1 (YES) OR Q40 2 = 1 (YES)*
*FILTER: (Q39_3 =1 or Q39_4 =1) or (Q40 _1 =1 or Q40 _2 =1)*
*WRITE IN*
CLOSE1
Q46. You said that [your/NAME's] [workplace] [*ASK IF Q3 = 1,2 OR 3 OR Q25=1,2 OR 3*] OR [university or college] [*ASK IF Q3 = 8 OR Q26 = 3 OR 4*] OR [pre-school or nursery] [*ASK IF Q26 = 1*] OR [school] [*ASK IF Q26 = 2*] OR [*Secondary* school] [AKS IF IF NL OR BE AND *Q26 = 8*] was closed due to coronavirus (covid-19) for at least one day in the last seven days. Please select the date when it was first closed.
*IF CODE (Q39 3 AND Q39 4) OR (Q40 1 AND Q40 2) OR (Q41 3 AND Q41 4) ASK QUESTION ONCE FOR UNIVERSITY OR COLLEGE AND ONCE FOR WORKPLACE*

Please write in

Please use the format "DD/MM"

1. INSERT DATE USE DATE FORMAT DD/MM. EARLIEST DATE IS 01/02. 2 AND CANNOT BE A DATE IN THE FUTURE
2. Don't know
3. Prefer not to answer

*ASK FOR EACH PERSON CODED YES (1) AT Q39 3 OR Q39 4 OR Q40 1 or Q40 2. ASK IMMEDIATELY AFTER Q46 FOR EASH PERSON*
*FILTER: (Q39_3 =1 or Q39_4 =1) or (Q40 _1 =1 or Q40 _2 =1*
*WRITE IN*
CLOSE 2
Q47. And when did it open again?

Please write in

Please use the format "DD/MM"

1. INSERT DATE USE DATE FORMAT DD/MM. EARLIEST DATE IS 01/02. DATE MUST NOT BE EARLIER THAN Q42 AND CANNOT BE A DATE IN THE FUTURE
2. It is still closed
3. Don't know
4. Prefer not to answer

*ASK FOR EACH PERSON CODED NO (2) AT Q39 3 OR Q39 4 OR Q40 1 OR Q40 2 <u>AND</u> YES (1) AT Q41 3 or Q41 4*

FILTER: ((Q39_3 =2 or Q40_1=2) AND Q41_3 =1)

OR

((Q39_4 =2 or Q40 _2 =2) AND Q41_4 =1)

*MA*

NOTATTEND1

Q48. You said that [you/NAME] did not attend [the workplace] [*ASK IF Q3 = 1,2 OR 3 OR Q25=1, 2 3 OR 4*] OR [university or college] [*ASK IF Q3 = 8 OR Q26 = 3 OR 4*] OR [pre-school or nursery] [*ASK IF Q26 =1*] OR [school] [*ASK IF Q26 = 2*] OR [*Secondary* school] [AKS IF IF NL OR BE AND *Q26 = 8*] due to coronavirus (covid-19), but it was not closed. For the last seven days, please select the days when [you/NAME] did not go to [the workplace] [*ASK IF Q3 = 1,2 OR 3 OR Q25 = 1,2 OR 3*] OR [university or college] [*ASK IF Q3 = 8 OR Q26 = 3 OR 4*] OR [pre-school or nursery] [*ASK IF Q26 = 1*] OR [school] [*ASK IF Q26 = 2*] OR [*Secondary* school] [AKS IF IF NL OR BE AND *Q26 = 8*] due to coronavirus (covid-19). Only indicate those days where [you/NAME] would normally have gone there.
*IF CODE (Q39 3 AND Q39 4) OR (Q40 1 AND Q40 2) OR (Q41 3 AND Q41 4) ASK QUESTION ONCE FOR UNIVERSITY OR COLLEGE AND ONCE FOR WORKPLACE*
Please tick all that apply

4. SHOW DATES FOR LAST SEVEN DAYS
5. Don't know
6. Prefer not to answer

*ASK FOR EACH PERSON CODED NO (2) AT Q39 3 OR Q39 4 OR Q40 1 OR Q40 2 AND YES (1) AT Q41 3 or Q41 4*
FILTER: ((Q39_3 =2 or Q40_1=2) AND Q41_3 =1)

OR

((Q39_4 =2 or Q40 _2 =2) AND Q41_4 =1)

*MA*

Q48A.

What was your main reason for not attending [the workplace] [*ASK IF Q3 = 1,2 OR 3 OR Q25=1, 2 3 OR 4*] OR [university or college] [*ASK IF Q3 = 8 OR Q26 = 3 OR 4*] OR [pre-school or nursery] [*ASK IF Q26 =1*] OR [school] [*ASK IF Q26 = 2*] OR [*Secondary* school] [AKS IF IF NL OR BE AND *Q26 = 8*]?Please tick all that apply

1. 7 day isolation due to you having symptoms that may be coronavirus

4. 14 day quarantine due to someone else in your household having symptoms that may be coronavirus, or due to contact with a known coronavirus case
5. Other illness (not coronavirus) within the household, (including yourself
6. Caring for someone outside of the household who has been confirmed to have coronavirus (COVID-19)
7. Caring for someone outside of the household who has not been confirmed to have coronavirus (COVID-19)
8. At least one child in my household is home due to school closure
9. Other

*ASK FOR EACH PERSON CODED YES (1) OR NOT APPLICABLE (3) AT Q39 3 OR Q40 1 OR Q41 3*
FILTER: (Q39 _3=1,3) or (Q40_ 1=1,3) or (Q41_ 3=1,3) *MA*
INCOME
Q49. You said that [you/NAME] did not work/visit your workplace for at least one day due to coronavirus (covid-19) or that this was not applicable. Did this have a negative impact on your household income?
Please tick all that apply

1. No, [I/NAME] was able to work from home
2. No, [I/NAME] was able to take carer leave
3. No, but [I/NAME] had to take annual leave
4. No, as [I/NAME] was fully compensated by [my/their] employer
5. No, as [I/NAME] was fully compensated by the government
6. Yes, but [I/NAME] received partial compensation by [my/their] employer
7. Yes, but [I/NAME] received partial compensation by the government
8. Yes, and [I/NAME] received no compensation for [my/their] lost income [SINGLE CODE ONLY]
9. Other (please specify)
10. Don’t know
11. Prefer not to answer

*ASK FOR EACH PERSON CODED (1 OR 2 OR 8 AT Q26) AND (Q39 4 OR Q40 2)*
*MA*
FILTER: any(Q26,1,2,8) and (Q39_4=1 or Q40_2=1)

CHILDCARE
Q50. You said that [NAME’s] [pre-school or nursery] [*ASK IF Q26 = 1*] OR [school] [*ASK IF Q26 = 2*] OR [*Secondary* school] [AKS IF NL OR BE AND *Q26 = 8*]was closed for at least one day due to coronavirus (covid-19). When this happened, who looked after the child/children?
CHILDCARE2
Please tick all that apply

1. A parent, who is unemployed
2. A parent, who was working from home
3. A parent, who works part-time
4. A parent, who took annual leave
5. A parent, who took carer leave
6. A parent, who took unpaid leave
7. A sibling
8. Grandparent(s)
9. A baby sitter, childminder, au pair or nanny (paid)
10. A baby sitter, childminder, au pair or nanny (unpaid)
11. A neighbour, friend, uncle, or aunt
14. People at the school, as my child was eligible for childcare at the school
12. Not required **[PN: Exclusive]**?
13. Other (please specify)

*ASK FOR EACH PERSON CODED 1 or 2 or 8 AT Q26 AND 1 AT q41 4 AND 2 AT Q39_4 AND 2 AT Q40_2*
FILTER: Q26=1,2, 8 and Q41_4 =1 and Q39_4 =2 and Q40_2 =2*MA*

CHILDCARE2
Q51. You said that [NAME] did not attend [pre-school or nursery] [*ASK IF Q26 = 1*] OR [school] [*ASK IF Q26 = 2*] OR [*Secondary* school] [AKS IF IF NL OR BE AND *Q26 = 8*] for at least one day due to coronavirus (covid-19). When this happened, who looked after the child/children?
Please tick all that apply

1. A parent, who is unemployed
2. A parent, who was working from home
3. A parent, who works part-time
4. A parent, who took annual leave
5. A parent, who took carer leave
6. A parent, who took unpaid leave
7. A sibling
8. Grandparent(s)
9. A baby sitter, childminder, au pair or nanny (paid)
10. A baby sitter, childminder, au pair or nanny (unpaid)
11. A neighbour, friend, uncle, or aunt
14. People at the school, as my child was eligible for childcare at the school
12. Not required **[PN: Exclusive]**?
13. Other (please specify)

*ASK ALL*
*SA*
VISIT1
Q52. Did you visit, or intend to visit, any of the following events or locations in the last seven days?
*NOTE FOR SCRIPTER – PLEASE SCRIPT AS GRID*
Select only one for each row

*ROWS:*
a ) Pub, bar or café
b) Restaurant
c) Cinema

e) Supermarket or other shop for food or groceries
f) Sporting event (as attendee), e.g. a football match
g) Sporting event (as participant), e.g. weekly tennis practice
h) Religious gathering
i) Indoor location, where over 100 people were present,
j) Outdoor location, where over 100 people were present

*COLUMNS:*
1. Yes, I visited this event or location
2. I intended to visit but it was cancelled because of the coronavirus (covid-19) epidemic
3. I intended to visit but chose not to go because of the coronavirus (covid-19) epidemic

4. I intended to visit but I had to cancel/it was cancelled for reasons unrelated to the coronavirus (covid-19) epidemic
5. No, I did not visit or intend to visit this event or location

*ASK IF Q52 a)-h) = 1,2 ,3 OR 4 [ASK ALL WHO VISITED OR INTENDED TO VISIT EACH EVENT OR LOCATION. ASK FOR EACH EVENT OR LOCATION AT VISIT1 CODED 1-4*
*WRITE IN*
VISIT2
Q53. You said that you [*ASK IF CODE Q52a)-h) = 1*] visited [*OR IF Q52a)-h) = 2,3 OR 4*] intended to visit [a INSERT EVENT OR LOCATION] *IF Q52a)-H)=2,3 OR 4 ENTER SECOND PART OF STATEMENT FROM Q52* but it was cancelled because of coronavirus / but chose not to go because of coronavirus / but you had to cancel or it was cancelled for reasons unrelated to coronavirus]. How many times did that happen in the last seven days?

Please write in

1. INSERT NUMBER OF TIMES
2. Don't know

**Individual preventive measures**

*ASK ALL*
*SA*
MASK1
Q54. Did you use a face mask yesterday?
Select only one
1. Yes
2. No

*ASK ALL WHO USED A FACE MASK YESTERDAY (Q54=1)*
*WRITE IN*
MASK2
Q55. For how long did you wear a face mask in total?
Provide an approximation of the total duration.

Please write in

INSERT NUMBER OF HOURS
INSERT NUMBER OF MINUTES

*ASK ALL WHO USED A FACE MASK YESTERDAY (Q54=1)*
*MA*
MASK3
Q56. Where did you use your face mask?

1. Everywhere outside my house [*SINGLE CODE*]
2. When walking on the street
3. When cycling
4. On public transport

5. In supermarkets/shops
6. In cinema/bar/restaurant
8. At home
9. At work/school/college/university
7. Other (please specify)

*ASK ALL*
*WRITE IN*
WASH
Q57. How many times did you wash your hands with soap in the last three hours?

Please write in

INSERT NUMBER OF TIMES (RANGE FROM 0-25)

*ASK ALL*
*WRITE IN*
SANIT
Q58. How many times did you use hand sanitizer in the last three hours?

Please write in

INSERT NUMBER OF TIMES (RANGE FROM 0-25)

*ASK ALL*

*MA*

TRANSPORT

Q59. Did you travel on any public transport yesterday?

Please tick all that apply
1. No *[SINGLE CODE]*
2. Train/tube
3. Bus/tram
4. Taxi, Uber, or similar ride-hailing app
5. Aeroplane

*ASK ALL WHO TRAVELLED ON PUBLIC TRANSPORT (Q59 = CODES 2-5, FOR EACH TYPE)*
*WRITE IN*
TRANSDUR
Q60. And approximately how long did you spend on the [*INSERT FROM Q59*] in total?

Please write in

INSERT NUMBER OF HOURS
INSERT NUMBER OF MINUTES

**Contact survey**

*ASK ALL*
*INTRO SCREEN*
We will now ask you to remember who you have been in contact with yesterday, between 5am yesterday and 5am today.

These questions are voluntary but they are really important in helping us understand the spread of COVID-19 and the impact of different public health interventions. It will not be possible to identify you or any member of your household in the published findings.

We are only interested in direct contacts, which are **people who you met in person** and with whom you exchanged at least a few words, or with whom you had physical contact (e.g. a handshake, embracing, kissing, contact sports).
**Note that if you only spoke to someone over the phone or internet, they should not be included in this section.**

*ASK ALL*
*SA*
CONTACT1
Q62. Which of the following people did you have direct contact with in person, between 5am yesterday and 5am today, in person?
*NOTE FOR SCRIPTER – PLEASE SCRIPT AS GRID*

*Note for scripter to add (i) button for 'direct contact':*
*We are only interested in direct contacts, which are people who you met in person and with whom you exchanged at least a few words, or with whom you had physical contact (e.g. a handshake, embracing, kissing, contact sports).*
***Note that if you only spoke to someone over the phone or internet, they should not be included in this section.***

*ROWS:*
*INSERT ALL NAMES FROM Q21*

*COLUMNS:*
1. Yes
2. No

*ASK ALL*
*WRITE IN*
CONTACT2

Q63. And what **other people, outside of your household**, did you have direct contact with in person, between 5am yesterday and 5am today? This could include friends, family, work colleagues, or people you spoke to in shops and so on.

Please write the nickname of each person below. Note that this nickname is only needed to make it easier for you to complete the survey, so please pick a nickname that is easy to remember. Your individual responses will not be shared with anyone outside this survey.

*We are only interested in people who you met in person and* ***with whom you exchanged at least a few words, or with whom you had physical contact*** *(e.g. a handshake, embracing, contact sports)*.

It is easiest to list names in chronological order, e.g. After I had breakfast at home, I went to work where I met with Jack, Deborah, and two clients. On my way back home, I chatted with the shop assistant at the petrol station (give a nickname like “shop assistant”). When I returned home, I accepted a package from the delivery person, and I spoke to my friend, Fatima, in my garden. Etc.

**Please do not list yourself, anyone that you listed as being a part of your household, or anyone who you only spoke to over the phone or internet.**

*PN: SHOW IF Q20 IS NOT NONE OR IF NO-ONE IS SELECTED AT Q62]*
*You have already indicated contact with the following household members, add additional contacts in the textboxes below:*

LIST HOUSEHOLD MEMBERS SELECTED AT Q62

*ALLOW BOXES TO INPUT NICKNAMES PN SHOW 100 BOXES ON SCREEN*

*PN AT TOP OF PAGE WITH TEXT BOXES INCLUDE THE FOLLOWING.*

*PLEASE LIST ALL OTHER CONTACTS YOU HAVE OUTSIDE OF YOUR HOUSEHOLDE, FOR EXAMPLE, JACK, DEBORAH, CLIENT 1, CLIENT 2, SHOP ASSISTANT, DELIVERY PERSON, FATIMA*

*No-one/No other people [PN. ALLOW THIS TO BE SELECTED WHEN TEXT BOXES COMPLETED]*

*ASK THE FOLLOWING LOOP Q69 TO Q72 TO TIME FOR EACH NAME GIVEN AT Q62*
*ASK THE FOLLOWING LOOP Q66 TO Q72 TO TIME FOR EACH NAME GIVEN AT Q63*

*ASK FOR EACH PERSON AT Q63*
*SA*

Q66. Which of the following age groups does NAME fit into? Please give an estimate if you are not sure
Select only one

1. Under 1
2. 1-4
3. 5-9
4. 10-14
5. 15-19

6. 20-24
7. 25-34
8. 35-44
9. 45-54
10. 55-64
11. 65-69
12. 70-74
13. 75-79
14. 80-84
15. 85+
16. Don’t know
17. Prefer not to answer
18. This person is me

*ASK ALL WHERE NAME GIVEN AT Q63 AND Q66 IS NOT EQUAL TO CODE 18*
*SA*
CONTACTGEN
Q67. As far as you know, which of the following describes how [NAME] thinks of themselves?
Select only one

1. _1 Male
2. _2 Female
3. _3 In another way
4. _4 Prefer not to answer
5. _5 Don’t know

*ASK ALL WHERE NAME GIVEN AT Q63 AND Q66 IS NOT EQUAL TO CODE 18*
*SA*
CONTACTRELAT
Q68. What is [NAME]’s relationship to you?
Select only one
1. They are a family member who is not in my household
2. They are someone I work with
3. They are someone I go to school, college or university with
4. They are a friend
5. Other
6. Prefer not to answer

*ASK ALL WHERE NAME GIVEN AT Q62 OR Q63 AND Q66 IS NOT EQUAL TO CODE 18*
*SA*
CONTACTFREQ
Q69. Before the coronavirus epidemic started, how often did you usually have direct contact with [*NAME*]?
A direct contact is when you **meet with this person in person** and when you exchange at least a few words, or when you have physical contact (e.g. handshake, embracing, kissing, contact sports).
**Please do not include times that you speak to them over the phone or internet.**
Select only one for each person

1. Every day or almost every day

2. About once or twice a week
3. Every 2-3 weeks
4. About once per month
5. Less often than once per month
6. Never met them before
7. Prefer not to answer

*ASK ALL WHERE NAME GIVEN AT Q62 OR Q63 AND Q66 IS NOT EQUAL TO CODE 18*
*SA*
PHYS
Q70. When you had direct contact with [NAME] yesterday, did you have…?
Select only one

1. Physical contact (any sort of skin-to-skin contact such as e.g. hand shaking, embracing or kissing)
2. Non-physical contact (you did not touch the person)
3. Prefer not to answer

*ASK ALL WHERE NAME GIVEN AT Q62 OR Q63 AND Q66 IS NOT EQUAL TO CODE 18*
*MA*
WHERE
Q71. And where did you have direct contact with [NAME]?
Please tick all that apply

1. At home (including at your door, in your garden, and within entrances to your home such as stairways, lifts, and corridors)
2. At someone else's house
3. At work
4. At a place of worship
5. On any form of transport
6. At university, school, pre-school, or nursery
7. At a shop for essentials, eg a supermarket, grocery store, market, pharmacist, or bicycle shop
8. At a shop for non--essential items, eg a gardening center, or a clothes, electronics, furniture, or DIY shop
9. At a place of entertainment such as a restaurant, bar, cinema
10. At a place for sports such as a gym or sports club/match [PN. DO NOT SHOW IN BE AND NL]
11. Outside, for example in a park, on the street or in the countryside
14. In a healthcare setting, eg hospital, GP, A&E, outpatient facility, dentist, physiotherapist, optometrist, etc

15. At a hair dresser, barber, nail salon, beauty parlor or similar location

12. Somewhere else (please specify)
13. At an open air market [PN: SHOW IN BE AND NL ONLY.  SHOW AFTER OPTION 8]

*ASK ALL WHERE NAME GIVEN AT Q62 OR Q63 AND Q66 IS NOT EQUAL TO CODE 18*
*WRITE IN*
TIME
Q72. Please estimate the total amount of time you spent with [NAME] in person yesterday.

Please write in

*INSERT HOURS*
*INSERT MINUTES*

ASK If COUNTRY = UK AND Q71= 1, 2, 3, 4, 6, 7, 9, 10 or 12, 14 or 15
IF COUNTRY – BE OR NL ASK ALL WHERE NAME GIVEN AT Q62 OR Q63
Q73 Was the time you spent with [NAME] yesterday inside or outside?
Please tick all that apply
Inside
Outside

### Covid 19 behaviour and contact survey

**QCOUNTRY**

1. UK

**CAPTURE DATA FROM IMPORT:**

**Qsample=1 WAVE-TO-WAVE**
**Qsample=2 FRESH SAMPLE**

**Panel**
**3. Panel C**
**4. Panel D**

**Wave**

1. **Wave 1**
2. **Wave 2**
3. **Wave 3**
4. **Wave 4**
5. **Wave 5**
6. **Wave 6**

**Demographics**

*[Standard Screener: DO NOT MODIFY OR TRANSLATE]*
RESP_AGE [Hidden].
*ASK ALL*
*SA*
What is your age in years?
*YEAR/MONTH*

Hidden Question - RESP_AGE "this is a dummy question that will hold age"
USE RESP_AGE [Hidden] response list

1. _18 "18",
2. _19 "19",
3. _20 "20",
4. _21 "21",
5. _22 "22",
6. _23 "23",
7. _24 "24",
8. _25 "25",
9. . . .
10. _65 "65",
11. _66 "66",

12. _67 "67",
13. _68 "68",
14. _69 "69",
15. _70 "70",
16. _71 "71",
17. _72 "72",
18. _73 "73",
19. _74 "74",
20. _75 "75",
21. _76 "76",
22. _77 "77",
23. _78 "78",
24. _79 "79",
25. _80 "80",
26. _81 "81",
27. _82 "82",
28. _83 "83",
29. _84 "84",
30. _85 "85",
31. _86 "86",
32. _87 "87",
33. _88 "88",
34. _89 "89",
35. _90 "90",
36. _91 "91",
37. _92 "92",
38. _93 "93",
39. _94 "94",
40. _95 "95",
41. _96 "96",
42. _97 "97",
43. _98 "98",
44. _99 "99",
45. _100 "100",
46. _101 "101",
47. _102 "102",
48. _103 "103",
49. _104 "104",
50. _105 "105",
51. _998 "Age not calculated",
52. _999 "Age < 1"

*TERMINATE IS RESP_AGE < 18 YO*

*RECODE QUOTAGERANGE*

| |
|---|
| 18-24 |
| 25-34 |

| |
|---|
| 35-44 |
| 45-54 |
| 55-64 |
| 65+ |

*SA*
*[Standard Screener: DO NOT MODIFY OR TRANSLATE]*
[PN: STOP_REALLOCATION if GENDER_NONBINARY is 3 or 4, and RESP_GENDER was not answered in a previous survey]
GENDER_NONBINARY. Which of the following describes how you think of yourself?
Select only one

1. _1 Male
2. _2 Female
3. _3 In another way
4. _4 Prefer not to answer

*[Standard Screener: DO NOT MODIFY OR TRANSLATE]*
*IF Qsample=1 HIDE AND IMPORT DATA, CREATE RECODE VARIABLE WITH THE NAME QMktSize_GB*

*IF Qsample=2 ASK QMktSize_GB*

[PN: If Prefer not to answer is selected, ask UKREGION1]

QMktSize_GB. Where do you live? Please note: This question may be considered personal. We would like to remind you that your participation is strictly voluntary and that your responses are used for research purposes only. The answers that you provide will be presented in aggregate form and none of them will be linked back to you in any way. All data will be collected and processed in adherence to the Market Research Society's Code of Conduct and the General Data Protection Regulation (GDPR).

| |
|---|
| **Postcode** |
| **Postal Town** |
| **Prefer Not to Answer** |

*IF Qsample=1 ASK*

*HHCOMPCONFRIM In the last wave of research these are the people that you said live in your household. Please could you confirm that this is correct?*
*LIST ALL HOUSEHOLD MEMBERS LISTED AT Q21 FROM PREVIOUS WAVE*
*In case there are no respondents captured from import code 2 will not be shown.*

1. *Yes, this is correct (EXCLUSIVE)*
2. *No, one or more of these people no longer live in my household*
3. *No, there are one or more new people now living in my household*

*IF HHCOMPCONFIRM=2 ASK*

*HHCOMPREMOVE Please indicate which of these people **no longer** live in your household?*
*LIST ALL HOUSEHOLD MEMBERS AT Q21 FROM PREVIOUS WAVE AND ALLOW SELECTION OF THOSE WHO NO LONGER LIVE IN THE HOUSEHOLD.*

*IF HHCOMPCONFIRM=3 ASK*
*HHCOMPADD Please write in below the nicknames of the new people who now live in your household?*
*INSERT TEXTBOXES ONE AT A TIME TO RECORD NICKAMES OF NEW PEOPLE – ADD UP TO 4 TEXT BOXES*

*IF Qsample=1 HIDE AND IMPORT DATA, CREATE RECODE VARIABLE WITH THE NAME Q20.*
*IF Qsample=2 ASK Q20*

*ASK ALL*
*SA*
QHOUSE
Q20. Not including you, how many other people live in your household? By household, we mean anyone living at the same address as you, that you share a kitchen with.
Select only one

None
1
2
3
4
5
6
7
8
9
10
11 or more

*IF Q20=None, THANK AND CLOSE*

*ASK IF Q20=1 OR MORE (I.E. IF NOT A SINGLE PERSON HOUSEHOLD) ) OR HHCOMPCONFIRM <>1*

*SA*
Q20a . Which of the following best describes your household?
Select only one

2. Two or more non-family adults
3. Couple with dependent children aged 0-17
4. Couple with independent children only
5. Couple with no children

6. Lone parent with dependent children aged 0-17
7. Lone parent with independent children only
8. Households containing two or more families with children aged 0-17
9. Other

*IF Q20a=3,6,8 CONTINUE*
*IF Q2a=2,4,5,7,9 THANK AND CLOSE*

*ASK IF Q20a=3,6,8*
*SA*
Q20b. And are you the parent of one or more of these children aged 0-17?
Select only one

Yes
No

*IF Q20b=1 CONTINUE*
*IF Q20b=2 THANK AND CLOSE*

**MAIN INTRO SCREEN**

Thank you for agreeing to take part in this important research about the new coronavirus (COVID-19) pandemic. Please take the time to carefully read the information below and the detailed survey participation notice.

The survey is being run by Ipsos on behalf of a group of experts in mathematical and statistical modelling of infectious diseases and public health – London School of Hygiene and Tropical Medicine (LSHTM; United Kingdom).

The research is part of a larger programme of work funded by the UK Medical Research Council which aims to provide urgently needed answers about the epidemiological characteristics of COVID-19, the social dynamics of the outbreak, and the related public health preparedness and response to the ongoing pandemic, as well as to assess its economic impact. It will be used directly on active communication and interaction with policy makers/authorities, other scientific groups and the general public, to help minimise the COVID-19's public health, economic and social impact.

Taking part in this survey is completely voluntary, and you may refuse to do so. However, we would really value your support as the findings will provide invaluable information for decision making and strategies developed in the near future.

*NEW SCREEN*
**Ethics form for participants**

I confirm that I am 18 years or older.

I confirm that I have read and understood the information for this study. I have had the opportunity to consider the information before taking part in this study.

I understand that taking part is voluntary, and that I am free to withdraw at any time without giving any reason and without any of my rights being affected.

I understand that relevant sections of anonymized data collected during the study may be looked at by authorised individuals from the University of Antwerp, University of Hasselt, London School of Hygiene & Tropical Medicine, National Institute for Public Health and Environment in the Netherlands, University of Bern, and the ISI Foundation Turin.

I understand that ANONYMIZED data about me may be shared via a public data repository and that I and any of my household members or contacts will not be identifiable from this information.

I understand that this is a longitudinal study, and that I will be invited to complete multiple questionnaires over the course of the ongoing epidemic.

1. Agree to take part
2. Do not agree to take part

*SA*

Q3. What is your current employment status?

Select only one

2. _1 Employed full-time (34 hours or more)
3. _2 Employed part-time (less than 34 hours)
4. _3 Self employed
5. _4 Unemployed but looking for a job
6. _5 Unemployed and not looking for a job
7. _6 Full-time parent, homemaker
8. _7 Retired
9. _8 Student/Pupil
10. _9 Long-term sick or disabled

*ASK IN UK ONY*

*[Standard Screener: DO NOT MODIFY OR TRANSLATE]*

[To get UK01SG, need to ask EU01HINC. If EU01HINC=1,2, then ask UK01OCCR. If EU01HINC=3, ask UK01OCCHI. UK01SG is computed in the same manner as in the IIS panel. Details about that on panelstats.]

*[Standard Screener: DO NOT MODIFY OR TRANSLATE]*

[PN: if HHCMP10=1, then do not ask EU01HINC Q4 and assume EU01HINC Q4=_1]

*ASK IF IN UK AND EU01HINC=1 OR 2*

*[Standard Screener: DO NOT MODIFY OR TRANSLATE]*

*IF Qsample=1 HIDE AND IMPORT DATA, CREATE RECODE VARIABLE WITH THE NAME UK01OCCR.*
*IF Qsample=2 ASK UK01OCCR.*

*SA*
*UK01OCCR.*

In which of the below categories does your occupation fall? If retired or unemployed, please indicate the category closest to your previous occupation.

Select only one

1. USE UK01OCCR response list
2. Legislators, senior officials and managers [EXPANDABLE HEADER]
   a. _1100 Legislators and senior officials
3. Corporate managers [EXPANDABLE HEADER]
   a. _1210 Directors and chief executives
4. Production and operations department managers [EXPANDABLE HEADER]
   a. _1221 Production and operations department managers in agriculture, hunting, forestry and fishing
   b. _1222 Production and operations department managers in manufacturing
   c. _1223 Production and operations department managers in construction
   d. _1224 Production and operations department managers in wholesale and retail trade
   e. _1225 Production and operations department managers in restaurants and hotels
   f. _1226 Production and operations department managers in transport, storage and communications
   g. _1227 Production and operations department managers in business services
   h. _1228 Production and operations department managers in personal care, cleaning and related services
   i. _1229 Production and operations department managers not elsewhere classified
5. Other department managers [EXPANDABLE HEADER]
   a. _1231 Finance and administration department managers
   b. _1232 Personnel and industrial relations department managers
   c. _1233 Sales and marketing department managers
   d. _1234 Advertising and public relations department managers
   e. _1235 Supply and distribution department managers
   f. _1236 Computing services department managers
   g. _1237 Research and development department managers
   h. _1239 Other department managers not elsewhere classified
6. General managers [EXPANDABLE HEADER]
   a. _1311 General managers in agriculture, hunting, forestry/ and fishing
   b. _1312 General managers in manufacturing
   c. _1313 General managers in construction
   d. _1314 General managers in wholesale and retail trade
   e. _1315 General managers of restaurants and hotels
   f. _1316 General managers in transport, storage and communications
   g. _1317 General managers of business services
   h. _1318 General managers in personal care, cleaning and related services
   i. _1319 General managers not elsewhere classified
7. Physical, mathematical and engineering science professionals [EXPANDABLE HEADER]

   a. _2110 Physicists, chemists and related professionals
   b. _2120 Mathematicians, statisticians and related professionals
   c. _2130 Computing professionals
8. Architects, engineers and related professionals [EXPANDABLE HEADER]
   a. _2141 Architects, town and traffic planners
   b. _2142 Civil engineers
   c. _2143 Electrical engineers
   d. _2144 Electronics and telecommunications engineers
   e. _2145 Mechanical engineers
   f. _2146 Chemical engineers
   g. _2147 Mining engineers, metallurgists and related professionals
   h. _2148 Cartographers and surveyors
   i. _2149 Architects, engineers and related professionals not elsewhere classified
9. Life science and health professionals [EXPANDABLE HEADER]
   a. _2210 Life science professionals
   b. _2220 Health professionals (except nursing)
   c. _2230 Nursing and midwifery professionals
   d. _2300 Teaching professionals
10. Other professionals [EXPANDABLE HEADER]
   a. _2410 Business professionals
   b. _2420 Legal professionals
   c. _2430 Archivists, librarians and related information professionals
   d. _2440 Social science and related professionals
   e. _2450 Writers and creative or performing artists
   f. _2460 Religious professionals
11. Physical and engineering science associate professionals [EXPANDABLE HEADER]
   a. _3110 Physical and engineering science technicians
   b. _3120 Computer associate professionals
   c. _3130 Optical and electronic equipment operators
   d. _3140 Ship and aircraft controllers and technicians
   e. _3150 Safety and quality inspectors
   f. _3200 Life science and health associate professionals
   g. _3300 Teaching associate professionals
12. Other associate professionals [EXPANDABLE HEADER]
   a. _3410 Finance and sales associate professionals
   b. _3420 Business services agents and trade brokers
   c. _3430 Administrative associate professionals
   d. _3440 Customs, tax and related government associate professionals
   e. _3450 Police inspectors and detectives
   f. _3460 Social work associate professionals
   g. _3470 Artistic, entertainment and sports associate professionals
   h. _3480 Religious associate professionals
13. Clerks [EXPANDABLE HEADER]
   a. _4100 Office clerks
   b. _4200 Customer services clerks
14. Personal and protective services workers [EXPANDABLE HEADER]
   a. _5110 Travel attendants and related workers
   b. _5120 Housekeeping and restaurant services workers

c. _5130 Personal care and related workers
d. _5140 Other personal services workers
e. _5160 Protective services workers
f. _5200 Models, salespersons and demonstrators

15. Skilled agricultural and fishery workers [EXPANDABLE HEADER]
    a. _6000 Skilled agricultural and fishery workers
16. Craft and related trades workers [EXPANDABLE HEADER]
    a. _7100 Extraction and building trades workers
17. Metal, machinery and related trades workers [EXPANDABLE HEADER]
    a. _7210 Metal moulders, welders, sheet-metal workers, structural - metal preparers, and related trades workers
    b. _7220 Blacksmiths, tool-makers and related trades workers
    c. _7230 Machinery mechanics and fitters
    d. _7240 Electrical and electronic equipment mechanics and fitters
18. Precision, handicraft, printing and related trades workers [EXPANDABLE HEADER]
    a. _7310 Precision workers in metal and related materials
    b. _7320 Potters, glass-makers and related trades workers
    c. _7330 Handicraft workers in wood, textile, leather and related materials
    d. _7340 Printing and related trades workers
19. Other craft and related trades workers [EXPANDABLE HEADER]
    a. _7410 Food processing and related trades workers
    b. _7420 Wood treaters, cabinet-makers and related trades workers
    c. _7430 Textile, garment and related trades workers
    d. _7440 Pelt, leather and shoemaking trades workers
20. Plant and machine operators and assemblers [EXPANDABLE HEADER]
    a. _8000 Plant and machine operators and assemblers
21. Elementary occupations [EXPANDABLE HEADER]
    a. _9100 Sales and services elementary occupations
    b. _9200 Agricultural, fishery and related labourers
    c. _9300 Labourers in mining, construction, manufacturing and transport
22. Armed forces [EXPANDABLE HEADER]
    a. _9888 Armed forces
23. Did not work before [EXPANDABLE HEADER]
    a. _9991 Unemployed and not looking for a job / Long-term sick or disabled
    b. _9992 Pupil /Student/ in full time education
    c. _9993 Housewife

*ASK IF IN UK AND EU01HINC =3*

*[Standard Screener: DO NOT MODIFY OR TRANSLATE]*

[PN: Part of the occupation module, cannot be asked independent. Asked only if EU01HINC Q4=3. When respondent is also main earner (EU01HINC Q4=1 or 2), then values recorded in UK01OCCR are automatically transferred to UK01OCCHI. ]

*IF Qsample=1 HIDE AND IMPORT DATA, CREATE RECODE VARIABLE WITH THE NAME UK01OCCHI.*
*IF Qsample=2 ASK UK01OCCHI.*

*SA*

*UK01OCCHI.*

What is the occupation of the person with the highest income? If retired or unemployed, please indicate the category closest to his/her previous occupation.

Select only one

1. USE UK01OCCHI response list
2. Legislators, senior officials and managers [EXPANDABLE HEADER]
    a. _1100 Legislators and senior officials
3. Corporate managers [EXPANDABLE HEADER]
    a. _1210 Directors and chief executives
4. Production and operations department managers [EXPANDABLE HEADER]
    a. _1221 Production and operations department managers in agriculture, hunting, forestry and fishing
    b. _1222 Production and operations department managers in manufacturing
    c. _1223 Production and operations department managers in construction
    d. _1224 Production and operations department managers in wholesale and retail trade
    e. _1225 Production and operations department managers in restaurants and hotels
    f. _1226 Production and operations department managers in transport, storage and communications
    g. _1227 Production and operations department managers in business services
    h. _1228 Production and operations department managers in personal care, cleaning and related services
    i. _1229 Production and operations department managers not elsewhere classified
5. Other department managers [EXPANDABLE HEADER]
    a. _1231 Finance and administration department managers
    b. _1232 Personnel and industrial relations department managers
    c. _1233 Sales and marketing department managers
    d. _1234 Advertising and public relations department managers
    e. _1235 Supply and distribution department managers
    f. _1236 Computing services department managers
    g. _1237 Research and development department managers
    h. _1239 Other department managers not elsewhere classified
6. General managers [EXPANDABLE HEADER]
    a. _1311 General managers in agriculture, hunting, forestry/ and fishing
    b. _1312 General managers in manufacturing
    c. _1313 General managers in construction
    d. _1314 General managers in wholesale and retail trade
    e. _1315 General managers of restaurants and hotels
    f. _1316 General managers in transport, storage and communications
    g. _1317 General managers of business services
    h. _1318 General managers in personal care, cleaning and related services
    i. _1319 General managers not elsewhere classified
7. Physical, mathematical and engineering science professionals [EXPANDABLE HEADER]
    a. _2110 Physicists, chemists and related professionals
    b. _2120 Mathematicians, statisticians and related professionals
    c. _2130 Computing professionals
8. Architects, engineers and related professionals [EXPANDABLE HEADER]

    a. _2141 Architects, town and traffic planners
    b. _2142 Civil engineers
    c. _2143 Electrical engineers
    d. _2144 Electronics and telecommunications engineers
    e. _2145 Mechanical engineers
    f. _2146 Chemical engineers
    g. _2147 Mining engineers, metallurgists and related professionals
    h. _2148 Cartographers and surveyors
    i. _2149 Architects, engineers and related professionals not elsewhere classified
9. Life science and health professionals [EXPANDABLE HEADER]
    a. _2210 Life science professionals
    b. _2220 Health professionals (except nursing)
    c. _2230 Nursing and midwifery professionals
    d. _2300 Teaching professionals
10. Other professionals [EXPANDABLE HEADER]
    a. _2410 Business professionals
    b. _2420 Legal professionals
    c. _2430 Archivists, librarians and related information professionals
    d. _2440 Social science and related professionals
    e. _2450 Writers and creative or performing artists
    f. _2460 Religious professionals
11. Physical and engineering science associate professionals [EXPANDABLE HEADER]
    a. _3110 Physical and engineering science technicians
    b. _3120 Computer associate professionals
    c. _3130 Optical and electronic equipment operators
    d. _3140 Ship and aircraft controllers and technicians
    e. _3150 Safety and quality inspectors
    f. _3200 Life science and health associate professionals
    g. _3300 Teaching associate professionals
12. Other associate professionals [EXPANDABLE HEADER]
    a. _3410 Finance and sales associate professionals
    b. _3420 Business services agents and trade brokers
    c. _3430 Administrative associate professionals
    d. _3440 Customs, tax and related government associate professionals
    e. _3450 Police inspectors and detectives
    f. _3460 Social work associate professionals
    g. _3470 Artistic, entertainment and sports associate professionals
    h. _3480 Religious associate professionals
13. Clerks [EXPANDABLE HEADER]
    a. _4100 Office clerks
    b. _4200 Customer services clerks
14. Personal and protective services workers [EXPANDABLE HEADER]
    a. _5110 Travel attendants and related workers
    b. _5120 Housekeeping and restaurant services workers
    c. _5130 Personal care and related workers
    d. _5140 Other personal services workers
    e. _5160 Protective services workers
    f. _5200 Models, salespersons and demonstrators

15. Skilled agricultural and fishery workers [EXPANDABLE HEADER]
    a. _6000 Skilled agricultural and fishery workers
16. Craft and related trades workers [EXPANDABLE HEADER]
    a. _7100 Extraction and building trades workers
17. Metal, machinery and related trades workers [EXPANDABLE HEADER]
    a. _7210 Metal moulders, welders, sheet-metal workers, structural - metal preparers, and related trades workers
    b. _7220 Blacksmiths, tool-makers and related trades workers
    c. _7230 Machinery mechanics and fitters
    d. _7240 Electrical and electronic equipment mechanics and fitters
18. Precision, handicraft, printing and related trades workers [EXPANDABLE HEADER]
    a. _7310 Precision workers in metal and related materials
    b. _7320 Potters, glass-makers and related trades workers
    c. _7330 Handicraft workers in wood, textile, leather and related materials
    d. _7340 Printing and related trades workers
19. Other craft and related trades workers [EXPANDABLE HEADER]
    a. _7410 Food processing and related trades workers
    b. _7420 Wood treaters, cabinet-makers and related trades workers
    c. _7430 Textile, garment and related trades workers
    d. _7440 Pelt, leather and shoemaking trades workers
20. Plant and machine operators and assemblers [EXPANDABLE HEADER]
    a. _8000 Plant and machine operators and assemblers
21. Elementary occupations [EXPANDABLE HEADER]
    a. _9100 Sales and services elementary occupations
    b. _9200 Agricultural, fishery and related labourers
    c. _9300 Labourers in mining, construction, manufacturing and transport
22. Armed forces [EXPANDABLE HEADER]
    a. _9888 Armed forces
23. Did not work before [EXPANDABLE HEADER]
    a. _9991 Unemployed and not looking for a job / Long-term sick or disabled
    b. _9992 Pupil /Student/ in full time education
    c. _9993 Housewife

UK01SG [Hidden]. Hidden Question:

Social Grade

1. _1 A - Upper middle class
2. _2 B - Middle class
3. _3 C1 - Lower middle class
4. _4 C2 - Skilled working class
5. _5 D - Working class
6. _6 E - Lower level of subsistence

*ASK ALL*
*SA*
*[Standard Screener: DO NOT MODIFY OR TRANSLATE]*
*IF Qsample=1 HIDE AND IMPORT DATA, CREATE RECODE VARIABLE WITH THE NAME INCOMEUK.*
*IF Qsample=2 ASK INCOMEUK.*

INCOMEUK.

The next question may be considered personal, but it is not mandatory to answer. If you do, we assure you that your responses will be kept strictly confidential and used for research purposes only. What is the COMBINED TOTAL ANNUAL INCOME (pre-tax, pensions, or national insurance deductions) earned by all members of your family? Here, family is defined as everyone related by marriage, civil partnership, birth, or adoption living at the same address. Please include all your income sources: salaries, scholarships, pension and Social Security benefits, dividends from shares, income from rental properties, child support and alimony etc.

Select only one

1. _1 Under £5,000
2. _2 £5,000 - £9,999
3. _3 £10,000 - £14,999
4. _4 £15,000 - £19,999
5. _5 £20,000 - £24,999
6. _6 £25,000 - £34,999
7. _7 £35,000 - £44,999
8. _8 £45,000 - £54,999
9. _9 £55,000 - £64,999
10. _9 £65,000 - £99,999
11. _10 £100,000 or more
12. _11 Prefer not to answer

*IF QSAMPLE=1 1 HIDE AND IMPORT DATA, CREATE RECODE VARIABLE WITH THE NAME Q21IMPORT*
*IF QSAMPLE=2 AND Q20=1 OR MORE HAVE ONE ROW FOR EACH PERSON IN QHOUSE*
*LIMIT TO 11 ANSWER BOXES IF QHOUSE=11*
*WRITE IN*

QNAME

Q21. Please write the nickname of each other person in your household.

Note that this nickname is only needed to make it easier for you to complete the survey, so please pick a nickname that will help you identify each household member later in the questionnaire. Nicknames are not visible to anyone outside of this survey.

Please write in

1. NAME1
2. NAME2
3. NAME3

etc.

*ASK THE FOLLOWING LOOP Q23 TO Q27 FOR EACH NAME GIVEN AT Q21 Q21IMPORT (EXCLUDE NAMES SELECTED AT HHCOMPRREMOVE) AND EACH OF NAMES GIVEN AT HHCOMPADD*

*SA*

QHHAGE

Q23. Which of the following age groups do they fit into?

1. Under 1
2. 1-4
3. 5-11
4. 12-174
5. 18-19
6. 20-24
7. 25-34
8. 35-44
9. 45-54
10. 55-64
11. 65-69
12. 70-74
13. 75-79
14. 80-84
15. 85+
16. Don't know
17. Prefer not to answer

*IF QHHAGE DOES NOT EQUAL 1, 2, 3 O4 4 FOR ANY NAME GIVEN AT Q21, THANK AND CLOSE*

*IF Qsample=1 AND HHCOMPCONFIRM=1 HIDE AND IMPORT DATA, CREATE RECODE VARIABLE WITH THE NAME QHHGENDERI*
*IF Qsample=1 AND HHCOMPCONFIRM=3 ASK FOR EACH NAME AT HHCOMPADD ASK QHHGENDER*

IF Qsample=2 ASK QHHGENDER

*SA*
QHHGENDER
Q24. As far as you know, which of the following describes how [NAME] thinks of themselves?
Select only one

1. _1 Male
2. _2 Female
3. _3 In another way
4. _4 Prefer not to answer
5. _5 Don't know

*ONLY ASK THOSE AGED 16 AND OVER AT QHHAGE. IF QSAMPLE=1 EXCLUDE NAMES SELECTED AT HHCOMPREMOVE and ask each of names given at hhcompadd*
*SA*
Q25. What is [NAME]'s current employment status?
Select only one

1. _1 Employed full-time (34 hours or more)
2. _2 Employed part-time (less than 34 hours)
3. _3 Self employed
4. _4 Unemployed but looking for a job
5. _5 Unemployed and not looking for a job

6. _6 Full-time parent, homemaker
7. _7 Retired
8. _8 Student/Pupil
9. _9 Long-term sick or disabled

*SA*

IF QSAMPLE=1 EXCLUDE NAMES SELECTED AT HHCOMPREMOVE and ask each of names given at hhcompadd

QHHATTEND

Q26. Does [NAME] attend any of the following as a pupil or student?

Select only one

1. Nursery or pre-school [*UK ONLY SHOW THIS OPTION FOR THOSE AGED 4 OR UNDER AT QHHAGE]*
2. School [*UK ONLY SHOW THIS OPTION FOR THOSE AGED 4 TO 21 AT QHHAGE] [BE AND NL SHOW THIS OPTION FOR THOSE AGED 4-14 ONLY]*
3. Further education, e.g. college [*UK ONLY SHOW THIS OPTION FOR THOSE AGED 14 AND OVER AT QHHAGE] [DO NOT SHOW FOR BE AND NL]*
4. Higher education, e.g. university [*UK, BE AND NL SHOW THIS OPTION FOR THOSE AGED 14 AND OVER AT QHHAGE]*
5. None of the above
6. Don't know
7. Prefer not to answer

*ASK ALL*
*SA*
FLU
Q28. Are you or any other household member in a high-risk group under which the annual influenza vaccine would usually be offered by the NHS?
High risk groups include individuals with: chronic respiratory disease, chronic heart disease, chronic kidney disease, chronic liver disease, chronic neurological disease, diabetes (all types), immunosuppression (due to disease or treatment), asplenia or dysfunction of the spleen, class III obesity (BMI ≥ 40), and pregnant women.

Select only one for each row

*ASK FOR SELF AND EACH PERSON NAMED AT Q21/Q21 IMPORT. IF QSAMPLE=1 AND HHCOMPCONFRIM =2 THEN REMOVE ANYONE SELECTED AT HHCOMPREMOVE. QSAMPLE=1 AND HHCOMPCONFRIM =3 THEN ADD PEOPLE FROM HHCOMPADD*

*NOTE FOR SCRIPTER – PLEASE SCRIPT AS GRID*

*ROWS:*
0. Yourself
1. Name 1
2. Name 2
etc.

*COLUMNS:*
1. Yes
2. No
3. Don't know
4. Prefer not to answer

**Symptoms**
*ASK ALL*
*MA*
SYMPTOMS
Q29. Have you, or anyone else in your household, had any of the following symptoms in the last seven days?
Please tick all that apply for each row
*ASK FOR SELF AND EACH PERSON NAMED AT Q21/Q21 IMPORT. IF QSAMPLE=1 AND HHCOMPCONFRIM =2 THEN REMOVE ANYONE SELECTED AT HHCOMPREMOVE. QSAMPLE=1 AND HHCOMPCONFRIM =3 THEN ADD PEOPLE FROM HHCOMPADD*

*NOTE FOR SCRIPTER – PLEASE SCRIPT AS GRID – ROTATE COLUMNS 1-7*

*ROWS:*
0. Yourself
1. Name 1

2. Name 2
etc.

*COLUMNS:*
1. Fever or high temperature
2. A cough that has lasted for at least several hours
3. Shortness of breath
4. Aches and pains, e.g.  in  back, neck, shoulders or joints
5. Blocked nose
6. Sore throat
7. Feeling unusually tired
8. None of these
9 Don't know
10. Prefer not to answer

*ASK IF Q29=1,2,3,4,5,6 OR 7 (ALL WHO HAVE SUFFERED ANY SYMPTOMS). IF QSAMPLE=1 AND HHCOMPCONFRIM=2 THEN REMOVE ANYONE SELECTED AT HHCOMPREMOVE.  QSAMPLE=1 AND HHCOMPCONFRIM =3 THEN ADD PEOPLE FROM HHCOMPADD*

*MA*

SERVICE
Q30. Have you, or anyone else in your household, done any of the following for these symptoms?
*NOTE FOR SCRIPTER – PLEASE SCRIPT AS GRID – 6 MUST ALWAYS BE LAST*
Please tick all that apply for each row

*ROWS:*
0. Yourself
1. Name 1
2. Name 2
etc.

*COLUMNS:*
1. Phoned NHS 111 or used NHS 111 online service
2. Phoned a GP practice/GP out of hours service
3. Visited a GP practice/GP out of hours service
4. Visited a walk-in centre, urgent care centre, urgent treatment centre or minor injuries unit (PN. DO NOT SHOW IN BE or NL)
5. Visited Accident & Emergency (A&E)
6. Visited a testing location somewhere different to these services
7. Been admitted to hospital
8. Don't know
9. None of these
10. Prefer not to answer

*ASK IF Q30=1,2,3,4,5,6 OR 7 (FOR EACH SERVICE SELECTED AT SERVICE). IF QSAMPLE=1 AND HHCOMPCONFRIM =2 THEN REMOVE ANYONE SELECTED AT HHCOMPREMOVE. QSAMPLE=1 AND HHCOMPCONFRIM =3 THEN ADD PEOPLE FROM HHCOMPADD*

*WRITE IN*
WHEN
Q31. You said that [*IF FOR SELF AT Q30 –* you *OR IF FOR [Q21] AT Q30* – NAME] have/has [*INSERT SERVICE FROM Q30*]. When did [you/they] do that? If you don't know exactly, please provide an approximate date.
Please write in
Please use the format "DD/MM"
INSERT DATE USE DATE FORMAT DD/MM. EARLIEST DATE IS 01/02 AND CANNOT BE A DATE IN THE FUTURE
1. Don't know
2. Prefer not to answer

*ASK ALL*
*SA*
TEST
Q32. Have you, or any other household member, ever been tested for Coronavirus (Covid-19)?

Select only one

1. Yes
2. No

*ASK IF Q32=1*

*ASK FOR SELF AND EACH PERSON NAMED AT Q21*
*SA*
TESTPERSON
Q33. Who has been tested for Coronavirus (Covid-19)?
*ASK FOR SELF AND EACH PERSON NAMED AT Q21/Q21 IMPORT. IF QSAMPLE=1 AND HHCOMPCONFIRM=2 THEN REMOVE ANYONE SELECTED AT HHCOMPREMOVE. QSAMPLE=1 AND HHCOMPCONFIRM=3 THEN ADD PEOPLE FROM HHCOMPADD*

*NOTE FOR SCRIPTER – PLEASE SCRIPT AS GRID*
Select only one for each row

*ROWS:*
0. Yourself
1. Name 1
2. Name 2
etc.

*COLUMNS:*
1. Tested and the test showed I/they have Coronavirus
2. Tested, and the test showed I/they do not have Coronavirus
3. Yes, and I'm still waiting to hear the result
4. Not tested

5. Don’t know
6. Prefer not to answer

*ASK FOR SELF AND EACH PERSON NAMED AT Q21/Q21 IMPORT. IF QSAMPLE=1 AND HHCOMPCONFRIM =2 THEN REMOVE ANYONE SELECTED AT HHCOMPREMOVE. QSAMPLE=1 AND HHCOMPCONFRIM =3 THEN ADD PEOPLE FROM HHCOMPADD*

*SA*
CONTACT
Q34. To the best of your knowledge, do you think you or anyone else in your household have been in direct contact with someone who has Coronavirus (Covid-19) in the last seven days, or know someone close to them who has Coronavirus (Covid-19)?
Select only one for each row

*ASK FOR SELF AND EACH PERSON NAMED AT Q21 Q21/Q21 IMPORT. IF QSAMPLE=1 AND HHCOMPCONFRIM =2 THEN REMOVE ANYONE SELECTED AT HHCOMPREMOVE. QSAMPLE=1 AND HHCOMPCONFRIM =3 THEN ADD PEOPLE FROM HHCOMPADD*

*NOTE FOR SCRIPTER – PLEASE SCRIPT AS GRID*

*ROWS:*
0. Yourself
1. Name 1
2. Name 2
etc.

*COLUMNS:*
1. Yes, currently infected
2. Yes, passed away
3. Yes, recovered
4. No
5. Don’t know
6. Prefer not to answer

**Attitudes**

**Behaviour**

*ASK ALL*
*SA*
*INTRO*
Q39. You may have been asked or decided to participate in different responses to coronavirus (covid-19).
Thinking about the last seven days, please select the appropriate response for each of the interventions listed below.

INTER In the last seven days, [*IF FOR SELF –* have you] [*IF FOR NAME 1/2 ETC AT Q21 –* has NAME] been asked to…
*ASK FOR SELF AND EACH PERSON NAMED AT Q21/Q21 IMPORT. IF QSAMPLE=1 AND HHCOMPCONFRIM =2 THEN REMOVE ANYONE SELECTED AT HHCOMPREMOVE. QSAMPLE=1 AND HHCOMPCONFRIM =3 THEN ADD PEOPLE FROM HHCOMPADD*

*NOTE FOR SCRIPTER – PLEASE SCRIPT AS GRID*

Select only one for each row

*ROWS:*
1. Quarantine [yourself/themselves]
*Quarantine is the act of staying at home after a potential exposure to an infected case. If you are in quarantine, you can leave the house, but limit your movements.*

2. Isolate [yourself/themselves]
*Isolation is the act of separating yourself from people who are not infected, including any household members. You can be in isolation in your house or in a health facility.*
3. Work from home due to coronavirus or limit your/their time at your/their workplace *SHOW IF Q3 = 1,2,3 OR Q25=1,2,3*
4. Limit [your/their] time at the [university or college] [*SHOW IF Q3 = 8 OR Q26 = 3 OR 4*] OR [pre-school or nursery] [*Q26 = 1]* OR [school] [*Q26 = 2*] due to coronavirus (covid-19)

*COLUMNS:*
1. Yes
2. No
3. Not applicable *ONLY SHOW FOR THIRD STATEMENT*
4. Don't know
5. Prefer not to answer

*ASK FOR SELF AND EACH PERSON NAMED AT Q21/Q21 IMPORT. IF QSAMPLE=1 AND HHCOMPCONFRIM =2 THEN REMOVE ANYONE SELECTED AT HHCOMPREMOVE. QSAMPLE=1 AND HHCOMPCONFRIM =3 THEN ADD PEOPLE FROM HHCOMPADD*

*SA*
INTER2
Q40. In the last seven days, has…
Select only one for each row

*NOTE FOR SCRIPTER – PLEASE SCRIPT AS GRID*
*ROWS:*
1. [Your/ NAME's] workplace been closed due to coronavirus (covid-19) for at least one day *ASK IF Q3-CODE 1,2,3 OR Q25 = 1,2,3*
2. [Your/NAME's] university or college] [*ASK IF Q3 = 8 OR Q26 = 3 OR 4]* OR [pre-school or nursery] [*ASK IF Q26 = 1*] OR [school] [*ASK IF Q26 = 2*] been closed for at least one day

*COLUMNS:*
1. Yes
2. No

3. Not applicable *ONLY SHOW FOR FIRST STATEMENT*
4. Don't know
5. Prefer not to answer

*ASK FOR SELF AND EACH PERSON NAMED AT Q21/Q21 IMPORT. IF QSAMPLE=1 AND HHCOMPCONFRIM =2 THEN REMOVE ANYONE SELECTED AT HHCOMPREMOVE. QSAMPLE=1 AND HHCOMPCONFRIM =3 THEN ADD PEOPLE FROM HHCOMPADD*

*SA*
INTER3
Q41. In the last seven days, [*IF FOR SELF –* have you] [*IF FOR NAME 1/2 ETC AT Q21 has NAME*]...
Select only one for each row

*NOTE FOR SCRIPTER – PLEASE SCRIPT AS GRID*

*ROWS:*
1. Been in quarantine for at least one day *Note for scripter to add an (i) button*
*Quarantine is the act of staying at home after a potential exposure to an infected case. If you are in quarantine, you can leave the house, but limit your movements.*
2. Been in isolation for at least one day *Note for scripter to add an (i) button*
*Isolation is the act of separating yourself from people who are not infected, including any household members. You can be in isolation in your house or in a health facility.*
3. Not been to your/their workplace for at least one day due to coronavirus (covid-19) *ASK IF Q3=1,2,3 OR Q25 = 1,2,3*
4. Not attended [university or college] [*ASK IF Q3 = 8 OR Q26 = 3 OR 4*] OR [pre-school or nursery] [*ASK IF Q26 = 1*] OR [school] [*ASK IF Q26 = 2*] due to coronavirus (covid-19) for at least one day

*COLUMNS:*
1. Yes
2. No
3. Not applicable *ONLY SHOW FOR STATEMENT c)*
4. Don't know
5. Prefer not to answer

*ASK FOR EACH PERSON CODED Q39 1 = 1 (YES) OR Q41 1 = 1 (YES)*
*WRITE IN*
QUAR1
Q42. You said that [you have/NAME has] been in quarantine for at least one day. When did [you/they] start the quarantine?

Please write in

Please use the format "DD/MM"

1. INSERT DATE USE DATE FORMAT DD/MM. EARLIEST DATE IS 01/02 AND CANNOT BE A DATE IN THE FUTURE
2. Don't know
3. Prefer not to answer

*ASK FOR EACH PERSON CODED Q39 1 = 1 (YES) OR Q41 1 = 1 (YES). IF QSAMPLE=1 AND HHCOMPCONFRIM =2 THEN REMOVE ANYONE SELECTED AT HHCOMPREMOVE. QSAMPLE=1 AND HHCOMPCONFRIM =3 THEN ADD PEOPLE FROM HHCOMPADD*

*SA*
QUAR2
Q43. And when did [you/NAME] finish the quarantine?

Select one only

Please use the format "DD/MM"

1. INSERT DATE USE DATE FORMAT DD/MM. EARLIEST DATE IS 01/02. DATE MUST NOT BE EARLIER THAN Q42 AND CANNOT BE A DATE IN THE FUTURE
2. I am/they are still in quarantine
3. Don't know
4. Prefer not to answer

*ASK FOR EACH PERSON CODED Q39 2 = 1 (YES) OR Q41 2 = 1 (YES). IF QSAMPLE=1 AND HHCOMPCONFRIM =2 THEN REMOVE ANYONE SELECTED AT HHCOMPREMOVE. QSAMPLE=1 AND HHCOMPCONFRIM =3 THEN ADD PEOPLE FROM HHCOMPADD*
*WRITE IN*

*WRITE IN*
ISO1
Q44. You said that [you have/NAME has] have been in isolation for at least one day. When did [you/they] start isolating?

Please write in

Please use the format "DD/MM"

1. INSERT DATE USE DATE FORMAT DD/MM. EARLIEST DATE IS 01/02 AND CANNOT BE A DATE IN THE FUTURE.
2. Don't know
3. Prefer not to answer

*ASK FOR EACH PERSON CODED Q39 2 = 1 (YES) OR Q41 2 = 1 (YES). IF QSAMPLE=1 AND HHCOMPCONFRIM =2 THEN REMOVE ANYONE SELECTED AT HHCOMPREMOVE. QSAMPLE=1 AND HHCOMPCONFRIM =3 THEN ADD PEOPLE FROM HHCOMPADD*

*SA*
ISO2
Q45. And when did [you/NAME] finish isolating?

Select one only

Please use the format "DD/MM"

1. INSERT DATE USE DATE FORMAT DD/MM. EARLIEST DATE IS 01/02. DATE MUST NOT BE EARLIER THAN Q42 AND CANNOT BE A DATE IN THE FUTURE
2. I am/they are still isolating

3. Don't know
4. Prefer not to answer

*ASK FOR EACH PERSON CODED Q39 3 = 1 (YES) OR Q39 4 = 1 (YES) OR Q40 1 = 1 (YES) OR Q40 2 = 1 (YES).*
*IF QSAMPLE=1 AND HHCOMPCONFRIM =2 THEN REMOVE ANYONE SELECTED AT HHCOMPREMOVE.*
*QSAMPLE=1 AND HHCOMPCONFRIM =3 THEN ADD PEOPLE FROM HHCOMPADD*

*FILTER: (Q39_3 =1 or Q39_4 =1) or (Q40 _1 =1 or Q40 _2 =1)*
*WRITE IN*
CLOSE1
Q46. You said that [your/NAME's] [workplace] [*ASK IF Q3 = 1,2 OR 3 OR Q25=1,2 OR 3*] OR [university or college] [*ASK IF Q3 = 8 OR Q26 = 3 OR 4*] OR [pre-school or nursery] [*ASK IF Q26 = 1*] OR [school] [*ASK IF Q26 = 2*] OR was closed due to coronavirus (covid-19) for at least one day in the last seven days. Please select the date when it was first closed.
*IF CODE (Q39 3 AND Q39 4) OR (Q40 1 AND Q40 2) OR (Q41 3 AND Q41 4) ASK QUESTION ONCE FOR UNIVERSITY OR COLLEGE AND ONCE FOR WORKPLACE*

Please write in

Please use the format "DD/MM"

1. INSERT DATE USE DATE FORMAT DD/MM. EARLIEST DATE IS 01/02. 2 AND CANNOT BE A DATE IN THE FUTURE
2. Don't know
3. Prefer not to answer

*ASK FOR EACH PERSON CODED YES (1) AT Q39 3 OR Q39 4 OR Q40 1 or Q40 2. ASK IMMEDIATELY AFTER Q46 FOR EASH PERSON. IF QSAMPLE=1 AND HHCOMPCONFRIM =2 THEN REMOVE ANYONE SELECTED AT HHCOMPREMOVE. QSAMPLE=1 AND HHCOMPCONFRIM =3 THEN ADD PEOPLE FROM HHCOMPADD*
*FILTER: (Q39_3 =1 or Q39_4 =1) or (Q40 _1 =1 or Q40 _2 =1*
*WRITE IN*
CLOSE 2
Q47. And when did it open again?

Please write in

Please use the format "DD/MM"

1. INSERT DATE USE DATE FORMAT DD/MM. EARLIEST DATE IS 01/02. DATE MUST NOT BE EARLIER THAN Q42 AND CANNOT BE A DATE IN THE FUTURE
2. It is still closed
3. Don't know
4. Prefer not to answer

*ASK FOR EACH PERSON CODED NO (2) AT Q39 3 OR Q39 4 OR Q40 1 OR Q40 2 <u>AND</u> YES (1) AT Q41 3 or Q41 4. IF QSAMPLE=1 AND HHCOMPCONFRIM =2 THEN REMOVE ANYONE SELECTED AT HHCOMPREMOVE. QSAMPLE=1 AND HHCOMPCONFRIM =3 THEN ADD PEOPLE FROM HHCOMPADD*

FILTER: ((Q39_3 =2 or Q40_1=2) AND Q41_3 =1)

OR

((Q39_4 =2 or Q40 _2 =2) AND Q41_4 =1)

*MA*
NOTATTEND1

Q48. You said that [you/NAME] did not attend [the workplace] [*ASK IF Q3 = 1,2 OR 3 OR Q25=1, 2 3 OR 4*] OR [university or college] [*ASK IF Q3 = 8 OR Q26 = 3 OR 4*] OR [pre-school or nursery] [*ASK IF Q26 =1*] OR [school] [*ASK IF Q26 = 2* due to coronavirus (covid-19), but it was not closed. For the last seven days, please select the days when [you/NAME] did not go to [the workplace] [*ASK IF Q3 = 1,2 OR 3 OR Q25 = 1,2 OR 3*] OR [university or college] [*ASK IF Q3 = 8 OR Q26 = 3 OR 4*] OR [pre-school or nursery] [*ASK IF Q26 = 1*] OR [school] [*ASK IF Q26 = 2*] due to coronavirus (covid-19). Only indicate those days where [you/NAME] would normally have gone there.
*IF CODE (Q39 3 AND Q39 4) OR (Q40 1 AND Q40 2) OR (Q41 3 AND Q41 4) ASK QUESTION ONCE FOR UNIVERSITY OR COLLEGE AND ONCE FOR WORKPLACE*
Please tick all that apply

1. SHOW DATES FOR LAST SEVEN DAYS
2. Don't know
3. Prefer not to answer

*ASK FOR EACH PERSON CODED NO (2) AT Q39 3 OR Q39 4 OR Q40 1 OR Q40 2 AND YES (1) AT Q41 3 or Q41 4*
FILTER: ((Q39_3 =2 or Q40_1=2) AND Q41_3 =1)

OR

((Q39_4 =2 or Q40 _2 =2) AND Q41_4 =1)

*MA*
Q48A.
What was your main reason for not attending [the workplace] [*ASK IF Q3 = 1,2 OR 3 OR Q25=1, 2 3 OR 4*] OR [university or college] [*ASK IF Q3 = 8 OR Q26 = 3 OR 4*] OR [pre-school or nursery] [*ASK IF Q26 =1*] OR [school] [*ASK IF Q26 = 2*]?Please tick all that apply

1. 7 day isolation due to you having symptoms that may be coronavirus

4. 14 day quarantine due to someone else in your household having symptoms that may be coronavirus, or due to contact with a known coronavirus case
5. Other illness (not coronavirus) within the household, (including yourself
6. Caring for someone outside of the household who has been confirmed to have coronavirus (COVID-19)
7. Caring for someone outside of the household who has not been confirmed to have coronavirus (COVID-19)
8. At least one child in my household is home due to school closure

9. Other

*ASK FOR EACH PERSON CODED YES (1) OR NOT APPLICABLE (3) AT Q39 3 OR Q40 1 OR Q41 3. IF QSAMPLE=1 AND HHCOMPCONFRIM =2 THEN REMOVE ANYONE SELECTED AT HHCOMPREMOVE. QSAMPLE=1 AND HHCOMPCONFRIM =3 THEN ADD PEOPLE FROM HHCOMPADD*

FILTER: (Q39 _3=1,3) or (Q40_ 1=1,3) or (Q41_ 3=1,3) *MA*
INCOME
Q49. You said that [you/NAME] did not work/visit your workplace for at least one day due to coronavirus (covid-19) or that this was not applicable. Did this have a negative impact on your household income?
Please tick all that apply

1. No, [I/NAME] was able to work from home
2. No, [I/NAME] was able to take carer leave
3. No, but [I/NAME] had to take annual leave
4. No, as [I/NAME] was fully compensated by [my/their] employer
5. No, as [I/NAME] was fully compensated by the government
6. Yes, but [I/NAME] received partial compensation by [my/their] employer
7. Yes, but [I/NAME] received partial compensation by the government
8. Yes, and [I/NAME] received no compensation for [my/their] lost income [SINGLE CODE ONLY]
9. Other (please specify)
10. Don't know
11. Prefer not to answer

*ASK FOR EACH PERSON CODED (1 OR 2 OR 8 AT Q26) AND (Q39 4 OR Q40 2. IF QSAMPLE=1 AND HHCOMPCONFRIM =2 THEN REMOVE ANYONE SELECTED AT HHCOMPREMOVE. QSAMPLE=1 AND HHCOMPCONFRIM =3 THEN ADD PEOPLE FROM HHCOMPADD*
*)*
*MA*
FILTER: any(Q26,1,2,8) and (Q39_4=1 or Q40_2=1)

CHILDCARE
Q50. You said that [NAME's] [pre-school or nursery] [*ASK IF Q26 = 1*] OR [school] [*ASK IF Q26 = 2*] was closed for at least one day due to coronavirus (covid-19). When this happened, who looked after the child/children?
CHILDCARE2
Please tick all that apply

1. A parent, who is unemployed
2. A parent, who was working from home
3. A parent, who works part-time
4. A parent, who took annual leave
5. A parent, who took carer leave
6. A parent, who took unpaid leave
7. A sibling
8. Grandparent(s)
9. A baby sitter, childminder, au pair or nanny (paid)

10. A baby sitter, childminder, au pair or nanny (unpaid)
11. A neighbour, friend, uncle, or aunt
12. Not required **[PN: Exclusive]**?
13. Other (please specify)
14. People at the school, as my child was eligible for childcare at the school

*ASK FOR EACH PERSON CODED 1 or 2 or 8 AT Q26 AND 1 AT q41 4 AND 2 AT Q39_4 AND 2 AT Q40_2. IF QSAMPLE=1 AND HHCOMPCONFRIM =2 THEN REMOVE ANYONE SELECTED AT HHCOMPREMOVE. QSAMPLE=1 AND HHCOMPCONFRIM =3 THEN ADD PEOPLE FROM HHCOMPADD*

FILTER: Q26=1,2, 8 and Q41_4 =1 and Q39_4 =2 and Q40_2 =2*MA*

CHILDCARE2
Q51. You said that [NAME] did not attend [pre-school or nursery] [*ASK IF Q26 = 1*] OR [school] [*ASK IF Q26 = 2*] for at least one day due to coronavirus (covid-19). When this happened, who looked after the child/children?
Please tick all that apply

1. A parent, who is unemployed
2. A parent, who was working from home
3. A parent, who works part-time
4. A parent, who took annual leave
5. A parent, who took carer leave
6. A parent, who took unpaid leave
7. A sibling
8. Grandparent(s)
9. A baby sitter, childminder, au pair or nanny (paid)
10. A baby sitter, childminder, au pair or nanny (unpaid)
11. A neighbour, friend, uncle, or aunt
12. Not required **[PN: Exclusive]**?
13. Other (please specify)
14. People at the school, as my child was eligible for childcare at the school

*IF QSAMPLE=2*
*ASK ALL WHO HAVE MORE THAN ONE CHILD AGED UNDER 18 AT Q23 (MORE THAN 1 CODE 1-4 AT Q23)*
*Q53a*
*SA*
*SELECT CHILD*
We need to choose one child at random for you to think about when answering the remaining questions. To do this, please tell us which of your children will have their birthday next. If you have two or more children with birthdays on the same day, please select the one whose name comes first alphabetically. Please only think about the children you are a parent to.

*LIST ALL CHILDREN FROM Q21 IF THEY ARE UNDER THE AGE OF 18 (CODES 1-4) AT Q23*

*SHOW TEXT FOR ALL*
When answering the following questions, please answer only on behalf of [*IF MORE THAN ONE CHILD AGED UNDER 18 AT Q23 INSERT CHILD'S NAME FROM Q53a OR IF ONE CHILD AGED UNDER 18 AT Q23 INSERT CHILD'S NAME FROM Q21]*. Please answer to the best of your ability, with your best guess if you do not know the exact answer.

*IF QSAMPLE=1*

*SHOW TEXT FOR ALL*

*IF QSAMPLE=1 THEN ASK ALL SUBSEQUENT QUESTIONS [Q54, Q59, Q60, Q61a, Q61b, Q62, Q63, Q66, Q67, Q68, Q69, Q70, Q71, Q72, Q73, Q74, Q75, Q76] ABOUT CHILD SELECTED AT INITIAL WAVE in Q53a OR IF ONE CHILD AGED UNDER 18 AT Q23 INSERT CHILD'S NAME FROM Q21*

*If person selected at initial wave is removed at hhcompremove, then make a new allocation*

Last time you completed this survey for us you told us about INSERT CHILD'S NAME from Q53a When answering the following questions, please answer only on behalf of INSERT CHILD'S NAME from Q53a]. Please answer to the best of your ability, with your best guess if you do not know the exact answer.

*SHOW TEXT FOR ALL*

These questions are voluntary but they are really important in helping us understand the spread of COVID-19 and the impact of different public health interventions. It will not be possible to identify you or any member of your household in the published findings.

**Individual preventive measures**

*ASK ALL*
*SA*
*MASK1*
Q54. Did [INSERT CHILD'S NAME from Q53a] use a face mask yesterday?
Select only one
1. Yes
2. No

*ASK ALL*
*MA*
TRANSPORT
Q59. Did [INSERT CHILD'S NAME from Q53a] travel on any public transport yesterday?

Please tick all that apply
1. No *[SINGLE CODE]*
2. Train/tube
3. Bus/tram
4. Taxi, Uber, or similar ride-hailing app
5. Aeroplane

*ASK ALL WHO TRAVELLED ON PUBLIC TRANSPORT (Q59 = CODES 2-5, FOR EACH TYPE)*
*WRITE IN*
TRANSDUR
Q60. And approximately how long did [INSERT CHILD'S NAME from Q53a] spend on the [*INSERT FROM Q59*] in total?

Please write in

INSERT NUMBER OF HOURS
INSERT NUMBER OF MINUTES
**ADDITIONAL CHILDRENS QUESTIONS**

*ASK IF SELECTED CHILD'S Q47 DOES NOT = 2 AND IF Q26 = 1,2,3 OR 4*
Please answer the following questions about [INSERT CHILD'S NAME from Q53a]'s attendance at [pre-school or nursery] [*IF CHILD Q26 = 1*] OR [school] [*IF CHILD Q26 = 2*] OR [secondary school/college] [*IF Q26 = 3*] or [their university or other higher educational institution] [*IF Q26 = 4*]

**Q61a.** Did [INSERT CHILD'S NAME from Q53a] attend [pre-school or nursery] [*IF CHILD Q26 = 1*] OR [school] [*IF CHILD Q26 = 2*] OR [secondary school/college] [*IF Q26 = 3*] or [their university or other higher educational institution] [*IF CHILD Q26 = 4*] yesterday?

1. Yes
2. No, but it was open for my child
3. Not applicable as it was a weekend/holiday/day off
4. Not applicable as it was closed
5. Don't know
6. Prefer not to answer

*ASK IF Q61a = 1*
Q61b. How many [children/people] [IF CHILD Q26 = 1,2, or 3] OR [people] [IF CHILD Q26 = 4] were in [INSERT CHILD'S NAME from Q53a]'s [pre-school or nursery] [ *IF CHILD Q26 = 1*] OR [class] [ *IF CHILD Q26 = 2*] OR [secondary school/college ] [*IF CHILD Q26 = 3*] OR [class/lecture] [*IF CHILD Q26=4*] yesterday?

If you are unsure of the exact answer, please enter your best guess. Please answer this question only for yesterday.

INSERT TEXT [*restrict to numbers only up to max 500]*
Don't know
Prefer not to answer

**Contact survey**
*ASK ALL*
Q62a. Please can you write in a nickname for yourself? Note that this nickname is only needed to make it easier for you to complete the survey, so please pick a nickname that is easy to remember. Your individual responses will not be shared with anyone outside this survey.
*WRITE IN*

*INTRO SCREEN*
We will now ask you to remember who [INSERT CHILD'S NAME from Q53a] had been in contact with yesterday, between 5am yesterday and 5am today. We are only interested in direct contacts, which are **people who** [INSERT CHILD'S NAME from Q53a]**met in person** and with whom they exchanged at least a few words, or with whom they had physical contact (e.g. a handshake, embracing, kissing, contact sports).
**Note that if [INSERT CHILD'S NAME from Q53a] only spoke to someone over the phone or internet, you should answer 'No' in this section.**

*ASK ALL*
*SA*
CONTACT1
Q62. Which of the following people did ~~you~~ [INSERT CHILD'S NAME FROM Q53A] have direct contact with in person, between 5am yesterday and 5am today, in person?
*NOTE FOR SCRIPTER – PLEASE SCRIPT AS GRID*

*~~Note for scripter to add (i) button for 'direct contact':~~*
*We are only interested in direct contacts, which are people who you [INSERT CHILD'S NAME FROM Q53A] met in person and with whom ~~you~~ they exchanged at least a few words, or with whom ~~you~~/they had physical contact (e.g. a handshake, embracing, kissing, contact sports).*
***Note that if ~~you~~ [INSERT CHILD'S NAME FROM Q53A] only spoke to someone over the phone or internet, ~~they~~ you should answer 'No' ~~not be included~~ in this section.***

*ROWS:*
*INSERT ALL NAMES FROM Q21/Q21IMPORT EXCEPT CHILD"S NAME, AND* ***INCLUDE THE PARTICIPANT'S NAME FROM Q62a.***

*COLUMNS:*
1. Yes
2. No

*ASK ALL*
*WRITE IN*
CONTACT2
Q63. And what **other people, outside of your household**, did [INSERT CHILD'S NAME FROM Q53A] have direct contact with in person, between 5am yesterday and 5am today? This could include friends, family, classmates, or people they spoke to in shops and so on.

Please write the nickname of each person [INSERT CHILD'S NAME FROM Q53A] had direct contact with ~~below~~. Note that this nickname is only needed to make it easier for you to complete the survey, so please pick a nickname that is easy to remember. Your individual responses will not be shared with anyone outside this survey.

*We are only interested in people who* [INSERT CHILD'S NAME FROM Q53A] *met in person and* ***with whom they exchanged at least a few words, or with whom ~~you~~ they had physical contact*** *(e.g. a handshake, embracing, contact sports)***.**

It is easiest to list names in chronological order, e.g. When [INSERT CHILD'S NAME FROM Q53A] woke up they saw our neighbours Pete and Naomi. We then drove to school where they had lunch with their friends and Deborah. On our way back home, we stopped to see their older cousin, Kate, at a café and spoke to the staff (assign a nickname to staff). Etc.

**Please do not list yourself or anyone that you listed as being a part of your household. Note that if [INSERT CHILD'S NAME FROM Q53A] only spoke to *someone* over the phone or internet, *they should not be included in this section*.**

*PN: SHOW IF Q20 IS NOT NONE OR IF NO-ONE IS SELECTED AT Q62]*

*You have already indicated contact with the following household members, add additional contacts in the textboxes below:*

LIST HOUSEHOLD MEMBERS SELECTED AT Q62 AND THE PARTICIPANT FROM Q62a

*ALLOW BOXES TO INPUT NICKNAMES*
*PN SHOW 100 BOXES ON SCREEN*

*PN AT TOP OF PAGE WITH TEXT BOXES INCLUDE THE FOLLOWING.*

*PLEASE LIST ALL OTHER CONTACTS YOU HAVE OUTSIDE OF YOUR HOUSEHOLD, AS BEFORE PLEASE ASSIGN A **NICKNAME** TO EACH OF THESE CONTACTS*

*No-one/No other people [PN. ALLOW THIS TO BE SELECTED WHEN TEXT BOXES COMPLETED]*

*ASK THE FOLLOWING LOOP Q69 TO Q72 TO TIME FOR EACH NAME GIVEN AT Q62*
*ASK THE FOLLOWING LOOP Q66 TO Q72 TO TIME FOR EACH NAME GIVEN AT Q63*

*ASK FOR EACH PERSON AT Q63*
*SA*

Q66. Which of the following age groups does NAME fit into? Please give an estimate if you are not sure
Select only one

1. Under 1
2. 1-4
3. 5-9
4. 10-14
5. 15-19
6. 20-24
7. 25-34
8. 35-44
9. 45-54
10. 55-64
11. 65-69
12. 70-74
13. 75-79
14. 80-84
15. 85+
16. Don't know
17. Prefer not to answer

18. This person is [INSERT CHILD'S NAME FROM Q53A]
19. This is me (INSERT PARTICIPANT NICKNAME FROM Q62A)

*ASK ALL WHERE NAME GIVEN AT Q63 AND Q66 IS NOT EQUAL TO CODE 18 or CODE 19*
*SA*
CONTACTGEN
Q67. As far as you know, which of the following describes how [NAME] thinks of themselves?
Select only one

1. _1 Male
2. _2 Female
3. _3 In another way
4. _4 Prefer not to answer
5. _5 Don't know

*ASK ALL WHERE NAME GIVEN AT Q63 AND Q66 IS NOT EQUAL TO CODE 18 or CODE 19*
*SA*
CONTACTRELAT
Q68. What is [NAME]'s relationship to [INSERT CHILD'S NAME FROM Q53A]?
Select only one
1. They are family members not in our household
2. They are someone I work with
3. They are someone they go to nursery, pre-school, school, college or university with
7. They are a babysitter, childminder, or nanny
4. They are friends
5. Other
6. Prefer not to answer

*ASK ALL WHERE NAME GIVEN AT Q62 OR Q63 AND Q66 IS NOT EQUAL TO CODE 18 or CODE 19*
*SA*
CONTACTFREQ
Q69. Before the coronavirus epidemic started, how often did [INSERT CHILD'S NAME FROM Q53A] usually have direct contact with [*NAME*]? A direct contact is when [INSERT CHILD'S NAME FROM Q53A] **meets with this person in person** and when they exchange at least a few words, or when they have physical contact (e.g. a handshake, high five, embracing, touching while playing, contact sports).
**Please do not include times that [INSERT CHILD'S NAME FROM Q53A] speaks to them over the phone or internet.**
Select only one for each person

1. Every day or almost every day
2. About once or twice a week
3. Every 2-3 weeks
4. About once per month
5. Less often than once per month
6. Never met them before
7. Prefer not to answer

*ASK ALL WHERE NAME GIVEN AT Q62 OR Q63 AND Q66 IS NOT EQUAL TO CODE 18 or CODE 19*
*SA*

PHYS
Q70. When [INSERT CHILD'S NAME FROM Q53A] had direct contact with [NAME] yesterday, did [INSERT CHILD'S NAME FROM Q53A] have…?
Select only one

1. Physical contact (any sort of skin-to-skin contact such as e.g. high fiving, embracing, touching while playing or contact sports)
2. Non-physical contact (they did not touch the person)
3. Prefer not to answer

*ASK ALL WHERE NAME GIVEN AT Q62 OR Q63 AND Q66 IS NOT EQUAL TO CODE 18 or CODE 19*
*MA*
WHERE
Q71. And where did [INSERT CHILD'S NAME FROM Q53A] have direct contact with [NAME]?
Please tick all that apply

1. At home (including at the door, in the garden, and within entrances to the home such as stairways, lifts, and corridors)
2. At someone else's house
3. At work
4. At a place of worship
5. On any form of transport
6. At university, school, pre-school, or nursery
7. At a shop for essentials, eg a supermarket, grocery store, market, pharmacist, or bicycle shop
8. At a shop for non--essential items, eg a gardening center, or a clothes, electronics, furniture, or DIY shop
9. At a place of entertainment such as a restaurant or cinema
10. At a place for sports such as a gym or sports club/match [PN. DO NOT SHOW IN BE AND NL]
11. Outside, for example in a park, on the street or in the countryside
14. In a healthcare setting, eg hospital, GP, A&E, outpatient facility, dentist, physiotherapist, optometrist, etc

15. At a hair dresser, barber, nail salon, beauty parlor or similar location

16. Somewhere else (please specify)

*ASK ALL WHERE NAME GIVEN AT Q62 OR Q63 AND Q66 IS NOT EQUAL TO CODE 18 or CODE 19*
*WRITE IN*
TIME
Q72. Please estimate the total amount of time [INSERT CHILD'S NAME FROM Q53A] spent with [NAME] in person yesterday.

Please write in

*INSERT HOURS*
*INSERT MINUTES*

ASK If COUNTRY = UK AND Q71= 1, 2, 3, 4, 6, 7, 9, 10 or 12, 14 or 15

Q73 Was the time [INSERT CHILD'S NAME FROM Q53A] spent with [NAME] yesterday inside or outside?
Please tick all that apply
Inside
Outside

*SA*
*ASK ALL WHO COMPLETE AT LEAST ONE CONTACT AT Q63*
Q74. We do ask you to individually include every contact [INSERT CHILD'S NAME FROM Q53A] had, but if you were unable to include every single contact (for instance, because they were at school and have a large number of contacts in a day), please could you indicate this?

Select only one

1. I individually included every person [INSERT CHILD'S NAME FROM Q53A] had contact with.
2. I did not individually include every person [INSERT CHILD'S NAME FROM Q53A] had contact with.
3. [INSERT CHILD'S NAME FROM Q53A] did not have any contacts (PN: ONLY SHOW IF NO CONTACTS LISTED AT Q63 I.E. ALL TEXT BOXES WERE LEFT EMPTY AND NO CONTACTS ARE CODED AS 1 AT Q62)

PN: SHOW IF Q74= 2

Q75. Approximately how many people did [INSERT CHILD'S NAME FROM Q53A] have contact with and **you did not list** individually? Please provide your best estimate by age and setting.

*For this question, we are only interested in people who* [INSERT CHILD'S NAME FROM Q53A] *met in person and **with whom*** [INSERT CHILD'S NAME FROM Q53A] ***exchanged at least a few words, OR with whom*** [INSERT CHILD'S NAME FROM Q53A] ***had physical contact** (e.g.* **high fiving, embracing, touching while playing or contact sports*)*.**

*Please enter number of contacts for each age group and setting*
*PN INSERT TABLE WITH ROWS/COLUMNS BELOW FOR RESPONDENTS TO ENTER NUMBER OF CONTACTS*

"PN MINIMUM VALUE SHOUD BE 0 AND MAX 2000,. PARTICIPANTS CANNOT CODE 0 FOR EVERY BOX."
ROWS
1. Under 18
2. 18 – 64
3. 65 and over

COLUMNS
1. At work
2. At school or other educational setting
3. Somewhere else

PN: SHOW IF Q74= 2
"PN MINIMUM VALUE SHOUD BE 0 AND MAX 2000"

Q76. And approximately how many people did [INSERT CHILD'S NAME FROM Q53A] have **physical** contact with and you did not list individually? Please provide your best estimate by age and setting.

*For this question, we are only interested in people who* [INSERT CHILD'S NAME FROM Q53A] *met in person* ***AND had physical contact (e.g.*** **high fiving, embracing, touching while playing or contact sports*).***

*Please enter number of contacts for each age group and setting*
*PN INSERT TABLE WITH ROWS/COLUMNS BELOW FOR RESPONDENTS TO ENTER NUMBER OF CONTACTS*

ROWS
1. Under 18
2. 18 – 64
3. 65 and over

COLUMNS
1. At work
2. At school or other educational setting
3. Somewhere else

### CoMix Contact Survey – Panels E & F Final Version

**QCOUNTRY**

1. UK

**CAPTURE DATA FROM IMPORT:**

**Qsample=1 WAVE-TO-WAVE**
**Qsample=2 FRESH SAMPLE**

**CAPTURE DATA FROM IMPORT:**

**Sampletype=1 Main sample**
**Sampletype=2 Parent sample**

**Panel**

1. **Panel E**
2. **Panel F**

**Wave**

1. **Wave 1**
2. **Wave 2**
3. **Wave 3**
4. **Wave 4**
5. **Wave 5**
6. **Wave 6**
7. **Wave 7**
8. **Wave 8**
9. **Wave 9**
10. **Wave 10**
11. **Wave 11**
12. **Wave 12**
13. **Wave 13**
14. **Wave 14**
15. **Wave 15**
16. **Wave 16**
17. **Wave 17**
18. **Wave 18**
19. **Wave 19**
20. **Wave 20**

**Demographics**

*[Standard Screener: DO NOT MODIFY OR TRANSLATE]*
RESP_AGE [Hidden].
*ASK ALL*
*SA*
What is your age in years?
*YEAR/MONTH*
Hidden Question - RESP_AGE "this is a dummy question that will hold age"
USE RESP_AGE [Hidden] response list
1. _18 "18", [KEEP]
2. _19 "19", [KEEP]
3. _20 "20", [KEEP]
4. _21 "21", [KEEP]
5. _22 "22", [KEEP]
6. _23 "23", [KEEP]
7. _24 "24", [KEEP]
8. _25 "25", [KEEP]
9. . . . [KEEP]
10. _65 "65", [KEEP]
11. _66 "66", [KEEP]
12. _67 "67", [KEEP]
13. _68 "68", [KEEP]
14. _69 "69", [KEEP]
15. _70 "70", [KEEP]
16. _71 "71", [KEEP]
17. _72 "72", [KEEP]
18. _73 "73", [KEEP]
19. _74 "74", [KEEP]
20. _75 "75", [KEEP]
21. _76 "76", [KEEP]
22. _77 "77", [KEEP]
23. _78 "78", [KEEP]
24. _79 "79", [KEEP]
25. _80 "80", [KEEP]
26. _81 "81", [KEEP]
27. _82 "82", [KEEP]
28. _83 "83", [KEEP]
29. _84 "84", [KEEP]
30. _85 "85", [KEEP]
31. _86 "86", [KEEP]
32. _87 "87", [KEEP]
33. _88 "88", [KEEP]
34. _89 "89", [KEEP]
35. _90 "90", [KEEP]

36. _91 "91", [KEEP]
37. _92 "92", [KEEP]
38. _93 "93", [KEEP]
39. _94 "94", [KEEP]
40. _95 "95", [KEEP]
41. _96 "96", [KEEP]
42. _97 "97", [KEEP]
43. _98 "98", [KEEP]
44. _99 "99", [KEEP]
45. _100 "100", [KEEP]
46. _101 "101", [KEEP]
47. _102 "102", [KEEP]
48. _103 "103", [KEEP]
49. _104 "104", [KEEP]
50. _105 "105", [KEEP]
51. _998 "Age not calculated", [KEEP]
52. _999 "Age < 1" [KEEP]

*TERMINATE IS RESP_AGE < 18 YO*

*RECODE QUOTAGERANGE*

| |
|---|
| 18-24 |
| 25-34 |
| 35-44 |
| 45-54 |
| 55-64 |
| 65+ |

ASK ALL

*SA*
*[Standard Screener: DO NOT MODIFY OR TRANSLATE]*
[PN: STOP_REALLOCATION if GENDER_NONBINARY is 3 or 4, and RESP_GENDER was not answered in a previous survey]
GENDER_NONBINARY. Which of the following describes how you think of yourself?
Select only one

1. _1 Male [KEEP]
2. _2 Female [KEEP]
3. _3 In another way [KEEP]
4. _4 Prefer not to answer [KEEP]

*[Standard Screener: DO NOT MODIFY OR TRANSLATE]*

[PN: If Prefer not to answer is selected, ask UKREGION1]

QMktSize_GB. Where do you live? Please note: This question may be considered personal. We would like to remind you that your participation is strictly voluntary and that your responses are used for research purposes only. The answers that you provide will be presented in aggregate form and none of them will be linked back to you in any way. All data will be collected and processed in adherence to the Market Research Society's Code of Conduct and the General Data Protection Regulation (GDPR).

| |
|---|
| **Postcode**<br>**Postal Town**<br>**Prefer Not to Answer** |

**Ask only if Qsample=1 and sampletype=2**
QPConfirm1a. To date, you have completed *InsertAllCompletedWaves* surveys. Have you answered the contact section, where we ask the number of direct contacts your child has had, on behalf of the same child in every wave or for different children in some waves?
SELECT ONE

1. I have responded on behalf of the same child in all survey waves.
2. I have completed the survey on behalf of different people in my household, depending on the survey wave.

IF QPConfirm1a = 1
**QPConfirm1b** At the contact section, where you were asked to recall your child's contact with people outside the household, can we confirm which of the following household members you completed it on behalf of?

1. W1 – QP53a NAMESEXAGE
2. W2 – QP53a NAMESEXAGE
3. W3 – QP53a NAMESEXAGE
4. W4 – QP53a NAMESEXAGE
5. W5 – QP53a NAMESEXAGE
6. W6 – QP53a NAMESEXAGE
7. W7 – QP53a NAMESEXAGE
8. ….
9. Don't know
10. Prefer not to say

*IF Qsample=1 ASK*

*HHCOMPCONFRIM In the last wave of research these are the people that you said live in your household. Please could you confirm that this is correct?*
*LIST ALL HOUSEHOLD MEMBERS LISTED AT Q21 FROM PREVIOUS WAVE*
*IN CASE THERE ARE NO RESPONDENTS CAPTURED FROM IMPORT CODE 2 WILL NOT BE SHOWN.*

1. *Yes, this is correct (EXCLUSIVE)*
2. *No, one or more of these people no longer live in my household*
3. *No, there are one or more new people now living in my household*

*IF HHCOMPCONFIRM=2 ASK*

*HHCOMPREMOVE Please indicate which of these people **no longer** live in your household?*
*LIST ALL HOUSEHOLD MEMBERS AT Q21 FROM PREVIOUS WAVE AND ALLOW SELECTION OF THOSE WHO NO LONGER LIVE IN THE HOUSEHOLD.*

*IF HHCOMPCONFIRM=3 ASK*
*HHCOMPADD Please write in below the nicknames of the new people who now live in your household?*
*INSERT TEXTBOXES ONE AT A TIME TO RECORD NICKAMES OF NEW PEOPLE – ADD UP TO 4 TEXT BOXES*

*IF Qsample=1 HIDE AND IMPORT DATA, CREATE RECODE VARIABLE WITH THE NAME Q20.*
*IF Qsample=2 ASK Q20*

*ASK ALL*
*SA*
QHOUSE
Q20. Including you, how many people live in your household? By household, we mean anyone living at the same address as you, that you share a kitchen with.
Select only one

1
2
3
4
5
6
7
8
9
10
11 or more

*IF Q20=1 AND Sampletype=2, THANK AND CLOSE*

*ASK IF Q20=2 OR MORE (I.E. IF NOT A SINGLE PERSON HOUSEHOLD) OR HHCOMPCONFIRM <>1 (IF QSAMPLE=1 AND HHCONFIRM <>1). IF QSAMPLE=1 AND HHCONFIRM=1 HIDE AND IMPORT DATA*

Q**HH**L**V**
**{ASK ALL}**
**MA**
Q20b. Who do you live with at home?
*Select all that apply*

1. Spouse/Partner
2. Son/Daughter: own, step, foster children: under the age of 18
3. Son/Daughter: own, step, foster children: 18 or older
4. Grandchild: under the age of 18
5. Grandchild: 18 or older
6. Parent(s), step parent(s), parent(s) in law
7. Brother(s) /Sister(s): under the age of 18
8. Brother/Sister: 18 or older
9. Other relative

10, Other non-relative e.g. friends, housemates
11. On your own (EXCLUSIVE)

*IF sampletype=2 and Q20b=2 CONTINUE*
*IF sampletype=2 Q20ba<>2THANK AND CLOSE*

**MAIN INTRO SCREEN**
Thank you for agreeing to take part in this important research about the new coronavirus (COVID-19) pandemic. Please take the time to carefully read the information on the next screen and the detailed survey participation notice.
The survey is being run by Ipsos on behalf of a group of experts in mathematical and statistical modelling of infectious diseases and public health at the London School of Hygiene and Tropical Medicine (LSHTM); United Kingdom.

Throughout the survey you may see the (i) symbol next to a question or an answer option. You can touch or click on it for extra information to help you answer the question.

*NEW SCREEN*

The research is part of a larger programme of work funded by the UK Medical Research Council (MRC) and the EpiPose Consortium which aims to provide urgently needed answers about the epidemiological characteristics of COVID-19, the social dynamics of the outbreak, and the related public health preparedness and response to the ongoing pandemic, as well as to assess its economic impact. It will be used directly on active communication and interaction with policy makers/authorities, other scientific groups and the general public, to help minimise COVID-19's public health, economic and social impact. The study seeks to understand the spread of COVID-19. It covers things such as who currently makes up your household, recent behaviour and any preventative measures used, as well as people [you have– IF SAMPLETYPE=1]/ [your child has – IF SAMPLETYPE=2] been in contact with.

Taking part in this survey is completely voluntary, and you may refuse to do so. However, we would really value your support as the findings will provide invaluable information for decision making and strategies developed in the near future.

*NEW SCREEN*

**Ethics form for participants**

I confirm that I am 18 years or older.

I confirm that I have read and understood the information for this study. I have had the opportunity to consider the information before taking part in this study.

I understand that taking part is voluntary, and that I am free to withdraw at any time without giving any reason and without any of my rights being affected.

I understand that relevant sections of pseudonymized data collected during the study may be looked at by authorised individuals from the London School of Hygiene & Tropical Medicine, UK Medical Research Council (MRC) and the EpiPose Consortium.

I understand that ANONYMIZED data about me may be shared via a public data repository and that I and any of my household members or contacts will not be identifiable from this information.

I understand that I will be invited to complete multiple questionnaires over the course of the ongoing epidemic.

1. Agree to take part
2. Do not agree to take part [screen out]

*SA*

Q3. What is your current employment status?

Select only one

1. _1 Employed full-time (34 hours or more) [KEEP]
2. _2 Employed part-time (less than 34 hours) [KEEP]
3. _3 Self employed [KEEP]
4. _4 Unemployed but looking for a job [KEEP]
5. _5 Unemployed and not looking for a job
6. _6 Full-time parent, homemaker [KEEP]
7. _7 Retired [KEEP]
8. _8 Student/Pupil [KEEP]
9. _9 Long-term sick or disabled [KEEP]

*ASK IF QSAMPLETYPE=1*

*SA*

Q3b. Do you attend either of the following as a pupil or student?

Select only one

1. Further education, e.g. college
2. Higher education, e.g. university
3. None of the above
4. Don’t know
5. Prefer not to answer

*ASK IF QSAMPLETYPE=1 and Q3=8*
*SA*
Q3c. Are you also employed?

1. _1 Employed full-time (34 hours or more) [KEEP]
2. _2 Employed part-time (less than 34 hours) [KEEP]
3. No

*ASK IF QSAMPLETYPE=1 and (Q3=1 or 2 or Q3c=1 or 2)*
*SA*
Q3d. Are you currently furloughed?

1. Yes
2. No

*ASK ALL*

*[Standard Screener: DO NOT MODIFY OR TRANSLATE]*

[To get UK01SG, need to ask EU01HINC. If EU01HINC=1,2, then ask UK01OCCR. If EU01HINC=3, ask UK01OCCHI. UK01SG is computed in the same manner as in the IIS panel. Details about that on panelstats.]

*[Standard Screener: DO NOT MODIFY OR TRANSLATE]*

[PN: if HHCMP10=1, then do not ask EU01HINC Q4 and assume EU01HINC Q4=_1]

*IF Qsample=1 HIDE AND IMPORT DATA, CREATE RECODE VARIABLE WITH THE NAME EU01HINC*
*IF Qsample=2 ASK EU01HINC*
*SA*
*EU01HINC.*

Are you the one in your household who has the highest income? [person with the largest income from employment, pensions, state benefits, investments or other sources]

Select only one

1. _1 Yes [KEEP]
2. _2 Yes, together with another household member [KEEP]
3. _3 No [KEEP]

*ASK IF IN UK AND EU01HINC=1 OR 2*
*[Standard Screener: DO NOT MODIFY OR TRANSLATE]*

*IF Qsample=1 HIDE AND IMPORT DATA, CREATE RECODE VARIABLE WITH THE NAME UK01OCCR.*
*IF Qsample=2 ASK UK01OCCR.*

*SA*
*UK01OCCR.*

In which of the below categories does your occupation fall? If retired or unemployed, please indicate the category closest to your previous occupation.

Select only one

1. USE UK01OCCR response list
2. Legislators, senior officials and managers [EXPANDABLE HEADER] [KEEP]
   a. _1100 Legislators and senior officials [KEEP]
3. Corporate managers [EXPANDABLE HEADER] [KEEP]
   a. _1210 Directors and chief executives [KEEP]
4. Production and operations department managers [EXPANDABLE HEADER] [KEEP]
   a. _1221 Production and operations department managers in agriculture, hunting, forestry and fishing [KEEP]
   b. _1222 Production and operations department managers in manufacturing [KEEP]
   c. _1223 Production and operations department managers in construction [KEEP]

   d. _1224 Production and operations department managers in wholesale and retail trade [KEEP]
   e. _1225 Production and operations department managers in restaurants and hotels [KEEP]
   f. _1226 Production and operations department managers in transport, storage and communications [KEEP]
   g. _1227 Production and operations department managers in business services [KEEP]
   h. _1228 Production and operations department managers in personal care, cleaning and related services [KEEP]
   i. _1229 Production and operations department managers not elsewhere classified [KEEP]
5. Other department managers [EXPANDABLE HEADER] [KEEP]
   a. _1231 Finance and administration department managers [KEEP]
   b. _1232 Personnel and industrial relations department managers [KEEP]
   c. _1233 Sales and marketing department managers [KEEP]
   d. _1234 Advertising and public relations department managers [KEEP]
   e. _1235 Supply and distribution department managers [KEEP]
   f. _1236 Computing services department managers [KEEP]
   g. _1237 Research and development department managers [KEEP]
   h. _1239 Other department managers not elsewhere classified [KEEP]
6. General managers [EXPANDABLE HEADER] [KEEP]
   a. _1311 General managers in agriculture, hunting, forestry/ and fishing [KEEP]
   b. _1312 General managers in manufacturing [KEEP]
   c. _1313 General managers in construction [KEEP]
   d. _1314 General managers in wholesale and retail trade [KEEP]
   e. _1315 General managers of restaurants and hotels [KEEP]
   f. _1316 General managers in transport, storage and communications [KEEP]
   g. _1317 General managers of business services [KEEP]
   h. _1318 General managers in personal care, cleaning and related services [KEEP]
   i. _1319 General managers not elsewhere classified [KEEP]
7. Physical, mathematical and engineering science professionals [EXPANDABLE HEADER] [KEEP]
   a. _2110 Physicists, chemists and related professionals [KEEP]
   b. _2120 Mathematicians, statisticians and related professionals [KEEP]
   c. _2130 Computing professionals [KEEP]
8. Architects, engineers and related professionals [EXPANDABLE HEADER] [KEEP]
   a. _2141 Architects, town and traffic planners [KEEP]
   b. _2142 Civil engineers [KEEP]
   c. _2143 Electrical engineers [KEEP]
   d. _2144 Electronics and telecommunications engineers [KEEP]
   e. _2145 Mechanical engineers [KEEP]
   f. _2146 Chemical engineers [KEEP]
   g. _2147 Mining engineers, metallurgists and related professionals [KEEP]
   h. _2148 Cartographers and surveyors [KEEP]
   i. _2149 Architects, engineers and related professionals not elsewhere classified [KEEP]
9. Life science and health professionals [EXPANDABLE HEADER] [KEEP]
   a. _2210 Life science professionals [KEEP]
   b. _2220 Health professionals (except nursing) [KEEP]
   c. _2230 Nursing and midwifery professionals [KEEP]

   d. _2300 Teaching professionals [KEEP]
10. Other professionals [EXPANDABLE HEADER] [KEEP]
   a. _2410 Business professionals [KEEP]
   b. _2420 Legal professionals [KEEP]
   c. _2430 Archivists, librarians and related information professionals [KEEP]
   d. _2440 Social science and related professionals [KEEP]
   e. _2450 Writers and creative or performing artists [KEEP]
   f. _2460 Religious professionals [KEEP]
11. Physical and engineering science associate professionals [EXPANDABLE HEADER] [KEEP]
   a. _3110 Physical and engineering science technicians [KEEP]
   b. _3120 Computer associate professionals [KEEP]
   c. _3130 Optical and electronic equipment operators [KEEP]
   d. _3140 Ship and aircraft controllers and technicians [KEEP]
   e. _3150 Safety and quality inspectors [KEEP]
   f. _3200 Life science and health associate professionals [KEEP]
   g. _3300 Teaching associate professionals [KEEP]
12. Other associate professionals [EXPANDABLE HEADER] [KEEP]
   a. _3410 Finance and sales associate professionals [KEEP]
   b. _3420 Business services agents and trade brokers [KEEP]
   c. _3430 Administrative associate professionals [KEEP]
   d. _3440 Customs, tax and related government associate professionals [KEEP]
   e. _3450 Police inspectors and detectives [KEEP]
   f. _3460 Social work associate professionals [KEEP]
   g. _3470 Artistic, entertainment and sports associate professionals [KEEP]
   h. _3480 Religious associate professionals [KEEP]
13. Clerks [EXPANDABLE HEADER] [KEEP]
   a. _4100 Office clerks [KEEP]
   b. _4200 Customer services clerks [KEEP]
14. Personal and protective services workers [EXPANDABLE HEADER] [KEEP]
   a. _5110 Travel attendants and related workers [KEEP]
   b. _5120 Housekeeping and restaurant services workers [KEEP]
   c. _5130 Personal care and related workers [KEEP]
   d. _5140 Other personal services workers [KEEP]
   e. _5160 Protective services workers [KEEP]
   f. _5200 Models, salespersons and demonstrators [KEEP]
15. Skilled agricultural and fishery workers [EXPANDABLE HEADER] [KEEP]
   a. _6000 Skilled agricultural and fishery workers [KEEP]
16. Craft and related trades workers [EXPANDABLE HEADER] [KEEP]
   a. _7100 Extraction and building trades workers [KEEP]
17. Metal, machinery and related trades workers [EXPANDABLE HEADER] [KEEP]
   a. _7210 Metal moulders, welders, sheet-metal workers, structural - metal preparers, and related trades workers [KEEP]
   b. _7220 Blacksmiths, tool-makers and related trades workers [KEEP]
   c. _7230 Machinery mechanics and fitters [KEEP]
   d. _7240 Electrical and electronic equipment mechanics and fitters [KEEP]
18. Precision, handicraft, printing and related trades workers [EXPANDABLE HEADER] [KEEP]
   a. _7310 Precision workers in metal and related materials [KEEP]

    b. _7320 Potters, glass-makers and related trades workers [KEEP]
    c. _7330 Handicraft workers in wood, textile, leather and related materials [KEEP]
    d. _7340 Printing and related trades workers [KEEP]
19. Other craft and related trades workers [EXPANDABLE HEADER] [KEEP]
    a. _7410 Food processing and related trades workers [KEEP]
    b. _7420 Wood treaters, cabinet-makers and related trades workers [KEEP]
    c. _7430 Textile, garment and related trades workers [KEEP]
    d. _7440 Pelt, leather and shoemaking trades workers [KEEP]
20. Plant and machine operators and assemblers [EXPANDABLE HEADER] [KEEP]
    a. _8000 Plant and machine operators and assemblers [KEEP]
21. Elementary occupations [EXPANDABLE HEADER] [KEEP]
    a. _9100 Sales and services elementary occupations [KEEP]
    b. _9200 Agricultural, fishery and related labourers [KEEP]
    c. _9300 Labourers in mining, construction, manufacturing and transport [KEEP]
22. Armed forces [EXPANDABLE HEADER] [KEEP]
    a. _9888 Armed forces [KEEP]
23. Did not work before [EXPANDABLE HEADER] [KEEP]
    a. _9991 Unemployed and not looking for a job / Long-term sick or disabled [KEEP]
    b. _9992 Pupil /Student/ in full time education [KEEP]
    c. _9993 Housewife [KEEP]

*ASK IF IN UK AND EU01HINC =3*
*[Standard Screener: DO NOT MODIFY OR TRANSLATE]*

[PN: Part of the occupation module, cannot be asked independent. Asked only if EU01HINC Q4=3. When respondent is also main earner (EU01HINC Q4=1 or 2), then values recorded in UK01OCCR are automatically transferred to UK01OCCHI. ]

*IF Qsample=1 HIDE AND IMPORT DATA, CREATE RECODE VARIABLE WITH THE NAME UK01OCCHI.*
*IF Qsample=2 ASK UK01OCCHI.*

*SA*
*UK01OCCHI.*

What is the occupation of the person with the highest income? If retired or unemployed, please indicate the category closest to his/her previous occupation.

Select only one

1. USE UK01OCCHI response list
2. Legislators, senior officials and managers [EXPANDABLE HEADER] [KEEP]
    a. _1100 Legislators and senior officials [KEEP]
3. Corporate managers [EXPANDABLE HEADER] [KEEP]
    a. _1210 Directors and chief executives [KEEP]
4. Production and operations department managers [EXPANDABLE HEADER] [KEEP]
    a. _1221 Production and operations department managers in agriculture, hunting, forestry and fishing [KEEP]
    b. _1222 Production and operations department managers in manufacturing [KEEP]
    c. _1223 Production and operations department managers in construction [KEEP]

    d. _1224 Production and operations department managers in wholesale and retail trade [KEEP]
    e. _1225 Production and operations department managers in restaurants and hotels [KEEP]
    f. _1226 Production and operations department managers in transport, storage and communications [KEEP]
    g. _1227 Production and operations department managers in business services [KEEP]
    h. _1228 Production and operations department managers in personal care, cleaning and related services [KEEP]
    i. _1229 Production and operations department managers not elsewhere classified [KEEP]
5. Other department managers [EXPANDABLE HEADER] [KEEP]
    a. _1231 Finance and administration department managers [KEEP]
    b. _1232 Personnel and industrial relations department managers [KEEP]
    c. _1233 Sales and marketing department managers [KEEP]
    d. _1234 Advertising and public relations department managers [KEEP]
    e. _1235 Supply and distribution department managers [KEEP]
    f. _1236 Computing services department managers [KEEP]
    g. _1237 Research and development department managers [KEEP]
    h. _1239 Other department managers not elsewhere classified [KEEP]
6. General managers [EXPANDABLE HEADER] [KEEP]
    a. _1311 General managers in agriculture, hunting, forestry/ and fishing [KEEP]
    b. _1312 General managers in manufacturing [KEEP]
    c. _1313 General managers in construction [KEEP]
    d. _1314 General managers in wholesale and retail trade [KEEP]
    e. _1315 General managers of restaurants and hotels [KEEP]
    f. _1316 General managers in transport, storage and communications [KEEP]
    g. _1317 General managers of business services [KEEP]
    h. _1318 General managers in personal care, cleaning and related services [KEEP]
    i. _1319 General managers not elsewhere classified [KEEP]
7. Physical, mathematical and engineering science professionals [EXPANDABLE HEADER] [KEEP]
    a. _2110 Physicists, chemists and related professionals [KEEP]
    b. _2120 Mathematicians, statisticians and related professionals [KEEP]
    c. _2130 Computing professionals [KEEP]
8. Architects, engineers and related professionals [EXPANDABLE HEADER] [KEEP]
    a. _2141 Architects, town and traffic planners [KEEP]
    b. _2142 Civil engineers [KEEP]
    c. _2143 Electrical engineers [KEEP]
    d. _2144 Electronics and telecommunications engineers [KEEP]
    e. _2145 Mechanical engineers [KEEP]
    f. _2146 Chemical engineers [KEEP]
    g. _2147 Mining engineers, metallurgists and related professionals [KEEP]
    h. _2148 Cartographers and surveyors [KEEP]
    i. _2149 Architects, engineers and related professionals not elsewhere classified [KEEP]
9. Life science and health professionals [EXPANDABLE HEADER] [KEEP]
    a. _2210 Life science professionals [KEEP]
    b. _2220 Health professionals (except nursing) [KEEP]
    c. _2230 Nursing and midwifery professionals [KEEP]

d. _2300 Teaching professionals [KEEP]
10. Other professionals [EXPANDABLE HEADER] [KEEP]
    a. _2410 Business professionals [KEEP]
    b. _2420 Legal professionals [KEEP]
    c. _2430 Archivists, librarians and related information professionals [KEEP]
    d. _2440 Social science and related professionals [KEEP]
    e. _2450 Writers and creative or performing artists [KEEP]
    f. _2460 Religious professionals [KEEP]
11. Physical and engineering science associate professionals [EXPANDABLE HEADER] [KEEP]
    a. _3110 Physical and engineering science technicians [KEEP]
    b. _3120 Computer associate professionals [KEEP]
    c. _3130 Optical and electronic equipment operators [KEEP]
    d. _3140 Ship and aircraft controllers and technicians [KEEP]
    e. _3150 Safety and quality inspectors [KEEP]
    f. _3200 Life science and health associate professionals [KEEP]
    g. _3300 Teaching associate professionals [KEEP]
12. Other associate professionals [EXPANDABLE HEADER] [KEEP]
    a. _3410 Finance and sales associate professionals [KEEP]
    b. _3420 Business services agents and trade brokers [KEEP]
    c. _3430 Administrative associate professionals [KEEP]
    d. _3440 Customs, tax and related government associate professionals [KEEP]
    e. _3450 Police inspectors and detectives [KEEP]
    f. _3460 Social work associate professionals [KEEP]
    g. _3470 Artistic, entertainment and sports associate professionals [KEEP]
    h. _3480 Religious associate professionals [KEEP]
13. Clerks [EXPANDABLE HEADER] [KEEP]
    a. _4100 Office clerks [KEEP]
    b. _4200 Customer services clerks [KEEP]
14. Personal and protective services workers [EXPANDABLE HEADER] [KEEP]
    a. _5110 Travel attendants and related workers [KEEP]
    b. _5120 Housekeeping and restaurant services workers [KEEP]
    c. _5130 Personal care and related workers [KEEP]
    d. _5140 Other personal services workers [KEEP]
    e. _5160 Protective services workers [KEEP]
    f. _5200 Models, salespersons and demonstrators [KEEP]
15. Skilled agricultural and fishery workers [EXPANDABLE HEADER] [KEEP]
    a. _6000 Skilled agricultural and fishery workers [KEEP]
16. Craft and related trades workers [EXPANDABLE HEADER] [KEEP]
    a. _7100 Extraction and building trades workers [KEEP]
17. Metal, machinery and related trades workers [EXPANDABLE HEADER] [KEEP]
    a. _7210 Metal moulders, welders, sheet-metal workers, structural - metal preparers, and related trades workers [KEEP]
    b. _7220 Blacksmiths, tool-makers and related trades workers [KEEP]
    c. _7230 Machinery mechanics and fitters [KEEP]
    d. _7240 Electrical and electronic equipment mechanics and fitters [KEEP]
18. Precision, handicraft, printing and related trades workers [EXPANDABLE HEADER] [KEEP]
    a. _7310 Precision workers in metal and related materials [KEEP]

    b. _7320 Potters, glass-makers and related trades workers [KEEP]
    c. _7330 Handicraft workers in wood, textile, leather and related materials [KEEP]
    d. _7340 Printing and related trades workers [KEEP]
19. Other craft and related trades workers [EXPANDABLE HEADER] [KEEP]
    a. _7410 Food processing and related trades workers [KEEP]
    b. _7420 Wood treaters, cabinet-makers and related trades workers [KEEP]
    c. _7430 Textile, garment and related trades workers [KEEP]
    d. _7440 Pelt, leather and shoemaking trades workers [KEEP]
20. Plant and machine operators and assemblers [EXPANDABLE HEADER] [KEEP]
    a. _8000 Plant and machine operators and assemblers [KEEP]
21. Elementary occupations [EXPANDABLE HEADER] [KEEP]
    a. _9100 Sales and services elementary occupations [KEEP]
    b. _9200 Agricultural, fishery and related labourers [KEEP]
    c. _9300 Labourers in mining, construction, manufacturing and transport [KEEP]
22. Armed forces [EXPANDABLE HEADER] [KEEP]
    a. _9888 Armed forces [KEEP]
23. Did not work before [EXPANDABLE HEADER] [KEEP]
    a. _9991 Unemployed and not looking for a job / Long-term sick or disabled [KEEP]
    b. _9992 Pupil /Student/ in full time education [KEEP]
    c. _9993 Housewife [KEEP]

UK01SG [Hidden]. Hidden Question:

Social Grade

1. _1 A - Upper middle class [KEEP]
2. _2 B - Middle class [KEEP]
3. _3 C1 - Lower middle class [KEEP]
4. _4 C2 - Skilled working class [KEEP]
5. _5 D - Working class [KEEP]
6. _6 E - Lower level of subsistence [KEEP]

*ASK IF GENDER_NONBINARY Q2=2 OR 3 AND RESPAGE<55*
*SA*
PREGNANCY.
Q15. Are you currently pregnant?

Select only one

1. _1 Yes
2. _2 No
3. _3 Prefer not to answer

UK02INC

The next question may be considered personal, but it is not mandatory to answer. If you do, we assure you that your responses will be kept strictly confidential and used for research purposes only.

What is the COMBINED TOTAL ANNUAL INCOME (pre-tax) earned by all members of your household?

Please include all your income sources : salaries, scholarships, pension and Social Security benefits, dividends from shares, income from rental properties, child support and alimony etc.

_1 "Under £5,000",
_2 "£5,000 - £9,999",
_3 "£10,000 - £14,999",
_4 "£15,000 - £19,999",
_5 "£20,000 - £24,999",
_6 "£25,000 - £34,999",
_7 "£35,000 - £44,999",
_8 "£45,000 - £54,999",
_9 "£55,000 - £99,999",
_10 "£100,000 or more",
_11 "Prefer not to answer"

UK02ETH.
Some questions can be sensitive in nature. We would like to remind you that your participation is strictly voluntary and that your responses are used for research purposes only. A "Prefer not to answer" option is available for you to select, if the case.

What is your ethnic group?
Select only one
White [Expandable Header]
_1 English / Welsh / Scottish / Northern Irish / British
_2 Irish
_3 Gypsy or Irish Traveller
_4 Any other White background
Mixed / multiple ethnic groups [Expandable Header]
_5 White and Black Caribbean
_6 White and Black African
_7 White and Asian
_8 Any other Mixed / multiple ethnic background
Asian / Asian British [Expandable Header]
_9 Indian
_10 Pakistani
_11 Bangladeshi
_12 Chinese
_13 Any other Asian background
Black / African / Caribbean / Black British [Expandable Header]
_14 African

_15 Caribbean
_16 Any other Black / African / Caribbean background
Other ethnic group [Expandable Header]
_17 Arab
_18 Any other ethnic group, please write in ____________
_19 Prefer not to answer

*IF QSAMPLE=1 1 HIDE AND IMPORT DATA, CREATE RECODE VARIABLE WITH THE NAME Q21IMPORT*
*IF QSAMPLE=2 AND Q20=2 OR MORE HAVE ONE LESS ROW THAN THE NUMBER SELECTED AT Q20 FOR EACH PERSON IN QHOUSE I.E. IF SELECTED TWO PEOPLE IN THE HOUSE AT Q20 SHOW ONE ROW*
*LIMIT TO 10 ANSWER BOXES IF QHOUSE=11*
*WRITE IN*
QNAME
Q21. Please write the nickname or initials of each **other** person in your household.
Note that this nickname is only needed to make it easier for you to complete the survey, this nickname will appear in the following questions about your household, so please pick a nickname that will help you identify each household member. Nicknames are not visible to anyone outside of this survey.

Please write in
1. NAME1
2. NAME2
3. NAME3
etc.

*ASK THE FOLLOWING LOOP Q23 TO Q27 FOR EACH NAME GIVEN AT Q21 Q21IMPORT (EXCLUDE NAMES SELECTED AT HHCOMPRREMOVE) AND EACH OF NAMES GIVEN AT HHCOMPADD*
*SA*

QHHAGE

Q23. Which of the following age groups do they fit into?

1. Under 1
2. 1-4
3. 5-11
4. 12-15
5. 16-17
6. 18-19
7. 20-24
8. 25-34
9. 35-44
10. 45-54
11. 55-64
12. 65-69
13. 70-74
14. 75-79
15. 80-84
16. 85+
17. Don't know
18. Prefer not to answer

*IF sampletype=2 and QHHAGE DOES NOT EQUAL 1, 2, 3, 4 OR 5 FOR ANY NAME GIVEN AT Q21, THANK AND CLOSE*

*IF Qsample=1 AND HHCOMPCONFIRM=3 ASK FOR EACH NAME AT HHCOMPADD ASK QHHGENDER*
*IF Qsample=2 ASK QHHGENDER*

ASK ALL
*SA*
QHHGENDER
Q24.
As far as you know, which of the following describes how [NAME] thinks of themselves?
Select only one

1. _1 Male [KEEP]
2. _2 Female [KEEP]
3. _3 In another way [KEEP]
4. _4 Prefer not to answer [KEEP]
5. _5 Don't know

*CONCATANATE Q21, Q23, Q24 TO CREATE NAMEAGESEX FOR EACH HH MEMBER*
*Format: Insert Q21 (Insert Q23, InsertQ24)*

1. *NAMEAGESEX1*
2. *NAMEAGESEX2*
3. *NAMEAGESEX3*

*ONLY ASK THOSE AGED 16 AND OVER AT QHHAGE INCLUDING IF CODES 18 AND 19. IF QSAMPLE=1 EXCLUDE NAMES SELECTED AT HHCOMPREMOVE AND ASK EACH OF NAMES GIVEN AT HHCOMPADD*
*SA*
Q25.
What is [NAME]'s current employment status?
Select only one

1. _1 Employed full-time (34 hours or more) [KEEP]
2. _2 Employed part-time (less than 34 hours) [KEEP]
3. _3 Self employed [KEEP]
4. _4 Unemployed but looking for a job [KEEP]
5. _5 Unemployed and not looking for a job [KEEP]
6. _6 Full-time parent, homemaker [KEEP]
7. _7 Retired [KEEP]
8. _8 Student/Pupil [KEEP]
9. _9 Long-term sick or disabled [KEEP]

*SA*
*ASK IF q23=CODES 1-18. IF QSAMPLE=1 EXCLUDE NAMES SELECTED AT HHCOMPREMOVE AND ASK EACH OF NAMES GIVEN AT HHCOMPADD*

QHHATTEND
Q26. Does [NAME] normally attend or will they be attending in the new school term any of the following as a pupil or student?
Select only one
1. Nursery or pre-school [*UK ONLY SHOW THIS OPTION FOR THOSE AGED 11 OR UNDER AT QHHAGE]*
2. School [*UK ONLY SHOW THIS OPTION FOR THOSE AGED 1 TO 24 AND CODES 17 AND 18 AT QHHAGE]*
3. Further education, e.g. college [*SHOW THIS OPTION FOR THOSE AGED 12 AND OVER AND CODES 17 AND 18 AT QHHAGE]*
4. Higher education, e.g. university [*SHOW THIS OPTION FOR THOSE AGED 12 AND OVER OR CODES 17 AND 18 AT QHHAGE]*
5. None of the above
6. Don't know
7. Prefer not to answer

*SHOW TEXT FOR ALL*

These questions are voluntary but they are really important in helping us understand the spread of COVID-19 and the impact of different public health interventions. It will not be possible to identify you or any member of your household in the published findings.

*ASK FOR SELF AND EACH PERSON NAMED AT Q21/Q21 IMPORT. IF QSAMPLE=1 AND HHCOMPCONFRIM =2 THEN REMOVE ANYONE SELECTED AT HHCOMPREMOVE. QSAMPLE=1 AND HHCOMPCONFRIM =3 THEN ADD PEOPLE FROM HHCOMPADD*
*SA*
*HIGHRISK*
Q28a Are you or any other household member in a high risk group, meaning you could have serious symptoms if you contracted Coronavirus (COVID-19)?

*High risk groups include individuals who: have had an organ transplant, undergoing cancer treatment, have blood or bone marrow cancer, have had a bone marrow or stem cell transplant in the past 6 months, are taking immunosuppressant medicine or high doses of steroids, have a severe lung condition (such as cystic fibrosis, severe asthma or severe COPD), have a condition that makes risk of getting infections higher (e.g. SCID or sickle cell), and/or are pregnant and have a serious heart condition*

***ROWS:***
0. Yourself
1. Name 1
2. Name 2

1. Yes
2. No
3. Prefer not answer

*ASK IF Q28a =2*
*MEDRISK*
*SA*
Q28b Are you or any other household member in a medium risk group, meaning you could have serious symptoms if you contracted Coronavirus (COVID-19)?

*Medium risk groups include individuals who: have a lung condition that is not severe (such as asthma, COPD, emphysema or bronchitis), have heart disease (e.g. heart failure), have diabetes, have chronic, kidney or liver disease, have a condition affecting the brain or nerves (such as Parkinson's disease, motor neurone disease, multiple sclerosis or cerebral palsy), have a condition that means they have a high risk of getting infections, are taking medicine that can affect the immune system (such as low doses of steroids), have a BMI of 40 or above, and/or are pregnant*

*IF SAMPLE TYPE=1*
***ROWS:***
0. Yourself
1. Name 1
2. Name 2

1. Yes
2. No
3. Prefer not answer

**Symptoms**

*ASK FOR SELF AND EACH PERSON NAMED AT Q21/Q21 IMPORT. IF QSAMPLE=1 AND HHCOMPCONFRIM =2 THEN REMOVE ANYONE SELECTED AT HHCOMPREMOVE. QSAMPLE=1 AND HHCOMPCONFRIM =3 THEN ADD PEOPLE FROM HHCOMPADD*

*MA*

SYMPTOMS

Q29.

Have you or anyone else in your household had any of the following symptoms in the **last seven days**?

Please tick all that apply for each row

*SET UP AS PROGRESSIVE GRID*

*ROWS:*

0. Yourself
1. Name 1
2. Name 2

etc.

Please tick all that apply

*ROTATE LIST*

1. Fever or chills
2. Cough
3. Shortness of breath or difficulty breathing
4. Fatigue (extreme tiredness)
5. Muscle or body aches
6. Headache
7. New loss of taste or smell
8. Sore throat
9. Congestion or runny nose
10. Nausea or vomiting
11. Diarrhoea
12. None of these [EXCLUSIVE]
13. Don't know [EXCLUSIVE]
14. Prefer not to answer [EXCLUSIVE]

**Child selection**

*ASK IF QSAMPLETYPE=2 AND ALL WHO HAVE MORE THAN ONE CHILD AGED UNDER 18 AT Q23 (MORE THAN 1 CODE 1-5 AT Q23) OR IF ONE CHILD AGED UNDER 18 AT Q23 INSERT THAT CHILD*

*QP53a*

*SA*

*SELECT CHILD*

We need to choose one child at random for you to think about when answering the remaining questions. To do this, please tell us which of your children will have their birthday next. If you have two or more children with birthdays on the same day, please select the one whose name comes first alphabetically. Please only think about the children you are a parent to.

*LIST ALL CHILDREN FROM Q21 IF THEY ARE UNDER THE AGE OF 18 (CODES 1-5) AT Q23*

*Note to scripter – can you list the concatenated variable NAMEAGESEX rather than answer from Q21*

*SHOW TEXT FOR ALL*
When answering the following questions, please answer only on behalf of [*IF MORE THAN ONE CHILD AGED UNDER 18 AT Q23 INSERT CHILD'S NAME FROM QP53a OR IF ONE CHILD AGED UNDER 18 AT Q23 INSERT CHILD'S NAME FROM Q21]*. Please answer to the best of your ability, with your best guess if you do not know the exact answer.

*IF QSAMPLE=1*

*SHOW TEXT FOR ALL*
*IF QSAMPLE=1 THEN ASK ALL SUBSEQUENT QUESTIONS Q28, Q29, Q32, , Q32a, Q39, Q41, Q42, Q52, QP54, QP59, QP60, QP61a, QP61b, QP62, QP63, QP66, QP67, QP68, QP69, QP70, QP71, QP72, QP74, Qp79, Qp80, Qp81 c ABOUT CHILD SELECTED AT INITIAL WAVE in QP53a OR IF ONE CHILD AGED UNDER 18 AT Q23 INSERT CHILD'S NAME FROM Q21*

*If person selected at initial wave is removed at hhcompremove, then make a new allocation*

Last time you completed this survey for us you told us about INSERT CHILD'S NAME from QP53a
When answering the following questions, please answer only on behalf of INSERT CHILD'S NAME from QP53a]. Please answer to the best of your ability, with your best guess if you do not know the exact answer.

*SHOW TEXT FOR ALL*

These questions are voluntary but they are really important in helping us understand the spread of COVID-19 and the impact of different public health interventions. It will not be possible to identify you or any member of your household in the published findings.

**Vaccination**
*IF QSAMPLE=2 OR (QSAMPLE=1 AND RESPONSE TO QXX1/QPXX1 AT ANY PRIOR WAVE =2 or has never been asked) THEN ASK QXX1 TO QXX3/QPXX1 TO QPXX3*
*SA*
Vaccination
*IF SAMPLE TYPE=1*
QXX1. Have you been vaccinated against the virus that causes Coronavirus (COVID-19)?
*IF SAMPLE TYPE =2*
QPXX1. Has [NAME] been vaccinated against the virus that causes Coronavirus (COVID-19)?
1. Yes
2. No
3. Don't know [SHOW FOR *IF SAMPLE TYPE =2* ONLY]

SA
If QXX1 or QPXX1 = "YES"
*IF SAMPLE TYPE=1*
QXX2.

How many doses of vaccinations have you had against the virus that causes Coronavirus (COVID-19)?
*IF SAMPLE TYPE =2*
QPXX2.
How many doses of vaccinations has [NAME] had against the virus that causes Coronavirus (COVID-19)?
WRITE IN NUMBER OF DOSES: [PN: RANGE FROM 1 TO 5]
Don't know [SHOW FOR *IF SAMPLE TYPE =2* ONLY]

If QXX1 or QPXX1 = "YES"
QXX3.
Please provide the dates you received your vaccination(s) against the virus that causes COVID-19.

*Please records the date(s) in the following format – dd/mm/yyyy*

PN: INSERT NUMBER OF DATE BOXES THAT MATCHES ANSWER AT QXX2
INSERT DATE_BOX
INSERT DATE_BOX
Don't know

QPXX3.
Please provide the dates [NAME] received their vaccination(s) against the virus that causes COVID-19.

*Please records the date(s) in the following format – dd/mm/yyyy*

PN: INSERT NUMBER OF DATE BOXES THAT MATCHES ANSWER AT QPXX2
INSERT DATE_BOX
INSERT DATE_BOX
Don't know

*QSAMPLE=1 AND RESPONSE TO QXX1/QPXX1 AT ANY PRIOR WAVE =1 AND ASK QZZ1 TO QZZ3/QPZZ1 TO QPZZ3*
*SA*
Vaccination
*IF SAMPLE TYPE=1*
QZZ1. You previously said you have been vaccinated against the virus that causes Coronavirus (COVID-19). Have you had any new doses of the vaccination since you last completed the survey?
*IF SAMPLE TYPE =2*
QPZZ1. You previously said [NAME] had been vaccinated against the virus that causes Coronavirus (COVID-19). Has [NAME] had any new doses of the vaccination since you last completed the survey?
1. Yes
2. No
3. Don't know [SHOW FOR *IF SAMPLE TYPE =2* ONLY]

SA
If QZZ1 or QPZZ1 = "YES"
*IF SAMPLE TYPE=1*

QZZ2.
How many new doses of vaccinations have you had against the virus that causes Coronavirus (COVID-19)?
*IF SAMPLE TYPE =2*
QPZZ2.
How many new doses of vaccinations has [NAME] had against the virus that causes Coronavirus (COVID-19)?
WRITE IN NUMBER OF DOSES: [PN: RANGE FROM 1 TO 5]
Don't know [SHOW FOR *IF SAMPLE TYPE =2* ONLY]

If QZZ1 or QPZZ1 = "YES"
QZZ3.
Please provide the dates you received your new vaccination(s) against the virus that causes COVID-19.
PN: INSERT NUMBER OF DATE BOXES THAT MATCHES ANSWER AT QZZ2
INSERT DATE_BOX
INSERT DATE_BOX
Don't know

QPZZ3.
Please provide the dates [NAME] received their new vaccination(s) against the virus that causes COVID-19.
PN: INSERT NUMBER OF DATE BOXES THAT MATCHES ANSWER AT QPZZ2
INSERT DATE_BOX
INSERT DATE_BOX
Don't know

*ASK ALL*
*SA*
*IF SAMPLETYPE = 1* Q32. Have you been tested for Coronavirus (Covid-19) via a nose and/or throat swab in the **last fourteen days**?
*IF SAMPLETYPE = 2* Q32. Has NAME been tested for Coronavirus (Covid-19) via a nose and/or throat swab in the **last fourteen days**?

1. Tested and the test showed I *(SAMPLETYPE=1)/*they *(SAMPLETYPE=2)* have Coronavirus currently
2. Tested, and the test showed I*(SAMPLETYPE=1)/*they *(SAMPLETYPE=2)* do not have Coronavirus currently
3. Yes, and I'm *(SAMPLETYPE=1)/*they are *(SAMPLETYPE=2)* still waiting to hear the result
4. Not tested
5. Don't know
6. Prefer not to answer

*SA*
TEST
*ASK IF WAVE=1 AND IF SAMPLETYPE = 1* Q32a. Have you been tested for Coronavirus (Covid-19) via a nose and/or throat swab **prior to the past fourteen days?**
*ASK IF WAVE=1 AND IF SAMPLETYPE = 2* Q32a Has [NAME] been tested for Coronavirus (Covid-19) via a nose and/or throat swab **prior to the past fourteen days?**

*IF Qsample=1 HIDE AND IMPORT DATA*
*IF Qsample=2 ASK Q32a*

1. Tested and the test showed I *(SAMPLETYPE=1)/*they *(SAMPLETYPE=2)* did have Coronavirus at the time
2. Tested, and the test showed I*(SAMPLETYPE=1)/*they *(SAMPLETYPE=2)* <u>did not</u> have Coronavirus
3. Yes, and I'm *(SAMPLETYPE=1)/*they are *(SAMPLETYPE=2)* still waiting to hear the result
4. Not tested
5. Don't know
6. Prefer not to answer

*ASK ALL*
SA
*IF SAMPLETYPE = 1* Q32b. Have you ever been tested for Coronavirus (Covid-19) via a blood test (antibodies test)?
*IF SAMPLETYPE = 2* Q32b. Has [NAME] ever been tested for for Coronavirus (Covid-19) via a blood test (antibodies test)?

1 Tested and the test showed that I *(SAMPLETYPE=1)/*they *(SAMPLETYPE=2)* have or had Coronavirus
2 Tested and the test showed that I *(SAMPLETYPE=1)/*they *(SAMPLETYPE=2)* have not had Coronavirus
3. Yes, and I'm *(SAMPLETYPE=1)/*they are *(SAMPLETYPE=2)* still waiting to hear the result
4. Not tested
5. Don't know
6. Prefer not to answer

**Attitudes**

*ASK ALL WHERE SAMPLETYPE=1*
*SA*
ATT1
Q35. To what extent do you agree or disagree with each of the following statements?
*REVERSE ORDER OF SCALE. ROTATE STATEMENTS. SCRIPT AS PROGRESSIVE GRID. SA PER COLUMN.*
Select only one for each row

1. Strongly agree
2. Tend to agree
3. Neither agree nor disagree

4. Tend to disagree
5. Strongly disagree
6. Don't know

1. Coronavirus would be a serious illness for me
2. I am likely to catch coronavirus
3. I'm worried that I might spread coronavirus to someone who is vulnerable

**Behaviour**
*ASK ALL*
*ISOLATION*
*SA*
*IF SAMPLETYPE=1* Q39 In the last seven days have you been in isolation or quarantine due to coronavirus (Covid-19)?
*IF SAMPLETYPE=2* Q39 In the last seven days has [NAME] been in isolation or quarantine due to coronavirus (Covid-19)?
*This includes whether you have stayed at home after potential exposure to an infected case, or were asked to quarantine on return from a trip abroad, or separated yourself from people who are not infected, including any household members. You can be in isolation in your house or in a health facility.*

1. Yes
2. No
3. Prefer not to say

*ASK Q41A,B,C IN A LOOP*
*ASK (IF Q3=1,2,3 or Q3c=1 or 2) for sampletype=1 or Q25=1,2,3 for the selected child if sampletype=2*
SA
Q41a. In the last seven days, was [*IF SAMPLTYPE=1 –* your] [*IF SAMPLETYPE=2* NAME'S*]* workplace…

1. Fully open as normal
2. Partially open i.e. open some days, or for certain hours, or only open to some members of staff
6. Not open to any members of staff
3. Not applicable – I/they do not have a workplace
4. Don't know
5. Prefer not to answer

ASK IF Q41a=1 or 2
SA
Q41b. And, in the last seven days how often did [*IF SAMPLETYPE=1 –* you] [*IF SAMPLETYPE=2* NAME'S*]* go into [*IF SAMPLETYPE=1 –* your] [*IF SAMPLETYPE=2* their*]* workplace?

1. Every day
2. Most days
3. About once or twice per week
4. No days – they/I only worked from home
5. No days – they/I did not work this week for another reason

6. Don't know
7. Prefer not to answer

ASK IF Q41b=1,2,3
SA
Q41c. Did [*IF SAMPLETYPE=1 –* you] [*IF SAMPLETYPE=2* NAME'S] go into your/their workplace yesterday?

1. Yes
2. No
3. Prefer not to answer

*ASK IF Q3=8 or Q3b=1 or 2 for sampletype=1 OR Q26 = 1, 2, 3 OR 4 for the selected child if sampletype=2*
SA
Q42a. In the last seven days, was [*IF SAMPLTYPE=1 –* your] [*IF SAMPLETYPE=2* NAME'S] [university or college] [*SHOW IF Q3 = 8 OR Q3b=1 or 2 Q26 = 3 OR 4*] OR [pre- school or nursery] [*Q26 = 1]* OR [school] [Q3b =1 OR *Q26 = 2*]…

1. Fully open as normal
2. Partially open i.e. open some days, or for certain hours, or only open to some
6. Not open to any children, pupils, or students
children/pupils/students
3. Not applicable – it is closed for the holidays
4. Don't know
5. Prefer not to answer

ASK IF Q42a=1 or 2
SA
Q42b. And, in the last seven days how often did [*IF SAMPLTYPE=1 –* you] [*IF SAMPLETYPE=2* NAME] go into [university or college] [*SHOW IF Q3 = 8 OR Q3b= 1 or 2 OR Q26 = 3 OR 4*] OR [pre- school or nursery] [*Q26 = 1]* OR [school] [*Q26 = 2*]…

1. Every day
2. Most days
3. About once or twice per week
4. No days – [*IF SAMPLTYPE=1 –* I] [*IF SAMPLETYPE=2* NAME] only did [*IF SAMPLETYPE=2* their] [university or college] [*SHOW IF Q3 = 8 OR Q3b=1 or 2 Q26 = 3 OR 4*] OR [pre- school or nursery] [*Q26 = 1]* OR [school] [*Q26 = 2*] work/activities at home
5. No days - [*IF SAMPLTYPE=1 –* I] [*IF SAMPLETYPE=2* NAME] did not attend [university or college] [*SHOW IF Q3 = 8 OR Q3b=1 or 2 OR Q26 = 3 OR 4*] OR [pre- school or nursery] [*Q26 = 1]* OR [school] [*Q26 = 2*] for another reason
6. Don't know
7. Prefer not to answer

ASK IF SAMPLETYPE=1 AND Q42b=1,2,3
SA

Q42c. And, did you go into university or college yesterday?

1. Yes
2. No
3. Prefer not to answer

*ASK ALL*
*MA*
VISIT1
*IF SAMPLETYPE=1* Q52. Did you visit or do any of the following in the last seven days?
*SAMPLETYPE=2* Q52. Did [NAME] visit or do any of the following in the last seven days?

Please tick all that apply

1. Someone else's house (including their garden)
2. A place of worship, eg church, mosque or temple
3. Any form of public transport
4. A shop or market for essentials, eg a supermarket, grocery store, pharmacist
5. A shop for non-essential items, eg a DIY shop, or a clothes or furniture shop
6. A place of entertainment such as a restaurant, pub, cinema
7. A place for sports such as a gym or sports club/match
8. Outside, for example in a park, on the street or in the countryside
9. A healthcare setting, eg hospital, GP, A&E, outpatient facility, dentist, physiotherapist, optometrist, etc
10. A hair dresser, barber, nail salon, beauty parlor or similar location
11. None of the above [EXCLUSIVE]

***<u>Main sample Survey questions</u>***

**Individual preventive measures**

*ASK IF SAMPLETYPE=1*
*SA*
MASK1
Q54. Did you use a face mask yesterday?
Select only one
1. Yes
2. No

*ASK ALL WHO USED A FACE MASK YESTERDAY (Q54=1)*
*MA*
MASK3
Q56. Where did you use your face mask between 5am yesterday and 5am today?
Please tick all that apply
1. Everywhere outside my house [*SINGLE CODE EXCEPT WHEN SELECTED WITH CODE 8/ IF CODE 1 & 8 SELECTED, MAKE EXCLUSIVE – NO OTHER CODES ALONG WITH 1 & 8*]
2. When on the street
3. When cycling
4. On public transport
5. In supermarkets/shops
6. In cinema/bar/restaurant
8. At home
11. At someone else's house
9. At work/school/college/university
10. In the park/outside
7. Other (please specify)

*ASK ALL IF SAMPLETYPE=1*

*MA*
TRANSPORT
Q59. Did you travel on any public transport yesterday?
Please tick all that apply

1. No *[SINGLE CODE]*
2. Train/tube
3. Bus/tram
4. Taxi, Minicab, Uber, or similar ride-hailing app
5. Aeroplane
6. Ferry/boat

**Contact survey**

*THIS SECTION IS TO BE ASKED ONLY THOSE WHERE QSAMPLETYPE=1 (this includes Q62, Q63, Q66, Q67, Q68, Q69, Q71, Q77, Q72, Q73, Q74, Q79a-c, Q80a-c, Q81a-c).*

*ASK ALL*
*INTRO SCREEN*
We will now ask you to remember who you have been in contact with yesterday, between 5am yesterday and 5am today.

These questions are voluntary but they are really important in helping us understand the spread of COVID-19 and the impact of different public health interventions. It will not be possible to identify you or any member of your household in the published findings.

We are only interested in direct contacts, which are **people who you met in person** and with whom you exchanged at least a few words, or with whom you had physical contact (e.g. a handshake, embracing, kissing, contact sports).
**Note that if you only spoke to someone over the phone or internet, they should not be included in this section.**

*ASK ALL*
*SA*
CONTACT1
Q62. Which of the following people did you have direct in-person contact with between 5am yesterday and 5am today, in person?
*NOTE FOR SCRIPTER – PLEASE SCRIPT AS GRID*

*Note for scripter to add (i) button for 'direct contact':*
*We are only interested in direct contacts, which are people who you met in person and with whom you exchanged at least a few words, or with whom you had physical contact (e.g. a handshake, embracing, kissing, contact sports).*
***Note that if you only spoke to someone over the phone or internet, they should not be included in this section.***

*ROWS:*
*INSERT ALL NAMES FROM Q21/Q21IMPORT*

*COLUMNS:*
1. Yes
2. No

*ASK ALL*
*WRITE IN*
CONTACT2
*INTRO SCREEN*

Q63. Now we would like to know what **other people, outside of your household,** you had direct in person contact with between 5am yesterday and 5am today. This could include friends, family, work colleagues, customers, patients or people you spoke to in shops and so on.
*We are only interested in people who you met in person and **with whom you exchanged at least a few words, or with whom you had physical contact** (e.g. a handshake, embracing, contact sports)**. Please include people who you talked with while keeping a two-metre distance, wearing a mask, or otherwise social distancing.***

*NEW SCREEN*

Please write the nickname of each person outside your household who you had direct in person contact with below. Note that this nickname is only needed to make it easier for you to complete the survey, so please pick a nickname that is easy to remember. Your individual responses will not be shared with anyone outside this survey.

It is easiest to list names in chronological order, e.g. After I had breakfast at home, I went to work where I met with Jack, Deborah, and I served two customers. On my way back home, I chatted with the shop assistant at the petrol station (give a nickname like "shop assistant"). When I returned home, I accepted a package from the delivery person, and I spoke to my friend, Fatima, in my garden. Etc.
**Please do not list yourself, anyone that you listed as being a part of your household, or anyone who you only spoke to over the phone or internet.**

*PN: SHOW EXAMPLES IF Q20 IS NOT ONE AND WHEN CONTACTS ARE SELECTED AT Q62]*
*You have already indicated contact with the following household members, add additional contacts in the textboxes below:*

LIST HOUSEHOLD MEMBERS SELECTED AT Q62

*PN AT TOP OF PAGE WITH TEXT BOXES INCLUDE THE FOLLOWING.*

*PLEASE LIST ALL OTHER CONTACTS YOU HAVE OUTSIDE OF YOUR HOUSEHOLD, FOR EXAMPLE, J, D, CUSTOMER 1, CUSTOMER 2, SHOP ASSISTANT, DELIVERY PERSON, F*

*ALLOW BOXES TO INPUT NICKNAMES*
*PN SHOW TO ALL PARTICIPANTS REGARDLESS OF PREVIOUS ANSWERS*

*[] No-one/No other people [PN. ALLOW THIS TO BE SELECTED WHEN TEXT BOXES COMPLETED AND Please show this at the top of open text boxes.]*

*PN SHOW 15 BOXES ON SCREEN AND ADD ADDITONAL BOXES ONE AT A TIME AS COMPLETED. LIMIT TO 100 BOXES IN TOTAL.*

*ASK THE FOLLOWING LOOP Q71, Q77, Q72, Q73, Q69 TO TIME FOR EACH NAME GIVEN AT Q62*

*ASK THE FOLLOWING LOOP Q66, Q67, Q68, Q71, Q77, Q72, Q73, Q69 TO TIME FOR EACH NAME GIVEN AT Q63*

*ASK FOR EACH PERSON AT Q63*
*SA*
Q66. Which of the following age groups does NAME fit into? Please give an estimate if you are not sure
Select only one

1. Under 1
2. 1-4
3. 5-9
4. 10-14
5. 15-19
6. 20-24
7. 25-34
8. 35-44
9. 45-54
10. 55-64
11. 65-69
12. 70-74
13. 75-79
14. 80-84
15. 85+
16. Don't know
17. Prefer not to answer
18. This person is me

*ASK ALL WHERE NAME GIVEN AT Q63 AND Q66 IS NOT EQUAL TO CODE 18*
*SA*
CONTACTGEN
Q67. As far as you know, which of the following describes how [NAME] thinks of themselves?
Select only one

1. _1 Male [KEEP]
2. _2 Female [KEEP]
3. _3 In another way [KEEP]
4. _4 Prefer not to answer [KEEP]
5. _5 Don't know

*ASK ALL WHERE NAME GIVEN AT Q63 AND Q66 IS NOT EQUAL TO CODE 18*
*SA*
CONTACTRELAT
Q68. What is [NAME]'s relationship to you?
Select only one

1. They are a family member who is not in my household
7. They are my spouse/girlfriend/boyfriend/partner
2. They are someone I work with

8. They are a client or customer
3. They are someone I go to school, college or university with
4. They are a friend
5. Other
6. Prefer not to answer

*ASK ALL WHERE NAME GIVEN AT Q62 OR Q63 AND Q66 IS NOT EQUAL TO CODE 18*
*MA*
WHERE
Q71. And where did you have direct contact with [NAME] between 5am yesterday and 5am today? *If you workplace falls under one of these options i.e. a shop or a healthcare setting please select this as well as "at work".*
Please tick all that apply

1. At home (including at your door, in your garden, and within entrances to your home such as stairways, lifts, and corridors)
2. At someone else's house
3. At work
4. At a place of worship eg church, mosque or temple
5. On any form of public transport
6. At university, school, pre-school, or nursery
7. At a shop or market for essentials, eg a supermarket, grocery store, pharmacist
8. At a shop for non-essential items, eg a DIY shop, or a clothes or furniture shop
9. At a place of entertainment such as a restaurant, bar, cinema
10. At a place for sports such as a gym or sports club/match
11. Outside, for example in a park, on the street or in the countryside
14. In a healthcare setting, eg hospital, GP, A&E, outpatient facility, dentist, physiotherapist, optometrist, etc
15. At a hair dresser, barber, nail salon, beauty parlor or similar location
12. Somewhere else (please specify)

*ASK ALL WHERE NAME GIVEN AT Q62 OR Q63 AND Q66 IS NOT EQUAL TO CODE 18*
*MA*
*DIRCONT*
Q77. Which of the following apply to the direct contact with [NAME] yesterday?
Select all that apply

1. You had physical contact (any sort of skin-to-skin contact e.g. hand shaking, embracing or kissing) [PN. DO NOT ALLOW TO SELECT WITH CODED 2, 3 AND 4]
2. You kept a distance of two or more metres apart the entire time you were with them [PN. DO NOT ALLOW TO SELECT WITH CODED 1, 3 AND 4]
3. You kept a distance of one metre apart the entire time you were with them [PN. DO NOT ALLOW TO SELECT WITH CODED 1,2 AND 4]
4. You were less than one metre apart the entire time you were with them [PN. DO NOT ALLOW TO SELECT WITH CODED 1, 2 AND 3]
5. You wore a mask the entire time you were with them
6. You washed/sanitised your hands directly before contact
7. You washed/sanitised your hands directly after contact
9. Prefer not to say [PN: exclusive]
10. None of these [PN: exclusive]

*ASK ALL WHERE NAME GIVEN AT Q62 OR Q63 AND Q66 IS NOT EQUAL TO CODE 18*
*SA*
TIME
Q72. Please estimate the total amount of time you spent with [NAME] in person yesterday, this can include time when you were not in close proximity.

1. Less than 5 minutes
2. 5 minutes or more, but less than 15 minutes
3. 15 minutes or more, but less than 1 hour
4. 1 hour or more, but less than 4 hours
5. 4 hours or more

*ASK ALL WHERE NAME GIVEN AT Q62 OR Q63 AND Q66 IS NOT EQUAL TO CODE 18 AND IF Q71= 1, 2, 3, 4, 6, 7, 9, 10 or 12, 14 or 15*
MA
Q73 Was the time you spent with [NAME] yesterday inside and/or outside?
Please tick all that apply
Inside
Outside

*ASK ALL WHERE NAME GIVEN AT Q62 OR Q63 AND Q66 IS NOT EQUAL TO CODE 18*
*SA*
CONTACTFREQ
Q69. Before the coronavirus epidemic started, how often did you usually have direct contact with [*NAME*]?
A direct contact is when you **meet with this person in person** and when you exchange at least a few words, or when you have physical contact (e.g. handshake, embracing, kissing, contact sports).
**Please do not include times that you speak to them over the phone or internet.**
Select only one

1. Every day or almost every day
2. About once or twice a week
3. Every 2-3 weeks
4. About once per month
5. Less often than once per month
6. Never met them before
7. Prefer not to answer

*SA*
*ASK ALL*

Q74. We do ask you to individually include every contact you had between 5am yesterday and 5am today, but if you were unable to include every single contact (for instance, because you work in a shop and have a large number of contacts in a day), please could you indicate this?

For this section, as in previous sections, please report ONLY people that you have **had direct contact with**. By direct contact we mean with whom you exchanged at least a few words, or with whom you had physical contact *(e.g. a handshake, embracing, contact sports)*. *Please include people* with whom you **talked with while keeping a two-metre distance, wearing a mask, or otherwise social distancing.*

**Please DO NOT include people who were in the same location, but with whom you did not touch or exchange at least a few words.**

1. I individually included every person I had contact with.
2. I did not individually include every person I had contact with.
3. I did not have any contacts *(PN: ONLY SHOW IF NO CONTACTS LISTED AT Q63 I.E. ALL TEXT BOXES WERE LEFT EMPTY AND NO CONTACTS ARE CODED AS 1 AT Q62)*

*PN: SHOW IF Q74= 2*

*PN: SHOW IF Q74= 2 AND ASK (IF Q3=1,2,3 or Q3c=1 or 2)*
*SA*
SCRIPTER on each Q79a-d, Q80a-d, Q81a-d question, please add the following text above EACH question:
By direct contact we mean with whom you exchanged at least a few words, or with whom you had physical contact *(e.g. a handshake, embracing, contact sports)*. *Please include people with whom you talked with while keeping a two-metre distance, wearing a mask, or otherwise social distancing.*

Q79a. Approximately how many people did you have contact with **at work** and did not list individually?

*For this question, We are only interested in people who you met in person and **with whom you exchanged at least a few words***

*Please enter number of contacts for each age group and setting*

*PN MINIMUM VALUE SHOUD BE 0 AND MAX 2000*

1. Under 18 [INSERT TEXT BOX]
2. 18-64 [INSERT TEXT BOX]
3. 65+ [INSERT TEXT BOX]

*PN: SHOW IF Q74= 2 AND (IF q79a>0 for codes 1-3*
*SA*
Q79c. Were you able to take precautions (staying two metres apart, wearing a mask, or washing hands frequently) with most of the people you met **at work** to maintain a social distance?

1. Yes

2. No
3. Sometimes
4. Prefer not to answer

*PN: SHOW IF Q74= 2 AND (IF Q79a>0 for codes 1-3)*
*SA*

Q79d. How much time did you spend with each person on average of the people you met **at work.**
Select only one

1. Less than 1 minute
2 1 minutes or more, but less than 5 minutes
3. 5 minutes or more, but less than 15 minutes
4. 15 minutes or more, but less than 1 hour
5. 1 hour or more

*SHOW IF (Q74= 2) AND (Q3=8 or Q3b=1 or 2) AND sampletype=1*
*SA*
Q80a. Approximately how many people did you have contact with **at college/university** and did not list individually?

*For this question, We are only interested in people who you met in person and **with whom you exchanged at least a few words***

*Please enter number of contacts for each age group and setting*

*PN MINIMUM VALUE SHOUD BE 0 AND MAX 2000*

1. Under 18 [INSERT TEXT BOX]
2. 18-64 [INSERT TEXT BOX]
3. 65+ [INSERT TEXT BOX]

*PN: SHOW IF Q74= 2 AND (IF q80a>0 for codes 1-3)*
*SA*
Q80c. Were you able to take precautions (staying two metres apart, wearing a mask, or washing hands frequently) with most of the people you met **at college/university** to maintain a social distance?

1. Yes
2. No
3. Sometimes
4. Prefer not to answer

*PN: SHOW IF Q74= 2 AND (IF Q80a>0 for codes 1-3)*
*SA*

Q80d. How much time did you spend with each person on average of the people you met **at college/university**.

Select only one

1. Less than 1 minute
2 1 minutes or more, but less than 5 minutes
3. 5 minutes or more, but less than 15 minutes
4. 15 minutes or more, but less than 1 hour
5. 1 hour or more

*PN: SHOW IF Q74= 2*
*SA*
Q81a. Approximately how many people did you have contact with [**in another setting** *– add this text to the question wording (IF Q3=8 or Q3b=1 or 2) or (IF Q3=1,2,3 or Q3c=1 or 2) AND sampletype=1]* and did not list individually?

*For this question, We are only interested in people who you met in person and **with whom you exchanged at least a few words***

*Please enter number of contacts for each age group and setting*

*PN MINIMUM VALUE SHOUD BE 0 AND MAX 2000*

1. Under 18 [INSERT TEXT BOX]
2. 18-64 [INSERT TEXT BOX]
3. 65+ [INSERT TEXT BOX]

*PN: SHOW IF Q74= 2 AND (IF q81a>0 for codes 1-3)*
*SA*
Q81c. Were you able to take precautions (staying two metres apart, wearing a mask, or washing hands frequently) with most of the people you met [**in another setting** *– add this text to the question wording IF (Q3=8 or Q3b=1 or 2) or IF (Q3=1,2,3 or Q3c=1 or 2) AND sampletype=1]* to maintain a social distance?

1. Yes
2. No
3. Sometimes
4. Prefer not to answer

*PN: SHOW IF Q74= 2 AND (IF Q81a>0 for codes 1-3)*
*SA*

Q81d. How much time did you spend with each person on average of the people you met **in another setting** *– ADD THIS TEXT TO THE QUESTION WORDING IF (Q3=8 or Q3b=1 or 2) or IF (Q3=1,2,3 or Q3c=1 or 2) AND SAMPLETYPE=1]***.**
Select only one

1. Less than 1 minute

2 1 minutes or more, but less than 5 minutes
3. 5 minutes or more, but less than 15 minutes
4. 15 minutes or more, but less than 1 hour
5. 1 hour or more

***Parent Survey questions***

*ASK THE FOLLOWING QUESTIONS IF SAMPLETYPE=2, ALL QUESTIONS TO THE END*

**Individual preventive measures**

*ASK ALL*
*SA*
*MASK1*
QP54. Did [INSERT CHILD'S NAME from QP53a] use a face mask yesterday?
Select only one
1. Yes
2. No
3. Don't know

*ASK ALL*
*MA*
TRANSPORT
QP59. Did [INSERT CHILD'S NAME from QP53a] travel on any public transport yesterday?
Please tick all that apply
1. No *[SINGLE CODE]*
2. Train/tube
3. Bus/tram
4. Taxi, Minicab, Uber, or similar ride-hailing app
5. Aeroplane
6. Ferry/boat
7. Don't know *[SINGLE CODE]*

**ADDITIONAL CHILDRENS QUESTIONS**

*ASK IF Q26 = 1,2,3 OR 4*
Please answer the following questions about [INSERT CHILD'S NAME from QP53a]'s attendance at [pre-school or nursery] [*IF CHILD Q26 = 1*] OR [school] [*IF CHILD Q26 = 2*] OR [secondary school] [*IF Q26 = 3*] or [their university or other higher educational institution] [*IF Q26 = 4*]

*SA*
**QP61a.** Did [INSERT CHILD'S NAME from QP53a] attend [pre-school or nursery] [*IF CHILD Q26 = 1*] OR [school] [*IF CHILD Q26 = 2*] OR [secondary school] [*IF Q26 = 3*] or [their university or other higher educational institution]  [*IF CHILD Q26 = 4*] yesterday?

1. Yes
2. No, but it was open for my child
3. Not applicable as it was a weekend/holiday/day off
4. Not applicable as it was closed
5. Don't know
6. Prefer not to answer

*ASK IF QP61a = 1*
*SA*
QP61b. How many [children] [IF CHILD Q26 = 1,2, or 3] OR [people] [IF CHILD Q26 = 4] were in [INSERT CHILD'S NAME from QP53a]'s [pre-school or nursery] [ *IF CHILD Q26 = 1*] OR [class] [ *IF CHILD Q26 = 2*] OR [form] [*IF CHILD Q26 = 3*] OR [class/lecture] [*IF CHILD Q26=4*] yesterday?

If you are unsure of the exact answer, please enter your best guess. Please answer this question only for yesterday.

INSERT TEXT [*RESTRICT TO NUMBERS ONLY UP TO MAX 500]*
98. Don't know
99. Prefer not to answer

**Contact survey**

*INTRO SCREEN*
We will now ask you to remember who *[INSERT CHILD'S NAME FROM QP53a]* had been in contact with yesterday, between 5am yesterday and 5am today. We are only interested in direct contacts, which are **people who** *[INSERT CHILD'S NAME FROM QP53a]* **met in person** and with whom they exchanged at least a few words, or with whom ~~you~~ they had physical contact (e.g. a handshake, embracing, kissing, contact sports).
**Note that if** *[INSERT CHILD'S NAME FROM QP53a]* **only spoke to someone over the phone or internet, you should answer 'No' in this section.**

*ASK ALL*
*SA*
CONTACT1
QP62. Which of the following people did *[INSERT CHILD'S NAME FROM QP53a]* have direct contact with in person, between 5am yesterday and 5am today, in person?
*NOTE FOR SCRIPTER – PLEASE SCRIPT AS GRID*

*We are only interested in direct contacts, which are people who [INSERT CHILD'S NAME FROM QP53a] met in person and with whom ~~you~~ they exchanged at least a few words, or with whom they had physical contact (e.g. a handshake, embracing, kissing, contact sports).*
***Note that if [INSERT CHILD'S NAME FROM QP53a] only spoke to someone over the phone or internet, you should answer 'No' in this section.***
*Please include people they met while keeping a two-metre distance, wearing a mask, or otherwise social distancing.*

*ROWS:*
*INSERT ALL NAMES FROM Q21/Q21 IMPORT EXCLUDING SELECTED CHILD. ADD CODE 'You, the person completing the survey' TO THE LIST IF CHILD SELECTED TO ANSWER THIS SET OF QUESTIONSIF QSAMPLE=1 AND HHCOMPCONFRIM =2 THEN REMOVE ANYONE SELECTED AT HHCOMPREMOVE. QSAMPLE=1 AND HHCOMPCONFRIM =3 THEN ADD PEOPLE FROM HHCOMPADD*

*COLUMNS:*
1. Yes
2. No

*ASK ALL*
*WRITE IN*
CONTACT2

*INTRO SCREEN*
QP63. Now we would like to know what **other people, outside of your household,** *[INSERT CHILD'S NAME FROM QP53a]* had direct in person contact with between 5am yesterday and 5am today. This could include friends, family, classmates, or people they spoke to in shops and so on.
*We are only interested in people who [INSERT CHILD'S NAME FROM QP53a] met in person and **with whom they exchanged at least a few words, or with whom they had physical contact** (e.g. a handshake, embracing, contact sports)*.
*Please include people who* [INSERT CHILD'S NAME FROM QP53A] *met while keeping a two metre distance, wearing a mask, or otherwise social distancing.*

*NEW SCREEN*

Please write the nickname of each person *[INSERT CHILD'S NAME FROM QP53a]* had direct contact with outside your household. Note that this nickname is only needed to make it easier for you to complete the survey, so please pick a nickname that is easy to remember. Your individual responses will not be shared with anyone outside this survey.

It is easiest to list names in chronological order, e.g. When *[INSERT CHILD'S NAME FROM QP53a]* woke up they saw our neighbours Pete and Naomi. We then drove to school where they had lunch with their friends Jack and Deborah. On our way back home, we stopped to see their older cousin, Kate, at a café and spoke to the staff (assign a nickname to staff). Etc. You can add children in the classroom as C1, C2, C3 etc. if you are not sure of their names.
**Please do not list yourself or anyone that you listed as being a part of your household. Note that if *[INSERT CHILD'S NAME FROM QP53a]* only spoke to *someone* over the phone or internet, *they should not be included in this section*.**

*PN: SHOW IF Q20 IS NOT ONE OR IF NO-ONE IS SELECTED AT QP62*

*You have already indicated contact with the following household members, add additional contacts in the textboxes below:*

LIST HOUSEHOLD MEMBERS SELECTED AT QP62 AND THE PARTICIPANT FROM QP62a

*ALLOW BOXES TO INPUT NICKNAMES*
*PN SHOW 15 TEXT BOXES ON SCREEN AND ADD ADDITONAL BOXES ONE AT A TIME AS COMPLETED. LIMIT TO 100 BOXES IN TOTAL.*
*No-one/No other people [PN. ALLOW THIS TO BE SELECTED WHEN TEXT BOXES COMPLETED and Please show this option above the open text boxes.]*

*PN AT TOP OF PAGE WITH TEXT BOXES INCLUDE THE FOLLOWING.*

*PLEASE LIST ALL OTHER CONTACTS [INSERT CHILD'S NAME FROM QP53a] HAD OUTSIDE OF YOUR HOUSEHOLD, AS BEFORE PLEASE ASSIGN A **NICKNAME** TO EACH OF THESE CONTACTS*

*ASK THE FOLLOWING LOOP QP71, QP77, QP73 and QP69 TO TIME FOR EACH NAME GIVEN AT QP62. THIS SHOULD INCLUDE 'YOU, THE PERSON COMPLETING THE SURVEY WHEN THIS PERSON IS SELECTED AT QP62*
*ASK THE FOLLOWING LOOP QP66, QP67, QP68, QP71, QP77, QP73 and QP69TO TIME FOR EACH NAME GIVEN AT QP63*

*ASK FOR EACH PERSON AT QP63*
*SA*
QP66. Which of the following age groups does NAME fit into? Please give an estimate if you are not sure
Select only one

1. Under 1
2. 1-4
3. 5-9
4. 10-14
5. 15-19
6. 20-24
7. 25-34
8. 35-44
9. 45-54
10. 55-64
11. 65-69
12. 70-74
13. 75-79

14. 80-84
15. 85+
16. Don’t know
17. Prefer not to answer
18. This person is *[INSERT CHILD’S NAME FROM QP53A]*
19. This is me

*ASK ALL WHERE NAME GIVEN AT QP63 AND QP66 IS NOT EQUAL TO CODE 18 OR CODE 19*
*SA*
CONTACTGEN
QP67. As far as you know, which of the following describes how *[NAME]* thinks of themselves?
Select only one

1. _1 Male [KEEP]
2. _2 Female [KEEP]
3. _3 In another way [KEEP]
4. _4 Prefer not to answer [KEEP]
5. _5 Don’t know

*ASK ALL WHERE NAME GIVEN AT QP63 AND QP66 IS NOT EQUAL TO CODE 18 or CODE 19*
*SA*
CONTACTRELAT
QP68. What is [*NAME]*’s relationship to *[INSERT CHILD’S NAME FROM QP53A]?*
Select only one
1. They are family members not in our household
2. They are someone they see at nursery, pre-school, school, college or university
3. They are a babysitter, childminder, or nanny
4. They are friends
5. Other
6. Prefer not to answer

*ASK ALL WHERE NAME GIVEN AT QP62 OR QP63 AND QP66 IS NOT EQUAL TO CODE 18 or CODE 19 MA*

WHERE

QP71. And where did [INSERT CHILD'S NAME FROM QP53A] have direct contact with [YOU/NAME] between 5am yesterday and 5am today?

Please tick all that apply

1. At home (including at your door, in your garden, and within entrances to your home such as stairways, lifts, and corridors)
2. At someone else's house
3. At work
4. At a place of worship eg church, mosque or temple
5. On any form of public transport
6. At university, school, pre-school, or nursery
7. At a shop or market for essentials, eg a supermarket, grocery store, pharmacist
8. At a shop for non-essential items, eg a DIY shop, or a clothes or , furniture shop
9. At a place of entertainment such as a restaurant, bar, cinema
10. At a place for sports such as a gym or sports club/match
11. Outside, for example in a park, on the street or in the countryside
14. In a healthcare setting, eg hospital, GP, A&E, outpatient facility, dentist, physiotherapist, optometrist, etc
15. At a hair dresser, barber, nail salon, beauty parlor or similar location
12. Somewhere else (please specify)
16. Don't know [SINGLE CODE]

*ASK ALL WHERE NAME GIVEN AT QP62 OR QP63 AND QP66 IS NOT EQUAL TO CODE 18 or CODE 19 MA*

QP77

Which of the following apply to the direct contact *[INSERT CHILD'S NAME FROM QP53A*] had with [YOU/NAME] yesterday,

Select all that apply

1. They had physical contact (any sort of skin-to-skin contact such as e.g. hand shaking, embracing or kissing) [PN. DO NOT ALLOW TO SELECT WITH CODED 2, 3 AND 12]
2. They kept a distance of two or more metres apart the entire time [they/you] were with them [PN. DO NOT ALLOW TO SELECT WITH CODED 1, 3,12 ]
3. They kept a distance of one metre apart the entire time [they/you] were with them [PN. DO NOT ALLOW TO SELECT WITH CODED 1, 2,12]
12. They were less than one metre apart the entire time [they/you] were with them [PN. DO NOT ALLOW TO SELECT WITH CODED 1, 2, 3]
4. They wore a mask the entire time [they/you] were with them
5. They washed/sanitised their hands directly before contact
6. They washed/sanitised their hands directly after contact
9. Prefer not to say [PN: exclusive]
10. None of these [PN: exclusive]
11. Don't know [PN: exclusive]

*ASK ALL WHERE NAME GIVEN AT QP62 OR QP63 AND QP66 IS NOT EQUAL TO CODE 18 or CODE 19*
*SA*
TIME
QP72. Please estimate the total amount of time [INSERT CHILD'S NAME FROM QP53A] spent with [YOU/NAME] in person yesterday, this can include time when they were not in close proximity..
Select only one for each person

1. Less than 5 minutes
2. 5 minutes or more, but less than 15 minutes
3. 15 minutes or more, but less than 1 hour
4. 1 hour or more, but less than 4 hours
5. 4 hours or more
6. Don't know

*ASK ALL WHERE NAME GIVEN AT QP62 OR QP63 AND QP66 IS NOT EQUAL TO CODE 18 or CODE 19 AND If QP71= 1, 2, 3, 4, 6, 7, 9, 10 or 12, 14 or 15*
*MA*
QP73 Was the time [INSERT CHILD'S NAME FROM Q53A] spent with [YOU/NAME] yesterday inside and/or outside?
Please tick all that apply

Inside
Outside
Don't know [EXCLUSIVE]

*ASK ALL WHERE NAME GIVEN AT QP62 OR QP63 AND QP66 IS NOT EQUAL TO CODE 18 or CODE 19*
*SA*
CONTACTFREQ
QP69. Before the coronavirus epidemic started, how often did *[INSERT CHILD'S NAME FROM QP53A]* usually have direct contact with [YOU/*NAME*]? A direct contact is when [INSERT CHILD'S NAME FROM QP53A] **meets with this person in person** and when they exchange at least a few words, or when they have physical contact (e.g. a handshake, high five, embracing, touching while playing, contact sports). **Please do not include times that *[INSERT CHILD'S NAME FROM QP53A]* speaks to them over the phone or internet.**
Select only one

1. Every day or almost every day
2. About once or twice a week
3. Every 2-3 weeks
4. About once per month
5. Less often than once per month
6. Never met them before
7. Prefer not to answer

*SA*
*ASK ALL*

QP74. We do ask you to individually include every contact [INSERT CHILD'S NAME FROM QP53A] had between 5am yesterday and 5am today, but if you were unable to include every single contact (for instance, because they were at school and have a large number of contacts in a day), please could you indicate this?

For this section, as in previous sections, please report ONLY people that [*QP53a*] have **had direct contact with**. By direct contact we mean with whom you exchanged at least a few words, or with whom you had physical contact *(e.g. a handshake, embracing, contact sports)*. *Please include people who* *QP53a**talked with while keeping a two-metre distance, wearing a mask, or otherwise social distancing.*

**Please DO NOT include people who were in the same location, but with whom [QP53a*] did not touch or exchange at least a few words.**

Select only one

1. I individually included every person [INSERT CHILD'S NAME FROM QP53A] had contact with.
2. I did not individually include every person [INSERT CHILD'S NAME FROM QP53A] had contact with.
3. [INSERT CHILD'S NAME FROM QP53A] did not have any contacts (PN: ONLY SHOW IF NO CONTACTS LISTED AT QP63 I.E. ALL TEXT BOXES WERE LEFT EMPTY AND NO CONTACTS ARE CODED AS 1 AT QP62)

*PN: SHOW IF QP74= 2 AND Q25=1,2,3 for the selected child if sampletype=2*
*SA*
SCRIPTER on each QP79 a-d, QP80 a-d, QP81 a-d question, please add the following text above EACH question:
By direct contact we mean with whom *QP53a* exchanged at least a few words, or with whom *QP53a* had physical contact *(e.g. a handshake, embracing, contact sports)*. *Please include people who* *QP53a**talked with while keeping a two-metre distance, wearing a mask, or otherwise social distancing.*

QP79a. Approximately how many people did [INSERT CHILD'S NAME FROM QP53A] have contact with **at work** and you did not list individually?

*For this question, We are only interested in people who* [INSERT CHILD'S NAME FROM QP53A] *met in person and* ***with whom they exchanged at least a few words***

*Please enter number of contacts for each age group and setting*

*PN MINIMUM VALUE SHOUD BE 0 AND MAX 2000*

1. Under 18 [INSERT TEXT BOX]
2. 18-64 [INSERT TEXT BOX]
3. 65+ [INSERT TEXT BOX]

*PN: SHOW IF QP74= 2 AND (IF QP79a>0 for codes 1-3)*

*SA*
QP79c. Was [INSERT CHILD'S NAME FROM QP53A] able to take precautions (staying two metres apart, wearing a mask, or washing hands frequently) with most of the people they met **at work** to maintain a social distance?

1. Yes
2. No
3. Sometimes
4. Prefer not to answer

*PN: SHOW IF QP74= 2 AND (IF qp79a>0 for codes 1-3)*
*SA*

QP79d. How much time did [INSERT CHILD'S NAME FROM QP53A] spend with each person on average of the people they met **at work.**
Select only one

1. Less than 1 minute
2 1 minutes or more, but less than 5 minutes
3. 5 minutes or more, but less than 15 minutes
4. 15 minutes or more, but less than 1 hour
5. 1 hour or more

*PN: SHOW IF QP74= 2 AND IF Q26 = 1, 2, 3 OR 4 for the selected child if sampletype=2*
*SA*
QP80a. Approximately how many people did [INSERT CHILD'S NAME FROM QP53A] have contact with **at [college/university]** *SHOW IF Q26 = 3 OR 4*] OR **[pre- school or nursery]** [*Q26 = 1]* OR **[school]** [*Q26 = 2*] and you did not list individually?

*For this question, We are only interested in people who* [INSERT CHILD'S NAME FROM QP53A] *met in person and* ***with whom they exchanged at least a few words***

*Please enter number of contacts for each age group and setting*

*PN MINIMUM VALUE SHOUD BE 0 AND MAX 2000*

1. Under 18 [INSERT TEXT BOX]
2. 18-64 [INSERT TEXT BOX]
3. 65+ [INSERT TEXT BOX]

*PN: SHOW IF QP74= 2 AND (IF QP80a>0 for codes 1-3)*
*SA*
QP80c. Was [INSERT CHILD'S NAME FROM QP53A] able to take precautions (staying two metres apart, wearing a mask, or washing hands frequently) with most of the people they **at [college/university]**

*SHOW IF Q26 = 3 OR 4*] OR **[pre- school or nursery]** [*Q26 = 1]* OR **[school]** [*Q26 = 2*] and you did not list individually?

1. Yes
2. No
3. Sometimes
4. Prefer not to answer

*PN: SHOW IF QP74= 2 AND (IF qp80a>0 for codes 1-3)*
*SA*

QP80d. How much time did [INSERT CHILD'S NAME FROM QP53A] spend with each person on average of the people they met **at [college/university]** *SHOW IF Q26 = 3 OR 4*] OR **[pre- school or nursery]** [*Q26 = 1]* OR **[school]** [*Q26 = 2*] and you did not list individually?

Select only one

1. Less than 1 minute
2 1 minutes or more, but less than 5 minutes
3. 5 minutes or more, but less than 15 minutes
4. 15 minutes or more, but less than 1 hour
5. 1 hour or more

*PN: SHOW IF QP74= 2*
*SA*
QP81a. Approximately how many people did [INSERT CHILD'S NAME FROM QP53A] have contact with [**in another setting** *– add this text to the question wording IF Q25=1,2,3 for the selected child or IF Q26 = 1, 2, 3 OR 4 for the selected child]* and you did not list individually?

*For this question, We are only interested in people who [INSERT CHILD'S NAME FROM QP53A] met in person and **with whom they exchanged at least a few words***

*Please enter number of contacts for each age group and setting*

*PN MINIMUM VALUE SHOUD BE 0 AND MAX 2000*

1. Under 18 [INSERT TEXT BOX]
2. 18-64 [INSERT TEXT BOX]
3. 65+ [INSERT TEXT BOX]

*PN: SHOW IF QP74= 2 AND (IF QP81a>0 for codes 1-3)*
*SA*
QP81c. Was [INSERT CHILD'S NAME FROM QP53A] able to take precautions (staying two metres apart, wearing a mask, or washing hands frequently) with most of the people they met [**in another setting** *– add this text to the question wording IF Q25=1,2,3 for the selected child or IF Q26 = 1, 2, 3 OR 4 for the selected child]* to maintain a social distance?

1. Yes
2. No
3. Sometimes
4. Prefer not to answer

*PN: SHOW IF QP74= 2 AND (IF QP81a>0 for codes 1-3)*
*SA*

QP81d. How much time did [INSERT CHILD'S NAME FROM QP53A] spend with each person on average of the people they met [**in another setting** *– add this text to the question wording IF Q25=1,2,3 for the selected child or IF Q26 = 1, 2, 3 OR 4 for the selected child]***.**
Select only one

1. Less than 1 minute
2 1 minutes or more, but less than 5 minutes
3. 5 minutes or more, but less than 15 minutes
4. 15 minutes or more, but less than 1 hour
5. 1 hour or more

ASK ALL

SA
Q90
On a scale of 1 to 5 where 1 means far too lenient and 5 means far too strict, how would you describe the measures currently in place to control the Coranavirus (COVID-19) epidemic in the UK?
Select one only

1 Far too lenient
2 A bit too lenient
3 About right
4 A bit too strict
5 Far too strict
9 Don't know

MA
Q91
Which, if any, of these reasons apply to why you follow the current regulations and measures in place to control the Coranavirus (COVID-19) epidemic in the UK?

Select all that apply.

ROTATE ANSWERS EXCEPT EXCLUSIVE
1. I will be fined if I don't
2. It is the law
3. It is the right thing to do

4. To protect my family and friends from the virus
5. To protect myself for the virus
6. I want to - most of these measures make sense to me [PN: DO NOT ALLOW SELECTION WITH CODE 7]
7. I want to - some of these measures make sense to me [PN: DO NOT ALLOW SELECTION WITH CODE 6]
8. I do not follow the current regulations and measures [EXCLUSIVE]
9. Don't know [EXCLUSIVE]
