## Supplementary material for "CoMix: Changes in social contacts as measured by the contact survey during the COVID-19 pandemic in England between March 2020 and March 2021": Recruitment methods and social grades mapped to reported occupations

#### Survey Recruitment

Data was collected on behalf of our study team by Ipsos, an international market research company. Participants are recruited by Ipsos from a variety of sources to create panels that are representative of the population in which they are recruited. Ipsos primarily recruits through social networks, allowing them to target hard to recruit populations, and includes providing participant-relevant incentives for completing surveys. Other methods for recruitment include email lists, banners, website and text ads, co-registration, and search engine marketing. When necessary, they also partner with thoroughly vetted third party recruiters. Ipsos limits the number of surveys each participant is able to complete in a given time period and uses algorithms to detect fraud and remove users from the survey in real-time. For this survey, Ipsos recruited adults (ages 18 years and older) in census representative age bands. We compare our participants to census figures by age, gender, and household size

### Ipsos-MORI occupation to social grade map

| Main Earner Occupation | SOCIAL GRADE |
|---|---|
| 1100. Legislators and senior officials | 1. A - Upper middle class |
| 1210. Directors and chief executives | 1. A - Upper middle class |
| 1221. Production and operations department managers in agriculture, hunting, forestry and fishing | 3. C1 - Lower middle class |
| 1222. Production and operations department managers in manufacturing | 3. C1 - Lower middle class |
| 1223. Production and operations department managers in construction | 3. C1 - Lower middle class |
| 1224. Production and operations department managers in wholesale and retail trade | 3. C1 - Lower middle class |
| 1225. Production and operations department managers in restaurants and hotels | 3. C1 - Lower middle class |
| 1226. Production and operations department managers in transport, storage and communications | 3. C1 - Lower middle class |
| 1227. Production and operations department managers in business services | 3. C1 - Lower middle class |
| 1228. Production and operations department managers in personal care, cleaning and related services | 3. C1 - Lower middle class |
| 1229. Production and operations department managers not elsewhere classified | 3. C1 - Lower middle class |
| 1231. Finance and administration department managers | 2. B - Middle class |
| 1232. Personnel and industrial relations department managers | 2. B - Middle class |
| 1233. Sales and marketing department managers | 2. B - Middle class |
| 1234. Advertising and public relations department managers | 2. B - Middle class |
| 1235. Supply and distribution department managers | 2. B - Middle class |
| 1236. Computing services department managers | 2. B - Middle class |
| 1237. Research and development department managers | 2. B - Middle class |
| 1239. Other department managers not elsewhere classified | 2. B - Middle class |
| 1311. General managers in agriculture, hunting, forestry/ and fishing | 2. B - Middle class |
| 1312. General managers in manufacturing | 2. B - Middle class |
| 1313. General managers in construction | 2. B - Middle class |
| 1314. General managers in wholesale and retail trade | 2. B - Middle class |
| 1315. General managers of restaurants and hotels | 2. B - Middle class |
| 1316. General managers in transport, storage and communications | 2. B - Middle class |
| 1317. General managers of business services | 2. B - Middle class |
| 1318. General managers in personal care, cleaning and related services | 2. B - Middle class |
| 1319. General managers not elsewhere classified | 2. B - Middle class |
| 2110. Physicists, chemists and related professionals | 2. B - Middle class |
| 2120. Mathematicians, statisticians and related professionals | 2. B - Middle class |
| 2130. Computing professionals | 2. B - Middle class |
| 2141. Architects, town and traffic planners | 2. B - Middle class |
| 2142. Civil engineers | 2. B - Middle class |

| | |
|---|---|
| 2143. Electrical engineers | 2. B - Middle class |
| 2144. Electronics and telecommunications engineers | 2. B - Middle class |
| 2145. Mechanical engineers | 2. B - Middle class |
| 2146. Chemical engineers | 2. B - Middle class |
| 2147. Mining engineers, metallurgists and related professionals | 2. B - Middle class |
| 2148. Cartographers and surveyors | 2. B - Middle class |
| 2149. Architects, engineers and related professionals not elsewhere classified | 2. B - Middle class |
| 2210. Life science professionals | 2. B - Middle class |
| 2220. Health professionals (except nursing) | 2. B - Middle class |
| 2230. Nursing and midwifery professionals | 3. C1 - Lower middle class |
| 2300. Teaching professionals | 3. C1 - Lower middle class |
| 2410. Business professionals | 2. B - Middle class |
| 2420. Legal professionals | 2. B - Middle class |
| 2430. Archivists, librarians and related information professionals | 3. C1 - Lower middle class |
| 2440. Social science and related professionals | 2. B - Middle class |
| 2450. Writers and creative or performing artists | 2. B - Middle class |
| 2460. Religious professionals | 2. B - Middle class |
| 3110. Physical and engineering science technicians | 3. C1 - Lower middle class |
| 3120. Computer associate professionals | 3. C1 - Lower middle class |
| 3130. Optical and electronic equipment operators | 3. C1 - Lower middle class |
| 3140. Ship and aircraft controllers and technicians | 3. C1 - Lower middle class |
| 3150. Safety and quality inspectors | 4. C2 - Skilled working class |
| 3200. Life science and health associate professionals | 3. C1 - Lower middle class |
| 3300. Teaching associate professionals | 3. C1 - Lower middle class |
| 3410. Finance and sales associate professionals | 3. C1 - Lower middle class |
| 3420. Business services agents and trade brokers | 3. C1 - Lower middle class |
| 3430. Administrative associate professionals | 4. C2 - Skilled working class |
| 3440. Customs, tax and related government associate professionals | 3. C1 - Lower middle class |
| 3450. Police inspectors and detectives | 2. B - Middle class |
| 3460. Social work associate professionals | 4. C2 - Skilled working class |
| 3470. Artistic, entertainment and sports associate professionals | 3. C1 - Lower middle class |
| 3480. Religious associate professionals | 3. C1 - Lower middle class |
| 4100. Office clerks | 3. C1 - Lower middle class |
| 4200. Customer services clerks | 3. C1 - Lower middle class |
| 5110. Travel attendants and related workers | 3. C1 - Lower middle class |
| 5120. Housekeeping and restaurant services workers | 5. D - Working class |
| 5130. Personal care and related workers | 5. D - Working class |
| 5140. Other personal services workers | 5. D - Working class |
| 5160. Protective services workers | 3. C1 - Lower middle class |
| 5200. Models, salespersons and demonstrators | 4. C2 - Skilled working class |
| 6000. Skilled agricultural and fishery workers | 5. D - Working class |
| 7100. Extraction and building trades workers | 4. C2 - Skilled working class |
| 7210. Metal moulders, welders, sheet-metal workers, structural - metal preparers, and related trades workers | 4. C2 - Skilled working class |

| | |
|---|---|
| 7220. Blacksmiths, tool-makers and related trades workers | 4. C2 - Skilled working class |
| 7230. Machinery mechanics and fitters | 4. C2 - Skilled working class |
| 7240. Electrical and electronic equipment mechanics and fitters | 4. C2 - Skilled working class |
| 7310. Precision workers in metal and related materials | 4. C2 - Skilled working class |
| 7320. Potters, glass-makers and related trades workers | 4. C2 - Skilled working class |
| 7330. Handicraft workers in wood, textile, leather and related materials | 4. C2 - Skilled working class |
| 7340. Printing and related trades workers | 4. C2 - Skilled working class |
| 7410. Food processing and related trades workers | 4. C2 - Skilled working class |
| 7420. Wood treaters, cabinet-makers and related trades workers | 4. C2 - Skilled working class |
| 7430. Textile, garment and related trades workers | 4. C2 - Skilled working class |
| 7440. Pelt, leather and shoemaking trades workers | 4. C2 - Skilled working class |
| 8000. Plant and machine operators and assemblers | 5. D - Working class |
| 9100. Sales and services elementary occupations | 5. D - Working class |
| 9200. Agricultural, fishery and related labourers | 5. D - Working class |
| 9300. Labourers in mining, construction, manufacturing and transport | 5. D - Working class |
| 9888. Armed forces | 3. C1 - Lower middle class |
| 9991. Unemployed and not looking for a job / Long-term sick or disabled | 6. E - Lower level of subsistence |
| 9992. Pupil /Student/ in full time education | 6. E - Lower level of subsistence |
| 9993. Housewife | 6. E - Lower level of subsistence |
