## Supplementary material for "CoMix: Changes in social contacts as measured by the contact survey during the COVID-19 pandemic in England between March 2020 and March 2021": Original research proposal for the CoMix survey

### Measuring Behavioural Change during Covid-19 Epidemic

#### Introduction and Rationale

Appropriate response to Covid-19 requires an understanding of the epidemiological and behavioural drivers of disease transmission. Due to the dynamic nature of the epidemic and with the spread of the disease in the UK and Europe, analyses of the main factors affecting transmission need constant updating to provide relevant data-driven evidence to inform public health policies.

The predictive capabilities of mathematical models have been transformed through quantifying social contact rates. However, these data (such as the POLYMOD survey) were collected in “peace-time” and individuals’ behaviour may change during a severe epidemic.

Social mixing, a key determinant of COVID-19 spread, is likely to change over the course of the epidemic in relation to health risk perception. Therefore, timely data on social contacts and uptake of protective behaviours will be needed for shaping public health messaging and monitoring its effectiveness. These data will also be essential for improving transmission models and forecasting.

We will conduct a contact survey to assess changes in social mixing in relation with health risk perception. We will monitor contact patterns in the United Kingdom, Belgium and the Netherlands populations over the course of the epidemic, and assess psychological factors associated with contact. Behaviour tracking will be integrated in real-time models to improve the efficacy of public health interventions (e.g. messaging) and epidemic forecasts. These data will be used to refine transmission models, improve forecasting, and assess the effectiveness of social distancing measures.

We will simultaneously ask participants about knowledge, uptake and perceived efficacy of certain interventions (such as home-working). We will ask about cases in their social networks and the country, and measure how their behaviours change if/when they become unwell. Linking (at an individual-basis) changes in perceived and actual risk and changes in perceived efficacy of interventions with the uptake of different precautionary behaviours and individuals actual contact patterns will give unparalleled quantitative insight into how and why behaviours change during an epidemic.

Individuals will be regularly surveyed (every 2 weeks) to track if behaviours alter and what influences this. To our knowledge, this is the first time such data will be collected over the course of an epidemic. The development, piloting and analysis of the work will be led by Niel Hens (University of Antwerp) and John Edmunds (London School of Hygiene and Tropical Medicine). Real-time analysis and presentation of the data will be conducted by the LSHTM team, with additional input from partner organisations (RIVM, Bern, ISI Turin).

### Aim and objectives

This study aims to identify the impact of social distancing interventions implemented by the government and the perceived and actual threat of the Covid-19 epidemic on social mixing behaviours.

We will do so using the following objectives.

1. To collect accurate estimates of the following data during the Covid-19 outbreak
    a. Frequency of age-specific social contacts and their characteristics, at different points in the epidemic
    b. Household and individual level exposures to social distancing interventions
    c. Uptake of individual preventive measures
    d. Changes in attitudes and risk perceptions regarding the Covid-19 epidemic
2. To create social contact matrices that can be used to parametrize epidemiological transmission models
3. To assess the impact of different social distancing interventions on social mixing within the population
4. To use mathematical models to inform public health policy decisions related to Covid-19 outbreak.

### Study design

We will follow a cohort of adults prospectively during the course of the Covid-19 epidemic in the United Kingdom, Belgium and the Netherlands. We will ask questions about social contact behaviour, personal preventive measures, and exposure and impact of different social distancing interventions that may be implemented.

**Enrolment procedure**

We aim to survey 1500 adults per country every two weeks using an online survey, for a total of 16 weeks (8 surveys). With a ~12% dropout rate per wave of interviews, this would give us a final sample size of 500 individuals after the final wave. The study will be conducted among individuals who are already subscribed to internet panels from a market research company, in the United Kingdom, Belgium and the Netherlands. The company will ensure a sample that is representative of the national population on age of adults, sex, geographical location, and socio-economic status.

Panel members will be invited to participate by email on a random day of each week of data collection. In case of non-response, we will send up to two reminder emails for every week.

Participants will be paid an incentive for every survey they complete to compensate for their time..

**Measurements**

Panel members will answer questions regarding symptoms and impact of interventions (such as school or work closures) in the seven days preceding each survey, while answering questions about their social contacts for the day preceding the survey. Surveys will be conducted using an online webform, supplied by the market research company. Data will be stored on their secure encrypted servers.

**Survey**

During the first survey a respondent completes, we will ask certain household level characteristics including age, sex, household make-up, employment status, and income.

For every survey a respondent completes, we will ask about recent symptoms, Covid-19 tests, contact with known Covid-19 cases, and exposure and impact of different interventions, for each household member.

Impact of different social distancing interventions include:

- 1) whether household members have been asked to quarantine, isolate, work from home, or limit time spent at university (where applicable)
- 2) whether household members have had their workplace/university/school/nursery/pre-school close for at least one day in the preceding week, and
- 3) whether household members have been in quarantine, have been in isolation, have been working from home or unable to go into work/university/school/pre-school/nursery (where applicable).

We will also ask whether the respondent has been to certain events with large groups of people (such as a football match or a restaurant), whether they have taken any preventive measures (face-mask, handwashing, hand sanitizers), or whether they have used public transport.

We will also assess their attitudes and risk perceptions towards Covid-19 over time.

Finally, we will ask participants to complete a social contact survey, where we ask about all individuals contacted in the preceding day. For each contact, we will ask about their (estimated) age, sex, relationship to the respondent, general contact frequency, whether contact was physical or not, and how much time they spent with this contact.

### Statistics and data analysis

#### Sample size

We will measure social contact rates every 2 weeks in a panel of 500 individuals using diary-based methods. The precision around the average number of daily contacts in any age group will depend on the actual average number of daily contacts and degree of overdispersion in the distribution of the number of contacts. We estimate that with two records per individual we can detect a difference of 2.5 contacts between pairs of observations with 90% power, 5% Type 1 error, and 20% loss to follow up. Therefore repeated measurement of individuals should provide adequate power for further assessment of other endpoints.

#### Statistical analysis

We will use multilevel count models such as negative binomial regression techniques to assess the age-specific number of daily contacts, and control for confounding factors such as sex and household size. We will perform a stratified analysis by contact type (all contacts, only physical contacts, only nonphysical contacts) and setting (e.g. household, workplace, school, social settings).

The effect of any social distancing control on the average number of contacts per day will be estimated accounting for age in the multi-level model. Further to this, the individual social distancing measures will be evaluated separately. Effect modification by age and sex will be explored as appropriate.

In order to develop social contact matrices that can be used to parametrize epidemiological transmission models. We will estimate age-specific transmission parameters by correcting the age-specific daily frequency of social contacts between different age groups for the reciprocity of contacts, adjusting for the population age distribution. These estimates will be used in ongoing and future response work conducted by LSHTM and partners to help inform public policy.

We will conduct descriptive analyses of household and individual level exposures to social distancing interventions, and uptake of individual preventive measures. Frequencies will be presented in tables and exploratory analysis of the association of these measures on the number of contacts will be performed in the multilevel count models with a random effect for participants. Attitudes and risk perceptions relating to the Covid-19 epidemic will be assessed over time visually and using descriptive statistics.

We will estimate current transmission dynamics in the UK using the contact matrices, imputing data for children and adolescents under 18 years of age using data from the POLYMOD study. We will report the mean number of contacts and the reproduction number resulting from the change in social mixing and an assumed basic reproduction number.

We will combine the survey data with the results of other concurrent and historical surveys as relevant to improve the strength of our results.

### Regulatory details

We apply for ethics approval at LSHTM Research Ethics Committee. Further ethics approval will be sought where required.

Informed consent will be obtained prior to any respondent completing the first survey. All panel members will already have agreed to be included in the internet panel and to be approached for online research. The data will be collected using the public interest as the lawful basis for collecting the data as defined by GDPR article 14. We will inform participants about the rationale and topic of the survey, and the incentive they will receive upon completion. They will confirm their eligibility and understanding of their rights, access to survey responses, and planned public dissemination of the anonymised data from the survey as presented in the privacy notice before the survey will start.

The right of the participant to refuse to participate without giving reasons will be respected at all times. All participants are free to withdraw at any time from the survey without giving reasons.

The survey will be translated to Dutch, Flemish, and French, and back translated to English to ensure the meaning of each question will remain as intended.

All data will be stored securely and confidentiality will be protected in accordance with the Data Protection Act.

### Dissemination of findings

Findings will rapidly be made available on online sources within several days of completion of every wave of the survey. This will allow for rapid dissemination to improve public health policy decision making, specifically regarding putting interventions in place  or descaling interventions. The anonymised data will be made publicly available.
